# Serial cross-sectional estimation of vaccine and infection-induced SARS-CoV-2 sero-prevalence in children and adults, British Columbia, Canada: March 2020 to August 2022

**DOI:** 10.1101/2022.09.09.22279751

**Authors:** Danuta M Skowronski, Samantha E Kaweski, Michael A Irvine, Shinhye Kim, Erica SY Chuang, Suzana Sabaiduc, Mieke Fraser, Romina C Reyes, Bonnie Henry, Paul N Levett, Martin Petric, Mel Krajden, Inna Sekirov

## Abstract

**Background:** We chronicle SARS-CoV-2 sero-prevalence through eight cross-sectional sero-surveys (snapshots) in the Lower Mainland (Greater Vancouver and Fraser Valley), British Columbia, Canada from March 2020 to August 2022.

**Methods:** Anonymized-residual sera were obtained from children and adults attending an outpatient laboratory network. Sera were tested with at least three immuno-assays per snapshot to detect spike (S1) and/or nucleocapsid protein (NP) antibodies. Sero-prevalence was defined by dual-assay positivity, including any or infection-induced, the latter requiring S1+NP antibody detection from January 2021 owing to vaccine availability. Infection-induced estimates were used to assess the extent to which surveillance case reports under-estimated infections.

**Results:** Sero-prevalence was ≤1% by the 3^rd^ snapshot in September 2020 and <5% by January 2021 (4^th^). Following vaccine roll-out, sero-prevalence increased to >55% by May/June 2021 (5^th^), ∼80% by September/October 2021 (6^th^), and >95% by March 2022 (7^th^). In all age groups, infection-induced sero-prevalence remained <15% through September/October 2021, increasing through subsequent Omicron waves to ∼40% by March 2022 (7^th^) and ∼60% by July/August 2022 (8^th^). By August 2022, at least 70-80% of children ≤19 years, 60-70% of adults 20-59 years, but ∼40% of adults ≥60 years had been infected. Surveillance case reports under-estimated infections by 12-fold between the 6^th^-7^th^ and 92-fold between the 7^th^-8^th^ snapshots.

**Interpretation:** By August 2022, most children and adults had acquired SARS-CoV-2 vaccine and infection exposures, resulting in more robust hybrid immunity. Conversely the elderly, still at greatest risk of severe outcomes, remain largely-dependent on vaccine-induced protection alone, and should be prioritized for additional doses.

## INTRODUCTION

As lead public health agency in the westernmost province of Canada, the BC Centre for Disease Control (BCCDC) has long established a sero-surveillance protocol to monitor population susceptibility to emerging or re-emerging respiratory viruses. The approach was first deployed during the 2009 influenza A(H1N1) pandemic to monitor change in sero-prevalence across successive pandemic waves and roll-out of the mass vaccination campaign[1-7]. The methodology utilizes cross-sectional sampling of community-based (anonymized, residual) sera from children and adults of all ages presenting for outpatient laboratory testing in the most densely-populated Lower Mainland (Greater Vancouver and Fraser Valley) region of BC where ∼60% of the provincial population (of ∼5 million) resides (**Supplementary_Figure_1**)[8,9]. For viruses (such as influenza) for which serological correlates of protection are established, sero-prevalence estimates are used to assess progress toward community immunity[1-4]. For viruses without established antibody thresholds of protection (such as SARS-CoV-2), findings inform the proportion of the population remaining immunologically naïve or, conversely, the accumulating proportion with evidence of infection- or vaccine-induced priming to the novel pathogen.

In response to SARS-CoV-2 emergence, BCCDC launched its first sero-survey in March 2020, just prior to World Health Organization (WHO) declaration of a COVID-19 pandemic[10]. Since then, BCCDC has conducted eight sero-surveys, spanning seven pandemic waves and the trajectory of SARS-CoV-2 vaccine roll-out. We present the change in vaccine- and infection-induced SARS-CoV-2 sero-prevalence among children and adults in the Lower Mainland, BC, from March 2020 to August 2022. Infection-induced estimates are further used to assess period-specific attack rates and the extent of under-ascertainment by surveillance case reports.

## METHODS

### Sero-survey context

The timeline of SARS-CoV-2 sero-surveys (“snapshots”) in relation to pandemic waves is shown in **Figure_1**[11,12], with relevant public health context provided in **Supplementary_Material_1**. Publicly-funded nucleic-acid amplification testing (NAAT) was broadly-available for symptomatic individuals through most of the pandemic until January 2022 when access became limited to high-risk individuals and use of rapid antigen tests (RATs) was greatly expanded. NAAT (but not RAT) confirmed and epidemiologically-linked cases are reportable to the BCCDC, but reports to date have excluded re-infections[11]. Apart from temporary school closure between March and June (epi-weeks 12-22) 2020, children were able to attend scheduled classes in person. Masking within indoor public settings was mandated for all individuals ≥12 years old (except in schools) beginning November (epi-week 47) 2020, extended to schools for the 2021-22 academic year, including children ≥5 years from October (epi-week 43) 2021. Two spike (S1)-based mRNA vaccines, first authorized in Canada in mid-December 2020[13], were initially targeted to long-term care residents and healthcare workers with age-based prioritization of the oldest community-dwelling adults in BC beginning March 2020. Public health measures were lifted in February (epi-week 7) along with mask mandates in March (epi-week 10) and vaccine cards for social settings in April (epi-week 14) 2022.

**Figure 1.**
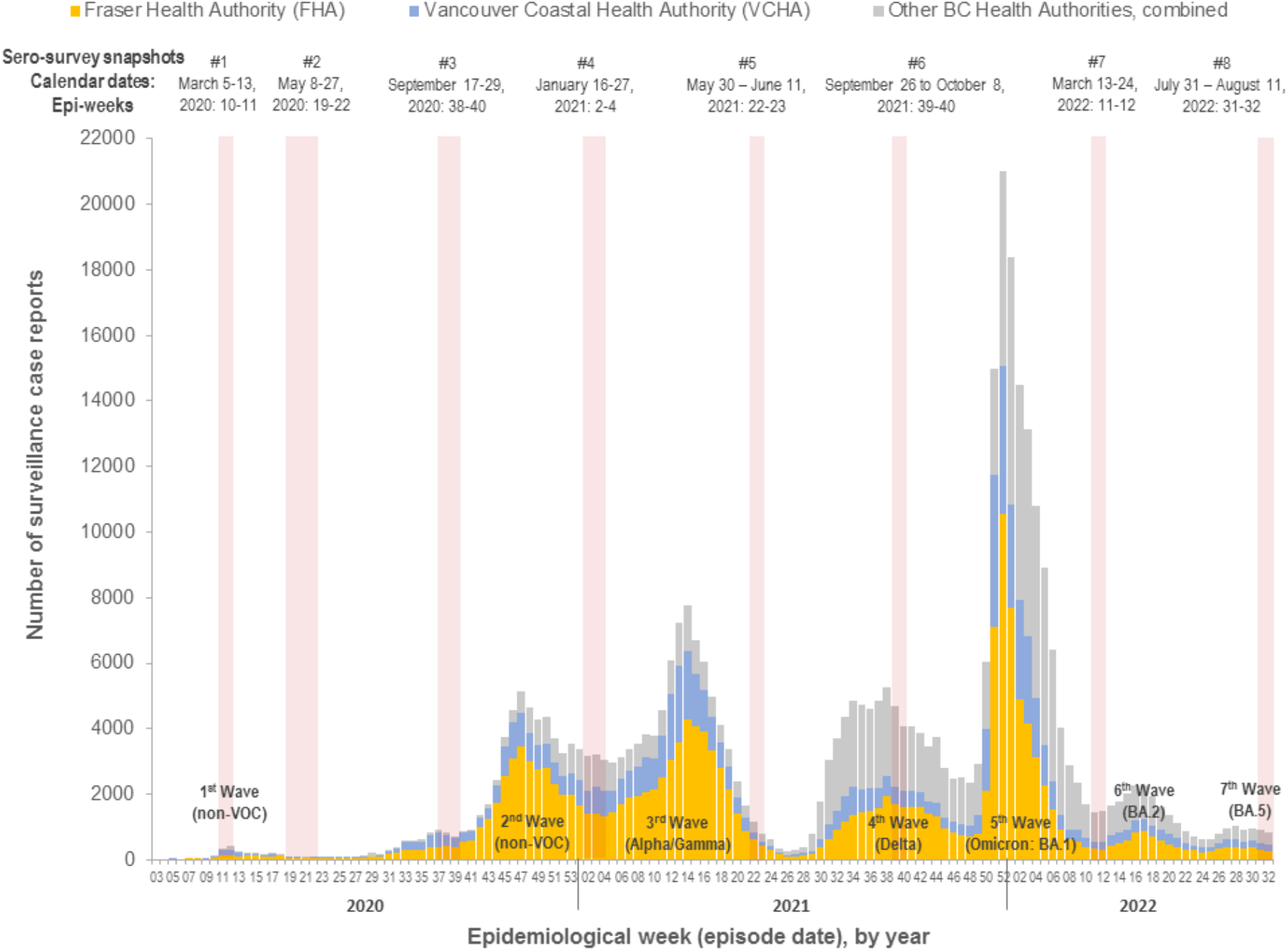
Epidemic curve of provincial surveillance case reports by epidemiological week with timing of sero-surveys overlaid, British Columbia, Canada Surveillance case reports as recorded by the BC Centre for Disease Control from Fraser Health Authority and Vancouver Coastal Health Authority. Other health regions include combined reports and populations of Interior Health Authority, Northern Health Authority and Vancouver Island Health Authority. Surveillance case reports reflect tallies of laboratory-confirmed, laboratory probable and epidemiologically-linked cases as of the August 2022 snapshot as posted on the BCCDC website [11] but excluding cases from out-of-province and care facility (long term care, assisted living, independent living) residents to reflect community-dwelling Lower Mainland residents. Surveillance case reports are timed by episode date defined hierarchically by onset date, or if unavailable then specimen collection or laboratory result date. Through the current study period spanning to August 2022, official BCCDC surveillance case report tallies include only the first (primary infection) and exclude re-infections [11]. Epidemic waves are enumerated with predominant variant of concern (VOC) contribution during specified waves.

### Sampling approach

Sera were sampled from the Lower Mainland, BC with eligible municipalities shown in **Supplementary Figure 1**. Two Lower Mainland health authorities (HA) are responsible for surveillance reporting including Fraser Health Authority (FHA:∼1.9 million) and Vancouver Coastal Health Authority (VCHA:∼1.2 million)[8,9]. Anonymized-residual sera were provided to BCCDC under legal order of the Provincial Health Officer by LifeLabs, the only outpatient laboratory network in the Lower Mainland. Individuals presenting for SARS-CoV-2 antibody testing and those residing in care or correctional facilities were excluded. For the first two snapshots, 100 sera per age group were sought but thereafter sampling increased to 200, equally male and female, per age groups: <5,5-9,10-19,20-29,30-39,40-49,50-59,60-69,70-79, and ≥80 years[14]. The study was approved by the University of British Columbia Clinical Research Ethics Board (H20-00653).

### Serological testing

At each snapshot, at least three commercially-available chemi-luminescent assays (CLIAs), targeting S1 or nucleocapsid protein (NP), were used in serological testing[15,16]. Sero-positivity was defined by signal above the cut-off threshold on at least two CLIAs. Prior to vaccine availability, all sero-positivity was assumed infection-induced. From the January 2021 sero-survey, infection-induced sero-positivity required that at least one of the two positive assays include anti-NP detection (i.e. S1+NP antibody).

Serological testing was undertaken real-time with adjustment based on evolving understanding of assay characteristics and their local availability(**Table_1**). For the first three snapshots in 2020, sera were screened with the Ortho (S1 total antibody: “Ortho-S1”) and Abbott (NP IgG: “Abbott-NP”) assays at the BCCDC Public Health Laboratory (PHL). Specimens positive on either CLIA were then also tested with the Siemens (S1 receptor-binding domain IgG/IgM: “Siemens-S1-RBD”) assay. With vaccine roll-out, anti-NP detection became more important but concerns related to waning and reduced anti-NP sensitivity also arose, particularly for Abbott-NP[17-21]. For the 4^th^ and 5^th^ snapshots, testing was supplemented, as volume permitted, by additional Roche (NP total antibody: “Roche-NP”) testing at the Providence Health Care Special Chemistry Laboratory. In the event a specimen was Abbott/Roche-NP discordant, NP positivity on either assay was accepted as indicative of infection. For the 6^th^ and 7^th^ snapshots, all sera were tested by Ortho-S1, Siemens-S1-RBD and Roche-NP assays. By the 8^th^ snapshot, the BCCDC PHL no longer offered Ortho-S1 testing, so it was replaced with the Abbott (S1-RBD IgG) assay[15,16,22].

**Table 1.**
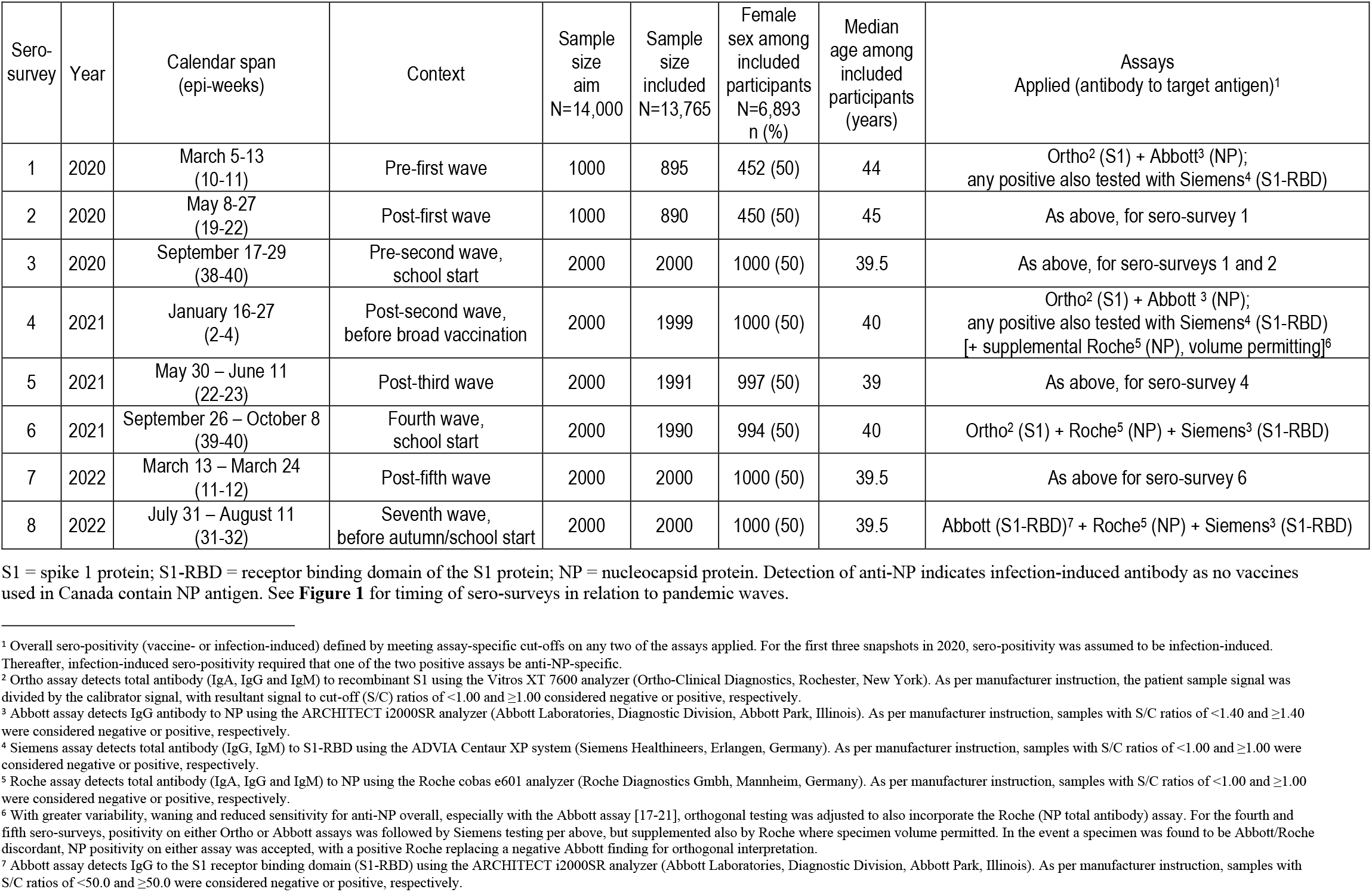
Sero-prevalence surveys: timing, sample size, participant characteristics, and assays applied

### Statistical analyses

#### Sero-prevalence

Sero-prevalence estimates based on dual-assay sero-positivity are presented as “any” (vaccine- and/or infection-induced) and “infection-induced” (with/without vaccination). Crude tallies and sero-prevalence estimates with 95% confidence intervals (CIs) are provided but primary sero-prevalence estimates are based upon Bayesian logistic regression modelling[23,24]. Prevalence log-odds were modeled for each snapshot independently, incorporating a hierarchical distribution for each age group, sex, and HA cell. High sensitivity and specificity have been reported for each of the CLIAs[15,16,22,25], but evaluations have not typically addressed potential variation by vaccination status, time since exposure, severity, age and/or target group[26-28]. Similar to other public health agencies[29,30], we do not adjust for sensitivity or specificity in primary analyses, but explore their effects as detailed in **Supplementary_Material_2**. Bayesian modelling was implemented in the Stan probabilistic programming language using a Hamiltonian Monte Carlo method to generate samples of the posterior. Eight thousand samples were generated across four chains including 4000 warm-up samples. Sero-prevalence estimates with 95% credible intervals (CrI) for each age group, sex, and HA stratum were sampled from the posterior with the post-stratification method[31]. These were applied to FHA and VCHA population estimates[8,9] to produce age-, sex- and HA-standardized cumulative and period-specific sero-prevalence estimates, the latter reflecting attack rates between snapshots. Visual inspection and the R-hat statistic were used to determine convergence and mixing of the chains.

#### Surveillance under-ascertainment ratio

Infection-induced sero-prevalence estimates and population census statistics were used to model the number of infections in the Lower Mainland. Surveillance under-ascertainment ratios (SUARs) with 95%CrIs were derived by dividing the modeled number of infections by the actual number of surveillance case reports from FHA+VCHA. Cumulative and period-specific SUARs were derived, the latter assuming no previously-infected individuals were re-infected as per surveillance case reporting in BC. Additional methodological details are provided in **Supplementary_Material_3**.

## RESULTS

### Participant profiles

Of 14,000 sera collected across eight sero-surveys, 13,765 (98%) contributed with 235 excluded due to insufficient volume(**Table_1**). Of excluded sera, 215 (91%) were collected during the earliest March and May 2020 snapshots, and most (189; 80%) were children <10 years. The number excluded, tested and dual-assay positive is detailed in **Supplementary_Table_1**.

Half of participants were female, reflecting the sampling framework and source population(**Supplementary_Table_2**). Median age was also similar to the source population (∼40 years), slightly older for the two earliest snapshots (∼44-45years). The percentage distribution by age group was within 5-10% (absolute) of the source population. However, more sera were collected from FHA compared to the Lower Mainland distribution (60%), notably among children <10 years(**Supplementary_Table_3**). As shown in **Figure_1**, FHA also reported more (about two-thirds) of Lower Mainland SARS-CoV-2 cases.

### Sero-prevalence and SUAR estimates

Primary Bayesian adjusted SARS-CoV-2 sero-prevalence estimates by snapshot are displayed for any and infection-induced antibody overall in **Figure_2**, and by age group in **Figure_3** and **Figure_4**. Crude tallies and precise Bayesian estimates with CrIs, including stratification by age, sex and HA are provided in **Supplementary_Tables_4-5**. Estimates stratified simultaneously by age group and sex are provided in **Supplementary_Tables_6-7**. Crude single-assay findings are provided for reference in **Supplementary_Table_8**.

**Figure 2.**
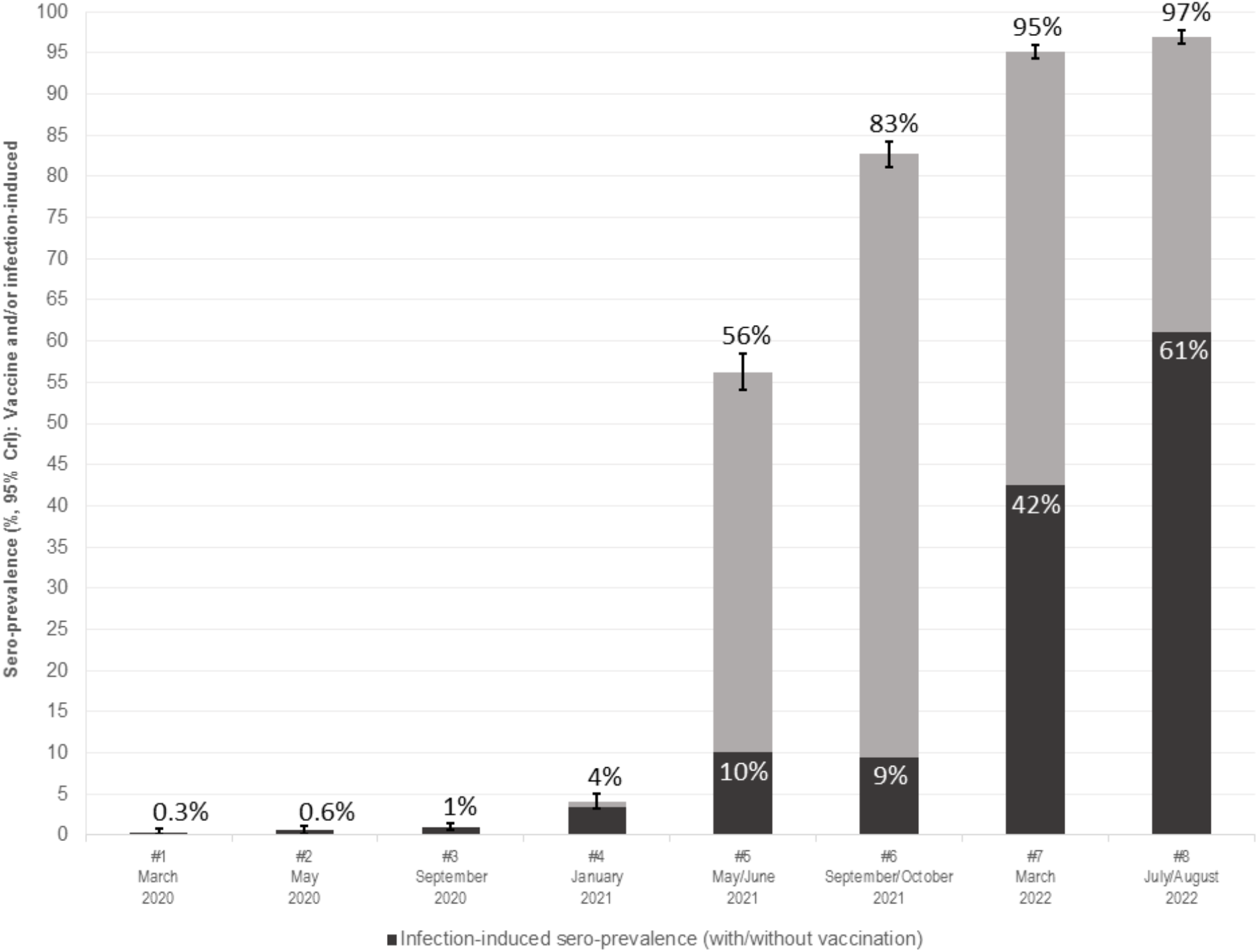
Sero-prevalence (any and infection-induced) overall, by sero-survey, March 2020 to July/August 2022, Lower Mainland, British Columbia, Canada 95% CrI = 95% credible interval (around the estimate of “any” which represents vaccine and/or infection induced sero-prevalence). Darker bars are infection-induced sero-prevalence. Displayed sero-prevalence estimates are based upon Bayesian model estimation, standardized for age, sex and health authority (HA) within the Lower Mainland (Fraser HA and Vancouver Coastal HA), British Columbia, Canada. See **Supplementary Table 4 (any) and Supplementary Table 5 (infection-induced)** for crude tallies and Bayesian estimates with CrIs.

**Figure 3.**
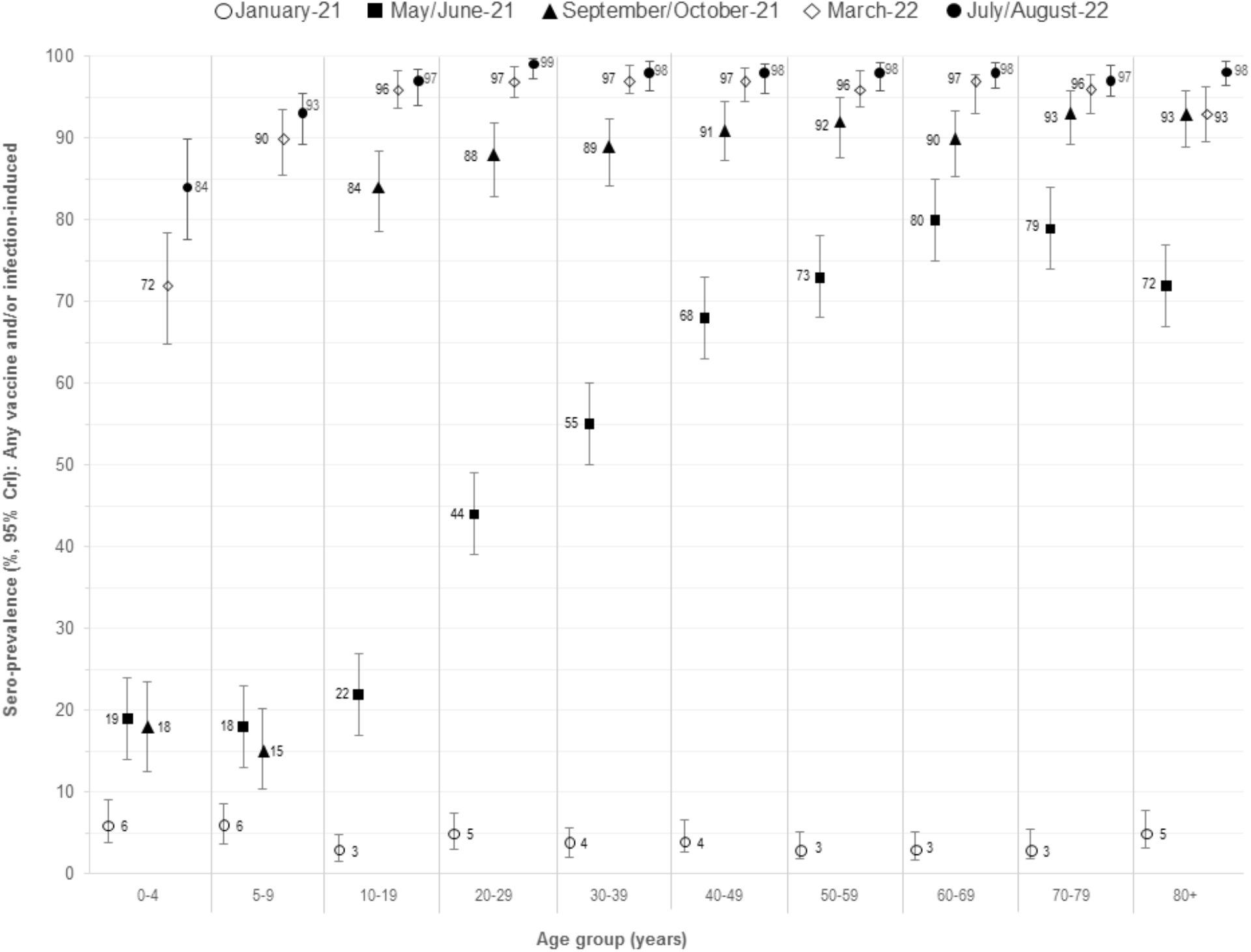
Change in any sero-prevalence by age group and sero-survey, March 2020 to July/August 2022, Lower Mainland, British Columbia, Canada 95% CrI = 95% credible interval Displayed sero-prevalence estimates are for any (vaccine and/or infection-induced) antibody, stratified by age group and based upon Bayesian model estimation, standardized for age, sex and health authority (HA) within the Lower Mainland (Fraser HA and Vancouver Coastal HA), British Columbia, Canada. See **Supplementary Table 4** for crude tallies and Bayesian estimates with CrIs.

**Figure 4.**
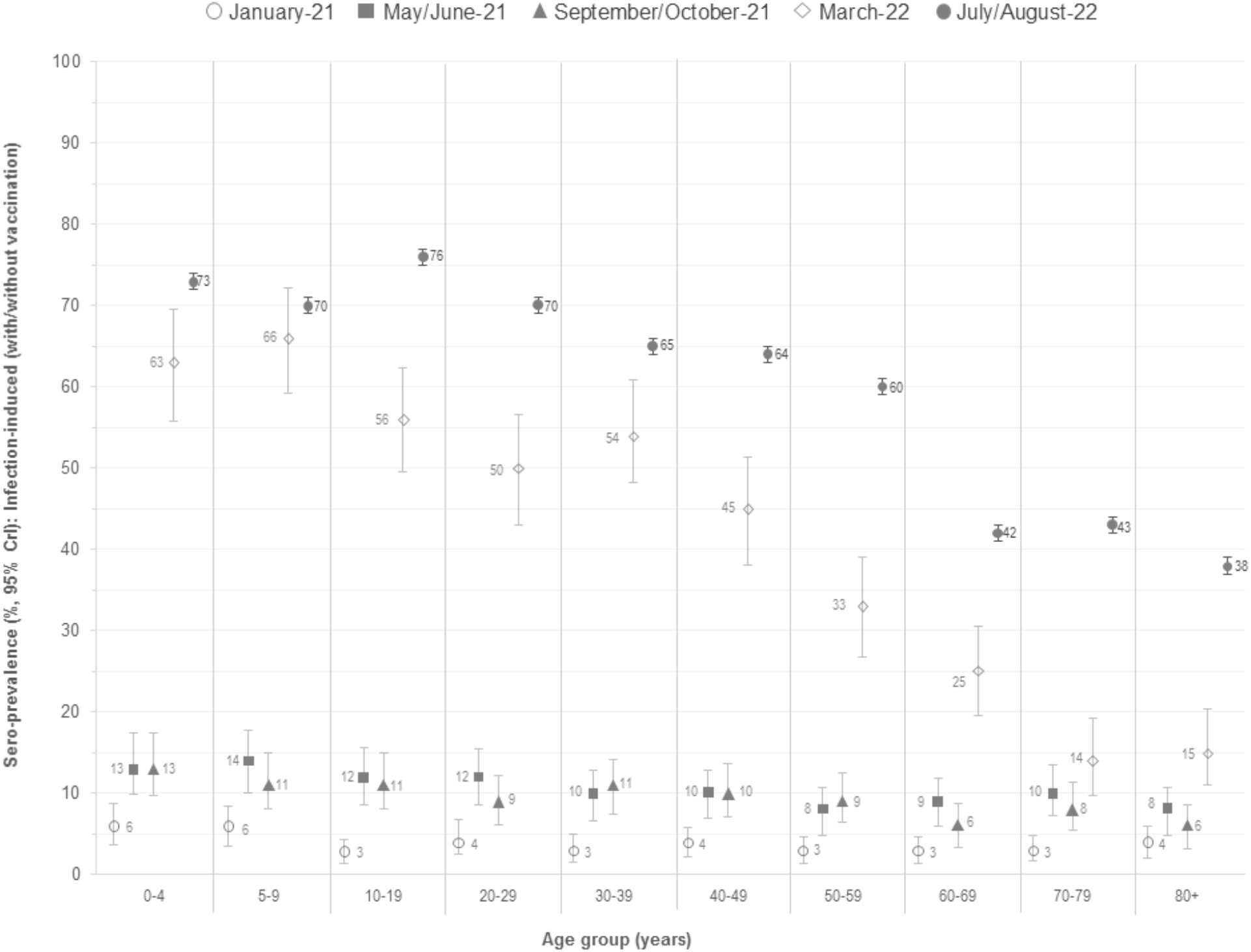
Change in infection-induced sero-prevalence by age group and sero-survey, March 2020 to July/August 2022, Lower Mainland, British Columbia, Canada 95% CrI = 95% credible interval Displayed sero-prevalence estimates are for infection-induced antibody, stratified by age group and based upon Bayesian model estimation, standardized for age, sex and health authority (HA) within the Lower Mainland (Fraser HA and Vancouver Coastal HA), British Columbia, Canada. See **Supplementary Table 5** for crude tallies and Bayesian estimates with CrIs.

Cumulative surveillance case reports since the beginning of the pandemic and SUARs by the 4^th^ (January 2021) through 8^th^ (July/August 2022) snapshots are provided in **Supplementary_Table_9**. Period-specific case reports, attack rates and SUARs between each consecutive snapshot are shown in **Supplementary_Table_10**, and by age group between the 6^th^-7^th^ and 7^th^-8^th^ snapshots in **Figure_5** and **Supplementary_Table_11**.

**Figure 5.**
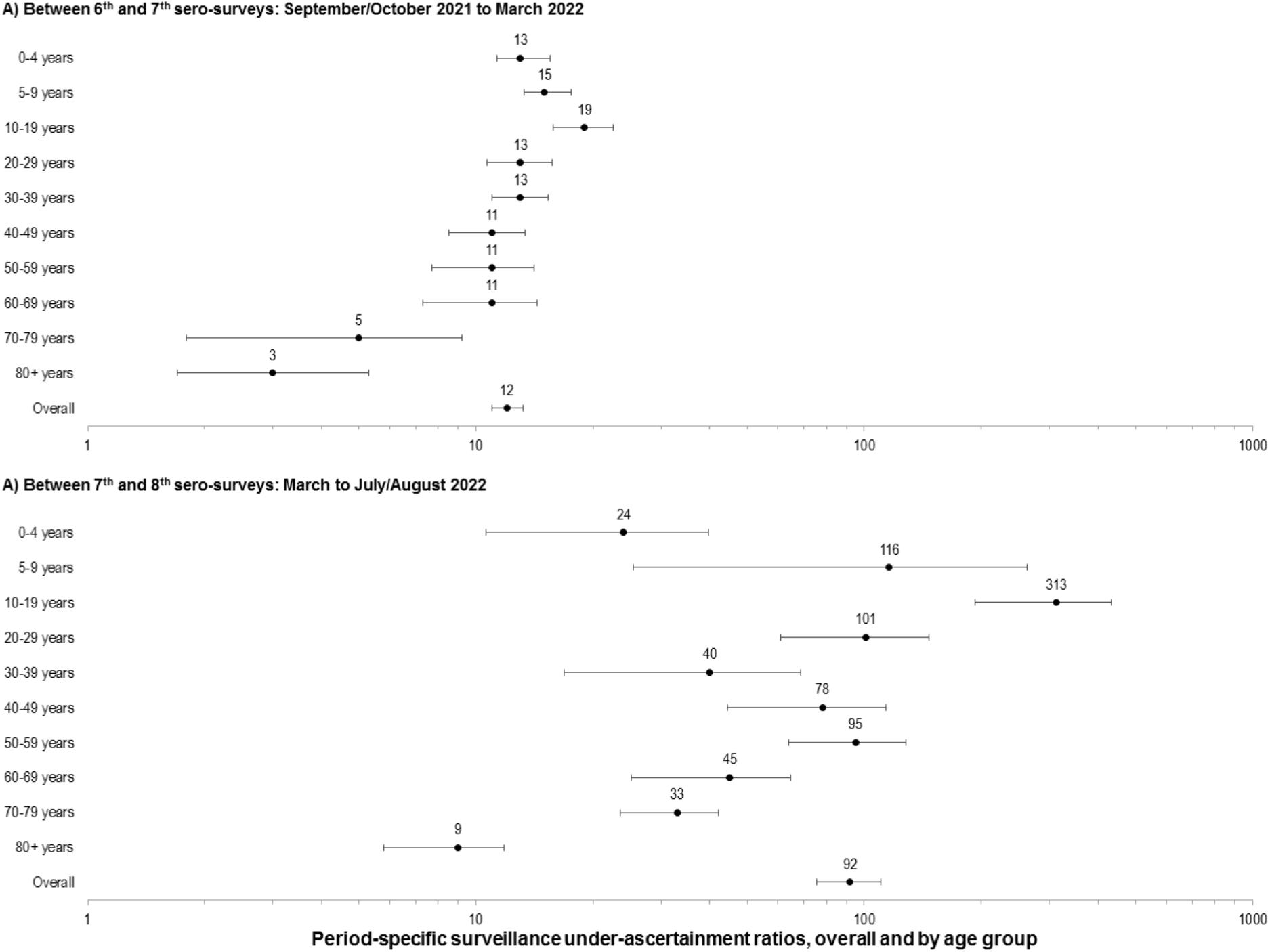
Period-specific surveillance under-ascertainment ratios, overall and by age group, 6^th^-7^th^ (September/October 2021 to March 2022) and 7^th^-8^th^ sero-surveys (March to July/August 2022), Lower Mainland, British Columbia, Canada 95% CrI = 95% credible interval. Displayed are the period-specific surveillance under-ascertainment ratios (SUARs) (log scale) overall and by age group. For precise values including period-specific case reports, attack rates and SUARS see **Supplementary Table 10** (overall period-specific, between all consecutive sero-surveys) and **Supplementary Table 11** (period-specific, by age group, 6^th^-7^th^ and 7^th^-8^th^ sero-surveys).

#### Sero-prevalence by snapshot

Estimates of any sero-prevalence did not exceed 1% through the first three snapshots to September 2020. By January 2021 (4^th^ snapshot), the estimate of any sero-prevalence was still <5% with minor difference from the infection-induced estimate consistent with low vaccine coverage. By May/June 2021 (5^th^), >55% had evidence of SARS-CoV-2 priming, driven by vaccine coverage provincially (∼60%), with ∼10% considered infection-induced. By September/October 2021 (6^th^), the estimate of any sero-prevalence was ∼80% (similar to vaccine coverage); whereas, infection-induced sero-prevalence remained stable at ∼10%, reflecting trough SARS-CoV-2 reports during the summer 2021(**Figure_1**). By March 2022 (7^th^), 95% of participants had detectable SARS-CoV-2 antibody (consistent with ∼85% vaccine coverage), with ∼40% for whom infection contributed. The latter reflects the greatest period-specific attack rate (33%) across the series, driven by the intervening Omicron BA.1 wave. By July/August 2022 (8^th^), ∼60% of participants were infected, a further ∼20% increase associated with the spring/summer Omicron BA.2 and BA.4/5 waves.

#### Sero-prevalence by HA, sex, age group and snapshot

Estimates of any sero-prevalence differed by <1% (absolute) by sex or HA and all infection-induced estimates were within 5% by sex and 10% by HA (consistently higher for FHA). When further stratified by age, all estimates remained within 5-10% between males and females (with inconsistent direction).

By May/June 2021, the effect of age-based vaccine roll-out was evident with sero-prevalence generally increasing with increased age (except among the very old who were vaccinated earliest). Consistent with later vaccine eligibility for children 5-11 years (from November 2021), estimates of any and infection-induced sero-prevalence were comparable among children <10 years (within 1% absolute) through September/October 2021. Children <5 years became vaccine-eligible in August 2022 with <1-2% vaccinated by the corresponding snapshot, but their estimates of any versus infection-induced sero-prevalence were more divergent in both March (72% vs. 63%) and July/August (84% vs. 73%) 2022. Since any sero-prevalence in that age group would necessarily be infection-induced, discrepancies may be a measure of reduced anti-NP detection.

In all age groups, cumulative infection-induced sero-prevalence remained <6% through January 2021 and <15% through September/October 2021 (6^th^). By the March (7^th^) or July/August (8^th^) 2022 snapshots, cumulative infection rates were much higher, showing gradual decrease with increasing age. For individuals <50 years, the greatest period-specific attack rates were between the 6^th^-7^th^ snapshots; whereas, elderly adults ≥70 years experienced their highest attack rates between the 7^th^-8^th^ snapshots.

By March 2022, at least 60-70% of children ≤19years had been infected and by July/August at least 70-80% had been infected. Of note, children <10years with the highest period-specific attack rates of any age group between the 6^th^-7^th^ snapshots (≥50%) subsequently had the lowest period-specific attack rates between the 7^th^-8^th^ (<10%). While just about 15% of elderly adults ≥70 years were had been by March 2022, this increased to ∼40% by July/August 2022. Adults 70-79 years with the lowest period-specific attack rates of any age group between the 6^th^-7^th^ snapshots (<10%) then had the highest period-specific attack rates between the 7^th^-8^th^ (∼30%).

#### SUARs

Cumulatively, surveillance case reports under-estimated infections in the Lower Mainland by ∼5-fold by March and ∼8-fold by July/August 2022. Period-specific SUARs ranged 3-8 up to September/October 2021 (higher at the first period when sero-prevalence was very low and CrIs were wide). With the subsequent Omicron waves and changes in testing practices, surveillance reporting more substantially under-estimated infections with SUAR of 12.1 (95%CrI: 11.0, 13.2) between September/October 2021 and March 2022 (6^th^-7^th^) and 91.9 (95%CrI: 75.3, 110.1) between March and July/August 2022 (7^th^-8^th^). Both cumulative and period-specific SUARs were generally highest in children and lowest in older adults, with overlapping CrIs between pediatric and adult age groups otherwise.

Sensitivity and specificity adjustment had litle impact, as shown in exploratory analyses displayed in **Supplementary_Table_12** (sero-prevalence), and **Supplementary_Table_13** (period-specific SUARs).

## INTERPRETATION

Through eight cross-sectional sero-surveys conducted in the 2.5 years since the WHO declared the SARS-CoV-2 pandemic, we chronicle change in pediatric and adult sero-prevalence in the Lower Mainland, BC. During the first year of the pandemic, virus circulation was effectively suppressed through extraordinary public health measures, with virtually all remaining uninfected and immunologically-naïve. Vaccine availability and roll-out then radically transformed the immuno-epidemiological landscape, such that by September 2021, >80% overall had antibody evidence of priming to the virus with <15% having been infected. A subsequent series of Omicron waves then dramatically resulted in at least 60% overall having been infected by August 2022, including at least 70-80% of children ≤19 years, 60-70% of adults 20-59 years and ∼40% ≥60 years, culminating in >85% of children and >95% of adults having been primed (by vaccine and/or infection) overall. Individuals <50 years were especially affected by the Omicron BA.1 wave, notably serving as priming exposure for young children, while older adults ≥70 years experienced their greatest attack rates during subsequent BA.2 or BA.4/5 waves, superimposing infection-induced boost upon their high vaccine coverage.

Multiple immunological, epidemiological and/or modeling studies have reinforced the improved protection afforded by hybrid (vaccine + infection) immunity over that induced by either exposure alone[32-45]. In general, booster doses using ancestral Wuhan-like antigen among previously-infected individuals have resulted in transient increased protection with marginal incremental improvement against severe outcomes. Our age-related findings are consistent with the very young now being most-infected but least-vaccinated, while the very old are most-vaccinated and least-infected. Among elderly adults who contribute the greatest number of severe outcomes, more than half remain reliant on vaccine-induced immunity for their protection as we enter the fall of 2022. The extent to which prior vaccine/infection history should help guide booster-dose recommendations overall depends upon a number of factors, not the least of which because a large proportion may not be aware of their infection status[46].

Moreover, the antigenic relatedness and immunological interactions between previously-infecting viruses, original and updated vaccine strains and currently circulating or emerging variants are complex and dynamic. The incremental value of boosting by age further depends upon immunization program goals and individual and population risk assessments, notably in relation to the prevention of severe outcomes. What ultimately seems clear amidst this complexity is that the immuno-epidemiological context and sense of urgency to receive additional doses is now remarkably changed for most of the population, owing to the accumulation of more robust hybrid immunity.

Compared to other sero-surveillance in Canada or globally, our approach is unique in serially-sampling all pediatric and adult age groups and both sexes simultaneously, enabling their direct comparison and extending the information available from other more restricted population subsets (e.g. prenatal sera confined to women of childbearing age, or blood donors confined to mostly non-elderly adults). Overall, our findings align well with comparable subsets evaluated elsewhere. In the United States, comparison across several sero-surveys indicates similar age-related gradation in accumulated infection rates through June 2022, highest in children and lowest in older adults[47]. Among Canadian blood donors ≥18 years, about half had evidence of infection by the end of June 2022, highest among younger adults 18-25 years (∼70%) and lowest among elderly adults >65 years (∼30%)[48,49]. Similarly, among blood donors ≥17 years in the United Kingdom through mid-August (epi-week 33) 2022, ∼70% were infected overall, also highest among younger adults 17-29 years (>80%) and lowest in older adults 70-84 years (about half)[30]. A consistent finding across published sero-surveys is the lower levels of hybrid immunity among older adults.

Our sero-surveillance also indicates that standard case-based reporting under-estimated infections in the Lower Mainland by up to 100-fold between March and August 2022 alone, following the loosening of public health measures, the tightening of NAAT access and the abundance of non-reportable RAT use. While other surveillance indicators may be warranted, including those for which access to testing is more consistent (e.g. hospitalizations) or sustainable (e.g. wastewater sampling), the derivation of per case severe outcome risks will continue to require denominator case tallies. Ongoing sero-surveillance and associated SUAR estimation is important in informing the fold-increase to case denominators (and commensurate fold-decrease to per case severe outcome estimates) to inform accurate risk assessment.

Our study has limitations. Residual clinical specimens are more likely to come from people with underlying comorbidities who may differ in their exposure risk and immune response, together tending to under-estimate infection-induced sero-prevalence. Delay in generating an antibody response and uncertainty in its duration, each also tend toward under-estimation. Such issue may be suggested in our comparison with anti-S1 detection among mostly unvaccinated children <5 years for whom estimates should be comparable but were ∼10% lower for anti-NP detection. We otherwise show good concordance between our estimates of any sero-prevalence and vaccine coverage and between our participant and source population age/sex profiles. Our period-specific SUAR estimates assumed no re-infections, consistent with surveillance case reporting in BC, but we anticipate their exclusion to have had minimal impact. In analysis from Quebec, Canada, Carazo et al identified re-infections among just 4% of community-dwelling cases ≥12 years old during the Omicron BA.1 dominant period[34], and among just 8% of healthcare worker cases during the Omicron BA.2 dominant period[35].

Finally, the extrapolation of our findings to other areas should take into account context-specific differences such as in-person school attendance, implementation of mask mandates, vaccination program adjustments and other mitigation measures.

In conclusion, sero-prevalence findings presented here indicate almost complete reversal in the proportion of the population immunologically naïve versus primed against SARS-CoV-2 since its first emergence 2.5 years ago. Age-based sero-prevalence estimates best inform risk assessment and response in relation to public health goals. In that regard, most of the pediatric and adult population are at low risk of severe COVID-19 outcomes due in part to their accrued vaccine and infection exposures, creating more robust hybrid immunity. Conversely the elderly, still at greatest risk of severe outcomes, remain largely dependent upon vaccine-induced protection alone, and should remain the priority for additional doses.

## Data Availability

Aggregate data are available within the manuscript and the supplementary material. Any further data sharing will be considered upon reasonable request to the corresponding author with appropriate review and aggregation as required to comply with the provincial legislation under which the data were assembled and respecting privacy and confidentiality requirements.

## REFERENCES

1. Skowronski DM, Hottes TS, McElhaney JE, et al. Immuno-epidemiologic correlates of pandemic H1N1 surveillance observations: higher antibody and lower cell-mediated immune responses with advanced age. J Infect Dis, 2011;203:158–67.

2. Skowronski DM, Hottes TS, Janjua NZ, et al. Prevalence of seroprotection against the pandemic (H1N1) virus after the 2009 pandemic. CMAJ, 2010;182:1851–6.

3. Skowronski DM, Chambers C, Sabaiduc S, et al. Pre- and post-pandemic estimates of 2009 pandemic influenza A(H1N1) seroprotection to inform surveillance-based incidence, by age, during the 2013-2014 epidemic in Canada. J Infect Dis, 2015;211:109–14.

4. Van Kerkhove MD, Hirve S, Koukounari A, et al. Estimating age-specific cumulative incidence for the 2009 influenza pandemic: a meta-analysis of A(H1N1)pdm09 serological studies from 19 countries. Influenza Other Respir Viruses 2013;7:872–86.

5. Skowronski DM, Moser FS, Janjua NZ, et al. H3N2v and other influenza epidemic risk based on age-specific estimates of sero-protection and contact network interactions. PLoS One, 2013;8:e54015.

6. Skowronski DM, Janjua NZ, De Serres G, et al. Cross-reactive and vaccine-induced antibody to an emerging swine-origin variant of influenza A virus subtype H3N2 (H3N2v). J Infect Dis, 2012;206:1852–61.

7. Skowronski DM, Chambers C, Gustafson R, et al. Avian Influenza A(H7N9) Virus Infection in 2 Travelers Returning from China to Canada, January 2015. Emerg Infect Dis, 2016;22:71–4.

8. BC STATS. Population estimates. Victoria, BC: BC Ministry of Citizens’ Services, 2021. [Accessed 8 September 2022]. Available at: https://www2.gov.bc.ca/gov/content/data/statistics/people-population-community/population/population-projections

9. BC STATS. Population projections. (P.E.O.P.L.E) Victoria, BC: BC Ministry of Citizens’ Services, 2021. [Accessed 8 September 2022]. Available at: https://www2.gov.bc.ca/gov/content/data/statistics/people-population-community/population/population-projections

10. World Health Organization. WHO Director-General’s opening remarks at the media briefing on COVID-19 – 11 March 2020. Geneva, Switzerland: WHO. [Accessed 8 September 2022]. Available at: https://www.who.int/director-general/speeches/detail/who-director-general-s-opening-remarks-at-the-media-briefing-on-covid-1911-march-2020

11. British Columbia Centre for Disease Control. BC COVID-19 Data Trends. [Accessed 8 September 2022]. Available: http://www.bccdc.ca/health-info/diseases-conditions/covid-19/data-trends

12. Hogan CA, Jassem AN, Sbihi H, et al. Rapid increase in SARS-CoV-2 P.1 lineage leading to codominance with B.1.1.7 lineage, British Columbia, Canada, January-April 2021. Emerg Infect Dis 2021;27:2802–9.

13. National Advisory Committee on Immunization (NACI): Statements and publications. COVID-19. Ottawa: NACI. [Accessed 8 September 2022]. Available: https://www.canada.ca/en/public-health/services/immunization/national-advisory-committee-on-immunization-naci.html#covid-19

14. World Health Organization (WHO). Coronavirus disease (COVID-19) technical guidance.The Unity Studies: Early Investigations Protocols. Population-based age-stratified seroepidemiological investigation protocol for COVID-19 infection. [Accessed 8 September 2022]. Available at: https://www.who.int/emergencies/diseases/novel-coronavirus-2019/technical-guidance/early-investigations

15. Health Canada. Authorized medical devices for uses related to COVID-19: List of authorized testing devices. [Accessed 8 September 2022]. Available at: https://www.canada.ca/en/health-canada/services/drugs-health-products/covid19-industry/medical-devices/authorized/list.html.

16. United States Food and Drug Administration. EUA Authorized Serology Test Performance. [Accessed 8 September 2022]. Available at: https://www.fda.gov/medical-devices/emergency-situations-medical-devices/eua-authorized-serology-test-performance.

17. Mohanraj D, Whitelegg A, Bicknell K, Bhole M, Taylor L, Webber C. Comparative assessment of SARS-CoV-2 serology in healthcare workers with Abbott Architect, Roche Elecsys and The Binding site ELISA immunoassays. [Accessed 8 September 2022]. medRxiv, Preprint; not peer-reviewed. Available at: https://www.medrxiv.org/content/10.1101/2021.03.19.21253518v1. Posted 24 March 2021.

18. Tan SS, Saw S, Chew KL, et al. Comparative clinical evaluation of the Roche Elecsys and Abbott Severe Acute Respiratory Syndrome Coronavirus 2 (SARS-CoV-2) serology assays for Coronavirus Disease 2019 (COVID-19). Arch Pathol Lab Med 2021;145:32–38.

19. El-Khoury JM, Schulz WL, Durant TJS. Longitudinal assessment of SARS-CoV-2 antinucleocapsid and antispike-1-RBD antibody testing following PCR-detected SARS-CoV-2 infection. J Appl Lab Med 2021;6:1005–1011.

20. Deshpande GR, Kaduskar O, Deshpande K, et al. Longitudinal clinico-serological analysis of anti-nucelocapsid and anti-receptor binding domain of spike protein antibodies against SARS-CoV-2. Int J Infect Dis 2021;112:103–10.

21. Nakagama Y, Komase Y, Kaku N, et al. Detecting waning serological response with commercial immunoassays: 18-month longitudinal follow-up of anti-SARS-CoV-2 nucleocapsid antibodies. Microbiol Spectr 2022 July14;e0098622. doi: 10.1128/spectrum.00986-22. Online ahead of print.

22. Stone M, Grebe E, Sulaeman H, et al. Evaluation of commercially Available high-throughput SARS-CoV-2 serologic assays for serosurveillance and related applications. Emerg Infect Dis 2022;28:672–83.

23. Gelman A, Carpenter B. Bayesian analysis of tests with unknown specificity and sensitivity. Journal of the Royal Statistical Society: Series C (Applied Statistics) 2020;69: 1269–1283. doi: 10.1111/rssc.12435. [Accessed 8 September 2022]. Available: https://www.semanticscholar.org/paper/Bayesian-analysis-of-tests-with-unknown-specificity-Gelman-Carpenter/7aabc20ee9be5d4a7aa18b4135ceab1b648c87b9

24. Stringhini, Silvia, et al. Seroprevalence of anti-SARS-CoV-2 IgG antibodies in Geneva, Switzerland (SEROCoV-POP): a population-based study. The Lancet 2020;396:313–319.

25. Sekirov I, Barakauskas VE, Simons J, et al. SARS-CoV-2 serology: Validation of high-throughput chemiluminescent immunoassay (CLIA) platforms and a field study in British Columbia. J Clin Virol;142:104914. doi:10.1016/j.jcv.2021.104914. Epub 2021 Jul 16.

26. Bailie CR, Tseng Y-Y, Carolan L, Kirk MD, Nicholson S, Fox A, Sullivan SG. Trend in sensitivity of SARS-CoV-2 serology one year after mild and asymptomatic COVID-19: unpacking potential bias in seroprevalence studies. Clin Infect Dis 2022 Jan 13;ciac020. doi: 10.1093/cid/ciac020. Online ahead of print.

27. Elslande JV, Oyaert M, Ailliet S, et al. Longitudinal follow-up of IgG anti-nucleocapsid antibodies in SARS-VoV-2 infected patients up to eight months after infection. J Clin Virol 2021;136:104765. doi: 10.1016/j.jcv.2021.104765. Epub 2021 Feb 18.

28. Allen N, Brady M, Carrion MAI, et al. Serological markers of SARS-CoV-2 infection; anti-nucleocapsid antibody positivity may not be the ideal marker of natural infection in vaccinated individuals. J Infect. 2021;83:e9–e10. doi: 10.1016/j.jinf.2021.08.012. Epub 2021 Aug 9.

29. Victorian Government. Seroprevalence of SARS-CoV-2 specific antibodies among Victorian blood donors. Summary report for the Victorian Government Department of Health. State of Victoria, Australia. 03 May 2022. [Accessed 8 September 2022]. Available: https://www.health.vic.gov.au/research-and-reports/seroprevalence-of-sars-cov-2-specific-antibodies-among-victorian-blood-donors

30. United Kingdom Health Security Agency. COVID-19 vaccine surveillance report. Week 35. 1 September 2022. [Accessed 8 September 2022]. Available: https://assets.publishing.service.gov.uk/government/uploads/system/uploads/attachment_data/file/1101870/vaccine-surveillance-report-week-35.pdf

31. Downes M, Gurrin LC, English DR, Pirkis J, Currier D, Spittal MJ, Carlin JB. Multilevel regression and poststratification: A modeling approach to estimating population quantities from highly selected survey samples. American J Epidemiol 2018;187:1780–90.

32. Khan K, Karim F, Ganga Y, et al. Omicron BA.4/BA.5 escape neutralizing immunity elicited by BA.1 infection. Nat Commun 2022 Aug 10;13(1):4686. doi:10.1038/s41467-022-32396-9.

33. Hachmann NP, Miller J, Collier AY, et al. Neutralization escape by SARS-CoV-2 subvariants BA.2.12.1, BA.4, and BA.5. N Engl J Med 2022;387:86–88.

34. Carazo S, Skowronski DM, Brisson M, et al. Protection against Omicron re-infection conferred by prior heterologous SARS-CoV-2 infection, with and without mRNA vaccination. [in press] medRxiv pre-print. May 3, 2022. [Accessed 8 September 2022]. Available: https://www.medrxiv.org/content/10.1101/2022.04.29.22274455v2

35. Carazo S, Skowronski DM, Brisson M, et al. Protection against Omicron BA.2 reinfection conferred by primary Omicron or pre-Omicron infection with and without mRNA vaccination: a test-negative case-control study among healthcare workers. Lancet Infec Dis; 2022 [in press].

36. Altarawneh HN, Chemaitelly H, Ayoub HH, et al. Effects of previous infection and vaccination on symptomatic Omicron infections. N Engl J Med 2022;387:21–34.

37. Shrestha NK, Burke PC, Nowacki AS, Terpeluk P, Gordon SM. Necessity of coronavirus disease 2019 (COVID-19) vaccination in persons who have already had COVID-19. Clin Infec Dis 2022;75:e662–71.

38. Hall V, Foulkes S, Insalata F, et al Protection against SARS-CoV-2 after Covid-19 vaccination and previous infection. NEJM 2022;386:1207–20.

39. Hammerman A, Sergienko R, Friger M, et al. Effectiveness of the BNT162b2 vaccine after recovery from Covid-19. N Engl J Med;386:1221–29.

40. Plumb ID, Feldstein LR, Barkley E, et al. Effectiveness of COVID-19 mRNA vaccination in preventing COVID-19-associated hospitalization among adults with previous SARS-CoV-2 infection – United States, June 2021-February 2022. MMWR Morb Mortal Wkly Rep 202271:549–55.

41. Lind ML, Robertson AJ, Silva J, et al. Effectiveness of primary and booster COVID-19 mRNA vaccination against Omicron variant SARS-CoV-2 infection in people with a prior SARS-CoV-2 infection. medRxiv pre-print. April 25, 2022. [Accessed 5 September 2022]. Available: https://www.medrxiv.org/content/10.1101/2022.04.19.22274056v3

42. Goldberg Y, Mandel M, Bar-On YM, et al. Protection and waning of natural and hybrid immunity to SARS-CoV-2. N Engl J Med 2022;386:2201–12.

43. Chin ET, Leidner D, Lamson L, et al. Protection against Omicron conferred by mRNA primary vaccination series, boosters, and prior infection. medRxiv pre-print. May 27, 2022. [Accessed 5 September 2022]. Available: https://www.medrxiv.org/content/10.1101/2022.05.26.22275639v1

44. Cerqueira-Silva T, Oliveira VA, Paixao ES, et al. Vaccination plus previous infection: protection during the Omicron wave in Brazil. Lancet Infect Dis 2022;22:945–6.

45. Khoury DS, Docken SS, Subbarao K, Kent SJ, Davenport MP, Cromer D. Predicting the efficacy of variant-modified COVID-19 vaccine boosters. medRxiv pre-print. August 25, 2022. [Accessed 5 September 2022]. Available: https://www.medrxiv.org/content/10.1101/2022.08.25.22279237v1

46. Joung SY, Ebinger JE, Sun N, et al. Awareness of SARS-CoV-2 Omicron variant infection among adults with recent COVID-19 seropositivity. JAMA Netw Open. 2022 Aug 1;5(8):e2227241. doi: 10.1001/jamanetworkopen.2022.27241.

47. National Cancer Institute, National Institute of Allergy and Infectious Diseases, Centers for Disease Control and Prevention. COVID-19 SeroHub. 2022. [Accessed 5 September 2022]. Available: https://covid19serohub.nih.gov

48. Canadian Blood Services. COVID-19 seroprevalence report. Report #23: June 2022 survey. The advance of Omicron. August 3rd, 2022. [Accessed 5 September 2022]. Available: https://www.covid19immunitytaskforce.ca/wp-content/uploads/2022/08/covid-19-full-report-june-2022.pdf

49. COVID-19 Immunity Task Force. Young adults remain the primary vector of SARS-CoV-2 transmission: latest data from Hema-Quebec. August 8, 2022. [Accessed 5 September 2022]. Available: https://www.covid19immunitytaskforce.ca/young-adults-remain-the-primary-vector-of-sars-cov-2-transmission-latest-data-from-hema-quebec/

