## Supplementary_Material for "Serial cross-sectional estimation of vaccine and infection-induced SARS-CoV-2 sero-prevalence in children and adults, British Columbia, Canada: March 2020 to August 2022"

**Supplementary Material 1.** Public health context for sero-surveys in Lower Mainland, British Columbia (BC), Canada

Testing and surveillance reporting

Publicly-funded access to nucleic acid amplification testing (NAAT) in British Columbia (BC) was initially exposure-based (until March 15, 2020), or targeted (until April 8, 2020) with an expanded but still targeted indication until April 20, 2020 (epi-week 17). Thereafter NAAT testing was broadly available for symptomatic individuals from April 21, 2020 (with revision of qualifying symptoms on December 17, 2020) through to January 18, 2022 at which time community access to NAAT testing was again limited to high-risk individuals (e.g. unvaccinated, immuno-compromised, or those who live/work in certain high risk settings) owing to laboratory capacity during the Omicron wave [1]. Shortly thereafter also, publicly-funded access to rapid antigen tests (RATs) increased for community-dwelling adults through pharmacies and for children through schools. NAAT (but not RAT) confirmed or epidemiologically-linked cases are reportable under the Public Health Act to the BC Centre for Disease Control (BCCDC), under delegation of the Provincial Health Officer [1]. Through the current study period spanning to August 2022, BCCDC surveillance reports capture only the first reported case, and exclude re-infections [1]. Although not available in BC, in analyses of hybrid (vaccine + infection-induced immunity) in Quebec, Canada, Carazo et al identified re-infections (≥90 days after a prior primary infection) among just 4% (9,505/224,007) of community-dwelling cases ≥12 years of age during the Omicron BA.1 dominant period spanning December 26, 2021 (epi-week 52) to March 12, 2022 (epi-week 10) [2], and among just 8% (2,991/37,543) of healthcare worker cases during the Omicron BA.2 dominant period spanning March 27 to June 4, 2022 (epi-weeks 13-22) [3].

Population-level public health measures

Population-level, public health measures began in BC in mid-March 2020 [4] and were variously relaxed and reinforced thereafter in response to epidemic waves and public health need. This includes "core bubble" social restrictions implemented in November 2020 (epi-week 45) in response to the second wave [5,6], or "circuit breaker" social restrictions in March 2021 (epi-week 13) during the third (Alpha/Gamma) wave [7], followed by a "restart plan" beginning in May 2021 (epi-week 21) that was disrupted by the start of the fourth (Delta) wave that peaked in the fall of 2021. Apart from temporary school closure between March 17 and June 1 (epi-weeks 12-22) of 2020, children were able to attend scheduled classes in person throughout the pandemic, with on-line options also available [4]. Masking within indoor public settings was mandated for all individuals ≥12 years old (except in schools) beginning November 2020 (epi-week 47) at the peak of the second wave, continuing through the third wave that peaked in spring 2021. With a brief summer pause beginning in July (epi-week 26) and ending in August (epi-week 34) of 2021, mandatory masking continued and was extended to the school setting for the 2021-22 academic year, including children ≥5 years from October 2021 (epi-week 43), during the fourth wave. With Omicron (BA.1) ultimately displacing Delta in December 2021 [1], BC experienced its most intense fifth wave, prompting more stringent "holiday season" public health measures beginning epi-week 51 of 2021 that were lifted in February 2022 (epi-week 7) along with mask mandates in March 2022 (epi-week 10). Thereafter, BC experienced sixth and seventh waves due to Omicron (predominantly BA.2 and BA.5, respectively) that were associated with lesser peaks based on surveillance case reports [1].

<u>Vaccine roll-out</u>

Two spike (S1)-based mRNA vaccines were first authorized in Canada in mid-December 2020 [8,9]. Vaccination in BC was initially targeted to long-term care residents, healthcare workers and clinically extremely vulnerable people, with age-based prioritization of the oldest community-dwelling adults beginning in March 2020. Adolescents ≥12 years old became eligible in May 2021 and children 5-11 years old in November 2021 followed by children 0-4 years from August 2, 2022 [8]. Booster dose access followed a similar prioritization sequence. A single-dose vaccine card for entry into social/recreational settings was introduced for all ≥12 years old in September 2021 (epi-week 37) and a two-dose card in October 2021 (epi-week 43), ultimately repealed in April 2022 (epi-week 14). Provincial vaccine coverage estimates (approximated from provincial immunization registry tallies and population census estimates) suggest that by the epi-week 2 start of the 4th sero-survey in January 2021, not more than ~1% of the BC population overall had received a first dose; whereas, by the epi-week 22 start of the 5th sero-survey in May/June 2021 this had increased to ~60% (including >90% ≥70 years (including care facility residents), about three-quarters 50-69 years, 60% 18-49 years and nearly 15% 12-17 years). By the epi-week 39 start of the 6th sero-survey in September 2021, nearly 80% of British Columbians overall were vaccinated (including nearly three-quarters of those 12-17 years) and nearly three-quarters overall were twice vaccinated. By the epi-week 11 start of the 7th sero-survey in March 2022 about half of children 5-11 years and 85% of adolescents 12-17 years had been vaccinated with about half the population overall having received a third dose. By the epi-week 31 start of the 8th sero-survey in July/August 2022, <1% of children 0-4 years, half 5-11 years, and nearly 90% of teens, respectively, had been vaccinated with ~10% of the population overall having received four doses, including nearly 60% of those ≥70 years.

### References for Supplementary Material 1

1. British Columbia Centre for Disease Control. BC COVID-19 Data Trends. [Accessed 28 August 2022]. Available: http://www.bccdc.ca/health-info/diseases-conditions/covid-19/data-trends
2. Carazo S, Skowronski DM, Brisson M, et al. Protection against Omicron re-infection conferred by prior heterologous SARS-CoV-2 infection, with and without mRNA vaccination. [in press] medRxiv pre-print. May 3, 2022. [Accessed 24 August 2022]. Available: https://www.medrxiv.org/content/10.1101/2022.04.29.22274455v2
3. Carazo S, Skowronski DM, Brisson M, et al. Protection against Omicron BA.2 reinfection conferred by primary Omicron or pre-Omicron infection with and without mRNA vaccination: a test-negative case-control study among healthcare workers. Lancet Infec Dis; 2022 [in press].
4. CTV News. Scroll through this timeline of the 1st year of COVID-19 in BC. Vancouver News. Thursday January 28, 2021 [Accessed 8 September 2022]. Available at: https://bc.ctvnews.ca/scroll-through-this-timeline-of-the-1st-year-of-covid-19-in-b-c-1.5284929
5. BC Government News. Joint statement on BC's COVID-19 response, latest updates. Saturday November 7, 2020 [Accessed 8 September 2022]. Available: https://news.gov.bc.ca/releases/2020HLTH0059-001922
6. BC Government News. Joint statement on BC's COVID-19 response, latest updates. Thursday November 19, 2020 [Accessed 8 September 2022]. Available: https://news.gov.bc.ca/releases/2020HLTH0061-001949
7. BC Government News. Office of the Premier. Three-week circuit breaker begins now to bend the curve, protect people. Monday March 29, 2021. [Accessed 8 September 2022]. Available: https://news.gov.bc.ca/releases/2021PREM0023-000578
8. National Advisory Committee on Immunization (NACI): Statements and publications. COVID-19. Ottawa: NACI. [Accessed 8 September 2022]. Available: https://www.canada.ca/en/public-health/services/immunization/national-advisory-committee-on-immunization-naci.html#covid-19
9. Skowronski DM, Febriani Y, Ouakki M, et al. Two-dose SARS-CoV-2 vaccine effectiveness with mixed schedules and extended dosing intervals: test-negative design studies from British Columbia and Quebec, Canada. Clin Infect Dis 2022 Apr 19:ciac290. Doi: 10.1093/cid/ciac290. Online ahead of print [Supplementary Material 1 provides additional details related to vaccine program modifications and roll-out]

**Supplementary Figure 1.** Sampling area and eligible municipalities

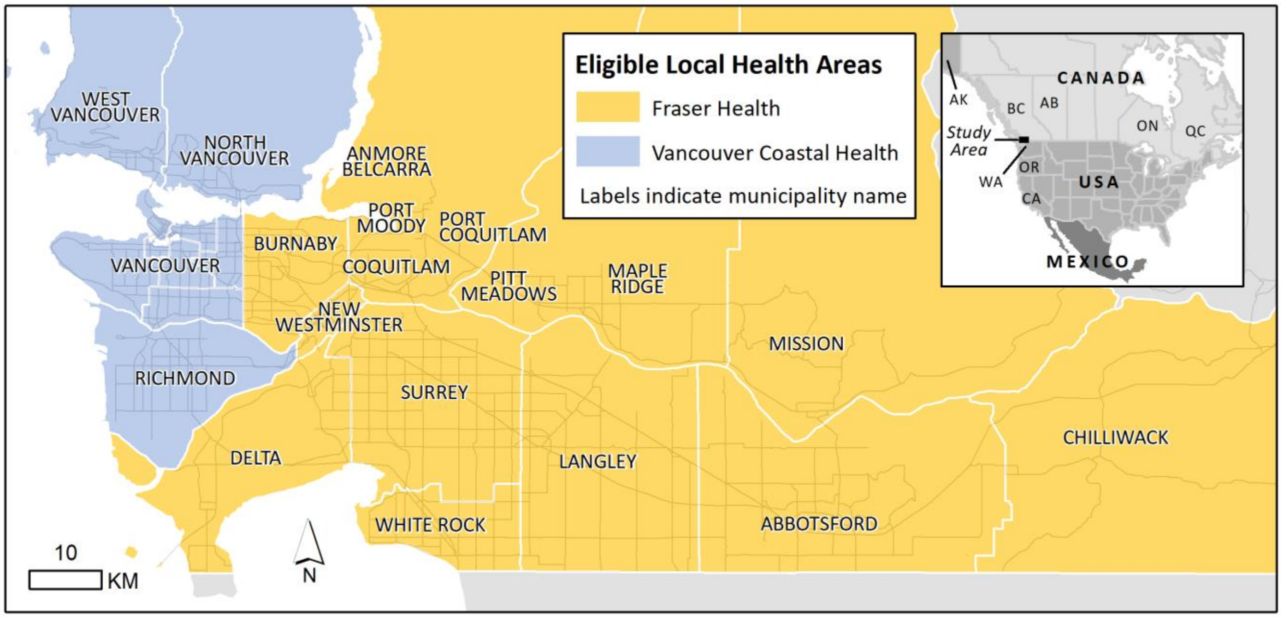

Abbreviations: AB = Alberta; AK = Alaska; BC = British Columbia; CA = California; ON = Ontario; OR = Oregon; QC = Quebec; USA = United States of America; WA = Washington

The sero-survey protocol utilizes anonymized residual sera collected from individuals of all ages presenting for outpatient blood work in the Lower Mainland (Greater Vancouver Area and Fraser Valley) of BC, Canada, where 60% (~3 million) of the total provincial population (~5 million) resides.[a,b]

Two health authorities (HA) are responsible for health administration and surveillance reporting within the Lower Mainland of BC including Fraser Health Authority (FHA: ~1.9 million) and Vancouver Coastal Health Authority (VCHA: ~1.2 million).[a,b] Eligible local health authorities of the Lower Mainland are displayed with their associated municipality names and with those belonging to FHA shaded yellow and those belonging to VCHA shaded blue.

[a] BC STATS. Population estimates. Victoria, BC: BC Ministry of Citizens' Services, 2021. [Accessed 8 September 2022]. Available at: https://www2.gov.bc.ca/gov/content/data/statistics/people-population-community/population/population-projections

[b] BC STATS. Population projections. (P.E.O.P.L.E) Victoria, BC: BC Ministry of Citizens' Services, 2021. [Accessed 8 September 2022]. Available at: https://www2.gov.bc.ca/gov/content/data/statistics/people-population-community/population/population-projections

**Supplementary Material 2.** Sensitivity and specificity considerations, by sero-survey

Five chemiluminescent immunoassays (CLIAs) were used in the current sero-survey including Ortho-S1[a], Abbott-NP[b], Siemens-S1-RBD[c], Roche-NP[d], and Abbott-S1-RBD[e]. The manufacturers of these CLIAs each reported high sensitivities (>98%) and specificities (>99.5%) but with minimal participant details provided [1,2]. Assay characteristics may vary by population sub-group characteristics such as vaccine status, time since exposure, disease severity, age group etc. but most validation studies to date have not addressed these issues. Like other public health agencies more recently, we did not adjust for sensitivity or specificity in primary sero-prevalence analyses [3-7]. In exploratory analyses we incorporated sensitivity and specificity adjustment assuming values (positive and negative controls) foremost informed by empirical field investigation by the BC Centre for Disease Control (BCCDC) Public Health Laboratory (PHL) [8]. These are summarized in the table immediately below with additional rationale and references detailed in subsequent sections of this Supplement. We recognize these attempts to adjust for sensitivity and specificity are imperfect and incomplete and further complicated by dual-assay interpretation and variation by population subgroups. Cautious interpretation is warranted.

| **Sero-prevalence estimate by sero-survey** | **Assumed Sensitivity values n/N; % (95%CI) [f]** | **Assumed Specificity values n/N; % (95%CI) [f]** | **Relevant assays included in dual-assay interpretation** [see detailed rationale in sections below this table] |
|---|---|---|---|
| **Any vaccine- and/or infection-induced dual-assay sero-positivity** | | | |
| Sero-surveys 1-5 | 92/92<br>100% (96.1, 100) | 188/189<br>99.5% (97.1, 100) | (Ortho-S1 + Abbott-NP) OR (Abbott-NP + Siemens-S1-RBD) OR (Ortho-S1 + Siemens-S1-RBD) |
| Sero-surveys 6-8 | 92/92<br>100% (96.1, 100) | 187/189<br>98.9% (96.2, 99.9) | (Ortho-S1/Abbott-S1-RBD + Roche-NP) OR (Roche-NP + Siemens-S1-RBD) OR (Ortho-S1/Abbott-S1-RBD + Siemens-S1-RBD) [g] |
| **Infection-induced (with/without vaccination) dual-assay sero-positivity** | | | |
| Sero-surveys 1-3 [h] | 92/92<br>100% (96.1, 100) | 188/189<br>99.5% (97.1, 100) | (Ortho-S1 + Abbott-NP) OR (Abbott-NP + Siemens-S1-RBD) OR (Ortho-S1 + Siemens-S1-RBD) [h] |
| Sero-surveys 4-5 | 88/92<br>95.7% (89.2, 98.8) | 189/189<br>100% (98.1, 100) | (Ortho-S1 + Abbott-NP) OR (Abbott-NP + Siemens-S1-RBD) [i] |
| Sero-surveys 6-7 | 91/92<br>98.9% (94.1, 100) | 187/189<br>98.9% (96.2, 99.9) | (Ortho-S1 + Roche-NP) OR (Roche-NP + Siemens-S1-RBD) |
| Sero-survey 8 [j] | 91/92<br>98.9% (94.1, 100) | 187/189<br>98.9% (96.2, 99.9) | (Abbott-S1-RBD [i] + Roche-NP) OR (Roche-NP + Siemens-S1-RBD) |

[a] Ortho assay detects total antibody (IgA, IgG and IgM) to recombinant spike (S1) using the Vitros XT 7600 analyzer (Ortho-Clinical Diagnostics, Rochester, New York).
[b] Abbott assay detects IgG antibody to nucleocapsid (NP) using the ARCHITECT i2000SR analyzer (Abbott Laboratories, Diagnostic Division, Abbott Park, Illinois).
[c] Siemens assay detects total (IgG, IgM) antibodies to the S1 receptor-binding domain (S1-RBD) by the ADVIA Centaur XPT system (Siemens Healthineers, Erlangen, Germany).
[d] Roche assay detects total antibody (IgA, IgG and IgM) to nucleocapsid (NP) using the Roche cobas e601 analyzer (Roche Diagnostics Gmbh, Mannheim, Germany).
[e] Abbott assay detects IgG to the spike receptor binding domain (S1-RBD) using the ARCHITECT i2000SR analyzer (Abbott Laboratories, Diagnostic Division, Abbott Park, Illinois).
[f] Exact confidence interval (CI). One sided 97.5% confidence intervals for cells with zero or 100% counts.
[g] Assumes comparable orthogonal interpretation of any (vaccine- or infection-induced) dual-assay sero-positivity whether predicated on Ortho-S1/Roche-NP/Siemens-S1-RBD OR Abbott-S1-RBD/Roche-NP/Siemens-S1-RBD.
[h] All dual-assay sero-positivity during the first three sero-surveys before vaccine roll-out assumed to be infection induced.
[i] Disregards supplemental Roche-NP testing undertaken during these sero-surveys.
[j] Assumes comparable orthogonal interpretation of infection-induced dual-assay sero-positivity whether using Ortho-S1 or Abbott-S1-RBD in combination with Roche-NP.

Rationale and references for assumed sensitivity and specificity values

The BCCDC PHL reported assay characteristics from an internal validation (n=97) and facility-based outbreak field investigation (n=281) [8]. We predicate our sensitivity and specificity assumptions on the larger field investigation. The latter included 92 SARS-CoV-2 cases, laboratory-confirmed by nucleic acid amplification test (NAAT), with sera collected >14 days to two months post-onset between March-May 2020 and 189 negative controls. Three-quarters of cases were female with median (range) of age 76.5 (20-102) years; 71% of test-negative controls were females with median (range) of age 57 (22-102) years. In that field investigation, sensitivity was highest for the Ortho-S1 and Roche-NP assays at 98.9% (95%CI: 94.1, 99.8) each (91/92 cases detected), followed by the Siemens-S1-RBD at 97.8% (95%CI: 92.4, 99.4) (90/92) and the Abbott-NP assay at 95.7% (95%CI: 89.4, 98.3) (88/92). Specificity was 97.9% (95%CI: 94.7, 99.2) for the Ortho-S1(185/189 controls correctly identified) but 98.4% (95%CI: 95.4, 99.5) for both the Abbott-NP and Roche-NP assays (186/189), and 98.9% (95%CI: 96.2, 99.7) for Siemens-S1-RBD (187/189) [8].

Applying the dual-assay requirements for sero-positivity from the current sero-survey to the BCCDC field investigation, sensitivity was improved for the detection of *any* (vaccine- or infection-induced) sero-prevalence at 100% (95% CI: 96.1, 100) (92/92) for both the Ortho-S1/Abbott-NP/Siemens-S1-RBD algorithm used in sero-surveys 1-5 and the Ortho-S1/Roche-NP/Siemens-S1-RBD algorithm used in sero-surveys 6-7, with comparable or higher specificities also of 99.5% (95%CI: 97.1, 100) (188/189) and 98.9% (95%CI: 96.2, 99.9) (187/189), respectively, for both algorithms. For infection-induced sero-positivity requiring that at least one of the two positive assays used in orthogonal interpretation include anti-NP detection, sensitivities were 95.7% (95%CI: 89.2, 98.8) using Abbott-NP (88/92) and 98.9% (95%CI: 94.1, 100) using Roche-NP (91/92). Specificities were 100% (95%CI: 98.1, 100) (189/189) and 98.9% (95%CI: 96.2, 99.9) (187/189), respectively.

Although the Abbott-S1-RBD was not included in the above BCCDC PHL field investigation, in side-by-side comparison to which the BCCDC PHL contributed with multiple other laboratories [9], Abbott-S1-RBD sensitivity ranged 95.3% to 98.1% (depending on the reference definition) while sensitivity of the Ortho-S1 assay ranged 95.8% to 98.7%, with specificities of 99.5% and 100%, respectively [9]. As per other recent reports [10-14], sensitivity of the Roche-NP assay tended higher than that of Abbott-NP (95.8-98.7% versus 88.5-90.9%, respectively) with more comparable specificities (99.5% and 100%, respectively) [9]. To assess the impact of our having replaced the Ortho-S1 with the Abbott-S1-RBD assay at the 8$^{th}$ (July/August 2022) sero-survey, we compared findings among a random selection of 200 sera from the 7$^{th}$ (March 2022) sero-survey (54% female, median age 47 years, interquartile range 26-69 years) subjected to Abbott-S1-RBD in addition to the

Ortho-S1 assay. As shown in the table below, use of the Abbott-S1-RBD individually or in dual-assay interpretation was associated with only slightly lower sensitivity for any sero-positivity, but this did not affect orthogonal interpretation of infection-induced sero-positivity.

| **Comparing 8th sero-survey using Abbott-S1-RBD vs. 7th sero-survey using Ortho-S1 (N=200 randomly selected 7th sero-survey sera)** | **n/N % sensitivity (95% CI) [a]** | **n/N % specificity (95% CI) [a]** |
|---|---|---|
| **Relative to Ortho-S1 findings at 7th sero-survey [N=200]** | | |
| Abbott-S1-RBD assay findings | 186/188<br>98.9 (96.2, 99.9) | 12/12<br>100 (75.3, 100) |
| **Relative to 7th sero-survey findings of any dual-assay positivity: (Ortho-S1 + Siemens-S1-RBD) OR (Ortho-S1 + Roche-NP) OR (Siemens-S1-RBD + Roche-NP) [N=200]** | | |
| 8th sero-survey orthogonal approach of any dual positivity with:<br>(Abbott-S1-RBD + Siemens-S1-RBD) OR<br>(Abbott-S1-RBD[a] + Roche-NP) OR<br>(Siemens-S1-RBD + Roche-NP) | 186/187<br>99.5 (97.1, 100) | 13/13<br>100 (75.3, 100) |
| **Relative to 7th sero-survey orthogonal findings of dual-assay positivity inclusive of anti-NP detection: (Ortho-S1 + Roche-NP) OR (Siemens-S1 + Roche-NP) [N=80[b]]** | | |
| 8th sero-survey orthogonal approach of any dual positivity with at least one assay anti-NP positive:<br>(Abbott-S1-RBD + Roche-NP) OR (Siemens-S1-RBD + Roche-NP) | 67/67<br>100 (94.6, 100) | 13/13<br>100 (75.3, 100) |

We also assessed the impact of having switched from the Ortho-S1 to the Abbott-S1-RBD assay among a separate immunity study cohort of 37 adults (59% female, median age 40 years, interquartile range 33-53 years) who were NAAT-confirmed cases between March and June 2020 [unpublished study led by Skowronski/Sadarangani, reviewed and approved by the University of British Columbia Research Ethics Board]. As shown in the table below sensitivity of each of the assays was higher at 30 days versus 14 days post-onset and lowest at both time points for anti-NP detection whether by Abbott-NP or Roche-NP assays. Consistent with findings above, Abbott-S1-RBD sensitivity was marginally reduced compared to Ortho-S1, notably at 14 days post-onset, but with minimal impact on dual-assay interpretation of any or infection-induced sero-positivity, notably at 30 days post-onset.

[a] Exact confidence interval (CI). One sided 97.5% confidence intervals for cells with zero or 100% counts.
[b] Among these 80 participants, 54% female, median age 29 years, interquartile range 12-50 years.

| **Immunity study: Individual assays and dual-assay combinations used in current sero-survey** | **Sensitivity relative to NAAT Sera collected 14 days post-onset** | | | **Sensitivity relative to NAAT Sera collected 30 days post-onset** | | |
|---|---|---|---|---|---|---|
| | **Assay positive** | **NAAT positive** | **% Assay Sensitivity (95% CI) [a]** | **Assay positive** | **NAAT positive** | **% Assay Sensitivity (95% CI) [a]** |
| Ortho-S1 assay | 36 | 36 | 100 (90.3, 100) | 37 | 37 | 100 (90.51, 100) |
| Abbott-NP assay | 29 | 36 | 80.6 (64.0, 91.8) | 32 | 37 | 86.49 (71.2, 95.46) |
| Siemens-S1-RBD assay | 26 | 36 | 72.2 (54.8, 85.8) | 36 | 37 | 97.3 (85.8, 99.9) |
| **Orthogonal infection-induced sero-positivity [as per sero-surveys 1-5]:** (Ortho-S1 + Abbott-NP) OR (Abbott-NP + Siemens-S1-RBD) | 29 | 36 | 80.6 (64.0, 91.8) | 32 | 37 | 86.49 (71.2, 95.46) |
| **Any dual-assay sero-positivity [as per sero-surveys 1-5]:** (Ortho-S1 + Abbott-NP) OR (Ortho-S1 + Siemens-S1-RBD) OR (Abbott-NP + Siemens-S1-RBD) | 33 | 36 | 91.7 (77.53, 98.3) | 36 | 37 | 97.3 (85.8, 99.9) |
| Roche-NP assay | 28 | 36 | 77.8 (60.9, 89.9) | 32 | 37 | 86.49 (71.2, 95.46) |
| **Orthogonal infection-induced sero-positivity [as per sero-surveys 6-7]:** (Ortho-S1 + Roche-NP) OR (Roche-NP + Siemens-S1-RBD) | 28 | 36 | 77.8 (60.9, 89.9) | 32 | 37 | 86.49 (71.2, 95.46) |
| **Any dual-assay sero-positivity [as per sero-surveys 6-7]:** (Ortho-S1 + Roche-NP) OR (Ortho-S1 + Siemens-S1-RBD) OR (Roche-NP + Siemens-S1-RBD) | 33 | 36 | 91.7 (77.53, 98.3) | 36 | 37 | 97.3 (85.8, 99.9) |
| Abbott-S1-RBD | 33 | 36 | 91.7 (77.53, 98.3) | 36 | 37 | 97.3 (85.8, 99.9) |
| **Orthogonal infection-induced sero-positivity [as per sero-survey 8]:** (Abbott-S1-RBD + Roche-NP) OR (Roche-NP + Siemens-S1-RBD) | 26 | 36 | 72.2 (54.8, 85.8) | 32 | 37 | 86.49 (71.2, 95.46) |
| **Any dual-assay sero-positivity [as per sero-survey 8]:** (Abbott-S1-RBD + Roche-NP) OR (Abbott-S1-RBD + Siemens-S1-RBD) OR (Roche-NP + Siemens-S1-RBD) | 31 | 36 | 86.1 (70.50, 95.3) | 36 | 37 | 97.3 (85.8, 99.9) |

CI = Confidence interval (exact). One sided 97.5% confidence intervals for cells with zero or 100% counts; NAAT = nucleic acid amplification test

Overall, our dual-assay approach for any or infection-induced sero-prevalence estimation tended to improve upon both sensitivity and specificity compared to single assay detection. This suggests that adjustment for sensitivity/specificity may be less critical. Recognizing other potential variability and uncertainty in assay characteristics by population sub-groups, our sensitivity and specificity adjusted estimates presented in **Supplementary Table 12** **and** **Supplementary Table 13** are for exploratory interest only.

### References, Supplementary Material 2

1. Health Canada. Authorized medical devices for uses related to COVID-19: List of authorized testing devices. [Accessed 8 September 2022]. Available at: https://www.canada.ca/en/health-canada/services/drugs-health-products/covid19-industry/medical-devices/authorized/list.html.
2. United States Food and Drug Administration. EUA Authorized Serology Test Performance. [Accessed 8 September 2022]. Available at: https://www.fda.gov/medical-devices/emergency-situations-medical-devices/eua-authorized-serology-test-performance
3. Bailie CR, Tseng Y-Y, Carolan L, Kirk MD, Nicholson S, Fox A, Sullivan SG. Trend in sensitivity of SARS-CoV-2 serology one year after mild and asymptomatic COVID-19: unpacking potential bias in seroprevalence studies. Clin Infect Dis 2022 Jan 13;ciac020. Doi: 10.1093/cid/ciac020. Online ahead of print.
4. Elslande JV, Oyaert M, Ailliet S, et al. Longitudinal follow-up of IgG anti-nucleocapsid antibodies in SARS-VoV-2 infected patients up to eight months after infection. J Clin Virol 2021;136:104765. Doi: 10.1016/j.jcv.2021.104765. Epub 2021 Feb 18.
5. Allen N, Brady M, Carrion MAI, et al. Serological markers of SARS-CoV-2 infection; anti-nucleocapsid antibody positivity may not be the ideal marker of natural infection in vaccinated individuals. J Infect. 2021;83:e9-e10. Doi: 10.1016/j.jinf.2021.08.012. Epub 2021 Aug 9.
6. Victorian Government. Seroprevalence of SARS-CoV-2 specific antibodies among Victorian blood donors. Summary report for the Victorian Government Department of Health. State of Victoria, Australia. 03 May 2022. [Accessed 8 September 2022]. Available: https://www.health.vic.gov.au/research-and-reports/seroprevalence-of-sars-cov-2-specific-antibodies-among-victorian-blood-donors
7. United Kingdom Health Security Agency. COVID-19 vaccine surveillance report. Week 35. 1 September 2022. [Accessed 8 September 2022]. Available: https://assets.publishing.service.gov.uk/government/uploads/system/uploads/attachment_data/file/1101870/vaccine-surveillance-report-week-35.pdf
8. Sekirov I, Barakauskas VE, Simons J, et al. SARS-CoV-2 serology: Validation of high-throughput chemiluminescent immunoassay (CLIA) platforms and a field study in British Columbia. J Clin Virol;142:104914. Doi:10.1016/j.jcv.2021.104914. Epub 2021 Jul 16.
9. Stone M, Grebe E, Sulaeman H, et al. Evaluation of commercially-available high-throughput SARS-CoV-2 serologic assays for serosurveillance and related applications. Emerg Infect Dis 2022;28:672-83.
10. Mohanraj D, Whitelegg A, Bicknell K, Bhole M, Taylor L, Webber C. Comparative assessment of SARS-CoV-2 serology in healthcare workers with Abbott Architect, Roche Elecsys and The Binding site ELISA immunoassays. medRxiv, doi: https://doi.org/10.1101/2021.03.19.21253518, Preprint; not peer-reviewed. [Accessed 8 September 2022]. Available at: https://www.medrxiv.org/content/10.1101/2021.03.19.21253518v1 . Posted 24 March 2021.
11. Tan SS, Saw S, Chew KL, et al. Comparative clinical evaluation of the Roche Elecsys and Abbott Severe Acute Respiratory Syndrome Coronavirus 2 (SARS-CoV-2) serology assays for Coronavirus Disease 2019 (COVID-19). Arch Pathol Lab Med 2021;145:32-38.
12. El-Khoury JM, Schulz WL, Durant TJS. Longitudinal assessment of SARS-CoV-2 antinucleocapsid and antispike-1-RBD antibody testing following PCR-detected SARS-CoV-2 infection. J Appl Lab Med 2021;6:1005-1011.
13. Deshpande GR, Kaduskar O, Deshpande K, et al. Longitudinal clinico-serological analysis of anti-nucelocapsid and anti-receptor binding domain of spike protein antibodies against SARS-CoV-2. Int J Infect Dis 2021;112:103-10.
14. Nakagama Y, Komase Y, Kaku N, et al. Detecting waning serological response with commercial immunoassays: 18-month longitudinal follow-up of anti-SARS-CoV-2 nucleocapsid antibodies. Microbiol Spectr 2022 July14;e0098622. Doi: 10.1128/spectrum.00986-22. Online ahead of print.

**Supplementary Material 3.** Derivation of surveillance under-ascertainment ratios (SUARs)

Surveillance under-ascertainment ratios (SUARs) with 95% credible intervals (CrIs) were derived by dividing the estimated number of infections by the number of cases reported from the Lower Mainland, British Columbia (BC). Infection-induced sero-prevalence estimates (SP; age-, sex- and health authority (HA)-standardized) were multiplied by the Fraser Health Authority (FHA) and Vancouver Coastal Health Authority (VCHA) population census estimates to derive infection rates [1,2], and surveillance case tallies are those reported by FHA and VCHA to the BC Centre for Disease Control (BCCDC).

Surveillance case reports reflect tallies of laboratory-confirmed, laboratory probable and epidemiologically-linked cases as of the August 2022 sero-survey but exclude cases from out-of-province and care facility (long term care, assisted living, independent living) residents to reflect community-dwelling Lower Mainland residents. Surveillance case reports were timed by episode date defined hierarchically by onset date, or if unavailable then specimen collection or laboratory result date. Taking into account a 10-14-day span from the first to last serum collection and comparable lag to antibody development, for each sero-survey we identified as referent the date two weeks earlier than the last serum collection date. We then summed surveillance case reports with episode date to the end of the corresponding complete epidemiological week. By way of example, for the 8$^{th}$ sero-survey spanning serum collection to August 11, 2022 we identified July 28, 2022 as referent date and summed surveillance case reports with episode date to the end of epi-week 30.

Through the current study period spanning to August 2022, official BCCDC surveillance case report tallies include only the first (primary infection) and exclude re-infections [3]. Although not available in BC, in analyses of hybrid (vaccine + infection-induced) immunity in Quebec, Canada, Carazo et al identified re-infections (≥90 days after a prior primary infection) among just 4% (9,505/224,007) of community-dwelling cases ≥12 years of age during the Omicron BA.1 dominant period spanning December 26, 2021 (epi-week 52) to March 12, 2022 (epi-week 10) [4], and among just 8% (2,991/37,543) of healthcare worker cases during the Omicron BA.2 dominant period spanning March 27 to June 4, 2022 (epi-weeks 13-22) [5]. On that basis we anticipate the exclusion of re-infections to have had minimal impact on SUAR estimates.

Cumulative SUARs by the 4$^{th}$ to 8$^{th}$ sero-surveys (January 2021, May/June 2021, September/October 2021, March 2022 and July/August 2022) were derived overall based on cumulative sero-prevalence-estimated infection rates and surveillance case reports. The sero-prevalence rate ($p_{ijkl}$) for each survey period $i$, age group $j$, sex $k$, and health region $l$ were drawn from the posterior distribution and combined with the estimated population size ($n_{jkl}$) to produce the binomially-distributed number of cumulative infections for cell $i, j, k, l$:

$$I_{ijkl} \sim \text{Bin}\left(n_{jkl}, p_{ijkl}\right).$$

Given the number of cumulative reported cases for each cell, the resulting cumulative SUAR can be calculated from the ratio of the expected number of cumulative cases to cumulative reported cases as,

$$\frac{\mathbb{E}[I_{ijkl}]}{C_{ijkl}}.$$

Cumulative SUARs adjusted for age group, sex, and health authority were calculated through post-stratification.

Inter-survey period-specific SUARs were also generated overall between all consecutive sero-surveys and additionally by age group between the 6$^{th}$ (September/October 2021) to 7$^{th}$ (March 2022) sero-surveys and 7$^{th}$ to 8$^{th}$ (July/August 2022) sero-surveys. Period-specific SUAR estimates were derived as below, notably assuming no previously infected individuals were re-infected during the specified period, in accordance with surveillance case reporting in BC. First the sero-prevalence rate ($p_{ijkl}$) for each survey period $i$, age group $j$, sex $k$, and health authority $l$ were drawn from the posterior distribution. The SUAR represents the ratio of the modeled number of infections for each period and cell to the total reported cases for the same period within that cell. We model the total infections using a binomial distribution with number of trials being the estimated population size for cell $j, k, l$ ($n_{jkl}$) and success probability being the rate of new infections ($\Delta p_{ijkl}$). Under the assumption of no re-infections and no waning we may calculate the rate of new infections as

$$\Delta p_{ijkl} = p_{ijkl} - p_{i-1jkl}.$$

Given that a negative rate of new infections is implausible, all draws from the posterior where $\Delta p_{ijkl}$ is negative are necessarily excluded. Using the assumed rate of new infections, the simulated number of new infections are drawn from a binomial distribution as

$$\Delta I_{ijkl} \sim \text{Bin}\left(n_{jkl}, \Delta p_{ijkl}\right).$$

Given the reported number of cases between sero-survey $i - 1$ and $i$ ($\Delta C_{ijkl}$), the estimate for the period SUAR can be calculated as,

$$\frac{\mathbb{E}[\Delta I_{ijkl}]}{\Delta C_{ijkl}}.$$

Cumulative surveillance case reports and derived under-ascertainment ratios at the 4th (January 2021) through 8th (July/August 2022) sero-surveys are provided in **Supplementary_Table_9.** Period-specific case reports, attack rates and SUARs between all consecutive sero-surveys are shown in **Supplementary_Table_10,** and by age group between the 6th-7th and 7th-8th sero-surveys in **Figure_5** and **Supplementary_Table_11**.

**References for Supplementary Material 1**

1. BC STATS. Population estimates. Victoria, BC: BC Ministry of Citizens' Services, 2021. [Accessed 8 September 2022]. Available at: https://www2.gov.bc.ca/gov/content/data/statistics/people-population-community/population/population-projections
2. BC STATS. Population projections. (P.E.O.P.L.E) Victoria, BC: BC Ministry of Citizens' Services, 2021. [Accessed 8 September 2022]. Available at: https://www2.gov.bc.ca/gov/content/data/statistics/people-population-community/population/population-projections.
3. British Columbia Centre for Disease Control. BC COVID-19 Data Trends. [Accessed 28 August 2022]. Available: http://www.bccdc.ca/health-info/diseases-conditions/covid-19/data-trends
4. Carazo S, Skowronski DM, Brisson M, et al. Protection against Omicron re-infection conferred by prior heterologous SARS-CoV-2 infection, with and without mRNA vaccination. [in press]. medRxiv pre-print. May 3, 2022. [Accessed 28 August 2022]. Available: https://www.medrxiv.org/content/10.1101/2022.04.29.22274455v2
5. Carazo S, Skowronski DM, Brisson M, et al. Protection against Omicron BA.2 reinfection conferred by primary Omicron or pre-Omicron infection with and without mRNA vaccination: a test-negative case-control study among healthcare workers. Lancet Infec Dis; 2022 [in press].

**Supplementary Table 1.** Sample size, exclusions and dual-assay positivity by sero-survey and testing algorithm

| Assays: Ortho-S1 + Abbott-NP + Siemens-S1-RBD [a] | | | | | | | | | | | | | | | |
|---|---|---|---|---|---|---|---|---|---|---|---|---|---|---|---|
| Age group years | Sero-survey 1: March 2020 [N=1000; 100 per age group] [b] | | | | | Sero-survey 2: May 2020 [N=1000; 100 per age group] | | | | | Sero-survey 3: September 2020 [N=2000; 200 per age group] | | | | |
| | Excluded: insufficient sera for specified testing [c] | Included | Ortho-S1, Abbott-NP, Siemens-S1-RBD Tested [a] | Dual assay SP | SP and Abbott-NP positive | Excluded: insufficient sera for specified testing [c] | Included | Ortho-S1, Abbott-NP, Siemens-S1-RBD Tested [a] | Dual assay SP | SP and Abbott-NP positive | Excluded: insufficient sera for specified testing [c] | Included | Ortho-S1, Abbott-NP, Siemens-S1-RBD Tested [a] | Dual assay SP | SP and Abbott-NP positive |
| All | 105 | 895 | 10 | 2 | 0 | 110 | 890 | 9 | 4 | 4 | 0 | 2000 | 28 | 19 | 14 |
| 0-4 | 63 | 38 | 0 | 0 | 0 | 84 | 16 | 0 | 0 | 0 | 0 | 200 | 2 | 2 | 1 |
| 5-9 | 23 | 76 | 0 | 0 | 0 | 11 | 89 | 0 | 0 | 0 | 0 | 200 | 3 | 1 | 1 |
| 10-19 | 11 | 89 | 0 | 0 | 0 | 5 | 95 | 1 | 1 | 1 | 0 | 200 | 3 | 3 | 3 |
| 20-29 | 1 | 99 | 1 | 0 | 0 | 2 | 98 | 0 | 0 | 0 | 0 | 200 | 4 | 3 | 2 |
| 30-39 | 2 | 98 | 0 | 0 | 0 | 0 | 100 | 1 | 1 | 1 | 0 | 200 | 6 | 5 | 4 |
| 40-49 | 0 | 100 | 1 | 1 | 0 | 1 | 99 | 0 | 0 | 0 | 0 | 200 | 1 | 0 | 0 |
| 50-59 | 2 | 98 | 1 | 1 | 0 | 2 | 98 | 4 | 2 | 2 | 0 | 200 | 3 | 3 | 1 |
| 60-69 | 0 | 100 | 1 | 0 | 0 | 0 | 100 | 0 | 0 | 0 | 0 | 200 | 2 | 1 | 1 |
| 70-79 | 0 | 100 | 2 | 0 | 0 | 1 | 99 | 2 | 0 | 0 | 0 | 200 | 3 | 1 | 1 |
| 80+ | 3 | 97 | 4 | 0 | 0 | 4 | 96 | 1 | 0 | 0 | 0 | 200 | 1 | 0 | 0 |

| Assays: Ortho-S1 + Abbott-NP + Siemens-S1-RBD [a] + supplemental Roche-NP as volume permits | | | | | | | | | | | | | | | | | | | | | | |
|---|---|---|---|---|---|---|---|---|---|---|---|---|---|---|---|---|---|---|---|---|---|---|
| Age group years | Sero-survey 4: January 2021 [N=2000; 200 per age group] | | | | | | | | | | | Sero-survey 5: May/June 2021 [N=2000; 200 per age group] | | | | | | | | | | |
| | Excluded: insufficient sera for specified testing [c] | Included | Ortho-S1, Abbott-NP, Siemens-S1-RBD Tested [a] | Dual assay SP | SP and Abbott-NP positive | SP and Roche-NP tested | SP and Abbott-NP negative Roche-NP negative | SP and Abbot-NP negative, Roche-NP positive | SP and Abbott-NP positive Roche-NP negative | SP and Abbott-NP and Roche-NP positive | SP and Abbott-NP or Roche-NP positive | Excluded: insufficient sera for specifiedl testing [c] | Included | Ortho-S1, Abbott-NP, Siemens-S1-RBD Tested [a] | Dual assay SP | SP and Abbott-NP positive | SP and Roche-NP tested | SP and Abbott-NP negative Roche-NP negative | SP and Abbott-NP negative, Roche-NP positive | SP and Abbott-NP positive Roche-NP negative | SP and Abbott-NP and Roche-NP positive | SP and Abbott-NP or Roche-NP positive |
| All | 1 | 1999 | 97 | 87 | 70 | 65 | 9 | 6 | 1 | 49 | 76 | 9 | 1991 | 1163 | 1060 | 156 | 1039 | 844 | 58 | 2 | 135 | 214 |
| 0-4 | 1 | 199 | 11 | 17 | 16 | 7 | 0 | 1 | 0 | 6 | 17 | 0 | 200 | 24 | 34 | 28 | 27 | 0 | 6 | 0 | 21 | 34 |
| 5-9 | 0 | 200 | 12 | 15 | 15 | 10 | 0 | 0 | 0 | 10 | 15 | 2 | 198 | 36 | 35 | 29 | 27 | 2 | 4 | 0 | 21 | 33 |
| 10-19 | 0 | 200 | 3 | 3 | 3 | 3 | 0 | 0 | 0 | 3 | 3 | 0 | 200 | 45 | 41 | 17 | 38 | 15 | 9 | 0 | 14 | 26 |
| 20-29 | 0 | 200 | 11 | 10 | 6 | 8 | 0 | 3 | 0 | 5 | 9 | 0 | 200 | 101 | 88 | 13 | 87 | 59 | 15 | 0 | 13 | 28 |
| 30-39 | 0 | 200 | 8 | 6 | 4 | 6 | 1 | 1 | 0 | 4 | 5 | 1 | 199 | 122 | 110 | 7 | 109 | 92 | 11 | 0 | 6 | 18 |
| 40-49 | 0 | 200 | 12 | 10 | 8 | 8 | 2 | 0 | 0 | 6 | 8 | 0 | 200 | 149 | 137 | 14 | 137 | 118 | 5 | 1 | 13 | 19 |
| 50-59 | 0 | 200 | 8 | 4 | 3 | 4 | 1 | 0 | 0 | 3 | 3 | 0 | 200 | 158 | 148 | 10 | 148 | 138 | 0 | 0 | 10 | 10 |
| 60-69 | 0 | 200 | 7 | 4 | 3 | 4 | 1 | 0 | 0 | 3 | 3 | 1 | 199 | 180 | 163 | 12 | 162 | 147 | 3 | 0 | 12 | 15 |
| 70-79 | 0 | 200 | 7 | 5 | 4 | 5 | 0 | 1 | 1 | 3 | 5 | 4 | 196 | 177 | 159 | 17 | 159 | 138 | 4 | 0 | 17 | 21 |
| 80+ | 0 | 200 | 18 | 13 | 8 | 10 | 4 | 0 | 0 | 6 | 8 | 1 | 199 | 171 | 145 | 9 | 145 | 135 | 1 | 1 | 8 | 10 |

| Age group years | Assays: Ortho-S1 + Roche-NP + Siemens-S1-RBD [d] | | | | | | | | Assays: Abbott-S1 + Roche-NP + Siemens-S1-RBD [d] | | | |
|---|---|---|---|---|---|---|---|---|---|---|---|---|
| | Sero-survey 6: September-October 2021 [N=2000; 200 per age group] | | | | Sero-survey 7: March 2022 [N=2000; 200 per age group] | | | | Sero-survey 8: July/August 2022 [N=2000; 200 per age group] | | | |
| | Excluded: insufficient sera for specified testing [c] | Included | Dual assay SP | SP and Roche-NP positive | Excluded: insufficient sera for specified testing [b] | Included | Dual assay SP | SP and Roche-NP positive | Excluded: insufficient sera for specified testing [c] | Included | Dual assay SP | SP and Roche-NP positive |
| All | 10 | 1990 | 1502 | 193 | 0 | 2000 | 1864 | 850 | 0 | 2000 | 1921 | 1235 |
| 0-4 | 5 | 195 | 34 | 33 | 0 | 200 | 136 | 132 | 0 | 200 | 164 | 157 |
| 5-9 | 0 | 200 | 28 | 26 | 0 | 200 | 178 | 140 | 0 | 200 | 182 | 147 |
| 10-19 | 0 | 200 | 168 | 26 | 0 | 200 | 193 | 117 | 0 | 200 | 194 | 159 |
| 20-29 | 1 | 199 | 175 | 18 | 0 | 200 | 196 | 102 | 0 | 200 | 200 | 147 |
| 30-39 | 0 | 200 | 178 | 23 | 0 | 200 | 197 | 111 | 0 | 200 | 198 | 139 |
| 40-49 | 1 | 199 | 183 | 22 | 0 | 200 | 195 | 90 | 0 | 200 | 196 | 134 |
| 50-59 | 1 | 199 | 184 | 19 | 0 | 200 | 194 | 64 | 0 | 200 | 197 | 121 |
| 60-69 | 0 | 200 | 180 | 6 | 0 | 200 | 196 | 47 | 0 | 200 | 197 | 79 |
| 70-79 | 0 | 200 | 187 | 15 | 0 | 200 | 192 | 22 | 0 | 200 | 195 | 81 |
| 80+ | 2 | 198 | 185 | 5 | 0 | 200 | 187 | 25 | 0 | 200 | 198 | 71 |

NC = nucleocapsid; S1 = spike 1 protein; S1-RBD = S1 receptor binding domain; SP = sero-positive based on dual-assay positivity

[a] As per specified algorithm for these sero-surveys, Siemens-S1-RBD testing undertaken only if screen positive on Ortho-S1 or Abbott-NP assays
[b] For this sero-survey, sera from 101 children 0-4 years and 99 children 5-9 years were instead provided
[c] At each sero-survey, specimens with insufficient volume to complete the specified algorithm (see Table 1, main manuscript) were excluded only if the requirement for dual positivity could not be resolved otherwise on that basis (e.g. if Ortho-S1 positive, Abbott-NP negative and insufficient serum for Siemens).
[d] For these assays, Roche-NP and Siemens S1-RBD assays applied to all specimens

**Supplementary Table 2.** Distribution by age group, sex and sero-survey

| **Age (years) and sex stratum** | **Sero-survey (N=14,000 specimens collected in total; 13,765 included)** | | | | | | | | **FHA and VCHA population (% only displayed)** | | |
|---|---|---|---|---|---|---|---|---|---|---|---|
| | March 2020 | May 2020 | Sept 2020 | Jan 2021 | May/June 2021 | Sept/Oct 2021 | March 2022 | July/Aug 2022 | 2020 [a] | 2021[b] | 2022 [b] |
| **Overall included (N)** | 895 | 890 | 2000 | 1999 | 1991 | 1990 | 2000 | 2000 | NA | NA | NA |
| Female (n, %) [c] | 452 (50) | 450 (51) | 1000 (50) | 1000 (50) | 997 (50) | 994 (50) | 1000 (50) | 1000 (50) | 51 | 51 | 51 |
| **0-4 (n, %)** [d] | 38 (4) | 16 (2) | 200 (10) | 199 (10) | 201 (10) | 195 (10) | 200 (10) | 200 (10) | 4 | 4 | 4 |
| Female (n, %) [c] | 19 (50) | 11 (69) | 100 (50) | 99 (50) | 101 (50) | 98 (50) | 100 (50) | 100 (50) | 48 | 48 | 48 |
| **5-9 (n, %)** [d] | 76 (8) | 89 (10) | 200 (10) | 200 (10) | 197 (10) | 200 (10) | 200 (10) | 200 (10) | 5 | 5 | 5 |
| Female (n, %) [c] | 38 (50) | 44 (49) | 100 (50) | 100 (50) | 97 (49) | 100 (50) | 100 (50) | 100 (50) | 48 | 48 | 48 |
| **10-19 (n, %)** [d] | 89 (10) | 95 (11) | 200 (10) | 200 (10) | 200 (10) | 200 (10) | 200 (10) | 200 (10) | 10 | 10 | 10 |
| Female (n, %) [c] | 49 (55) | 46 (48) | 100 (50) | 100 (50) | 100 (50) | 100 (50) | 100 (50) | 100 (50) | 49 | 49 | 49 |
| **20-29 (n, %)** [d] | 99 (11) | 98 (11) | 200 (10) | 200 (10) | 200 (10) | 199 (10) | 200 (10) | 200 (10) | 15 | 15 | 15 |
| Female (n, %) [c] | 50 (51) | 50 (51) | 100 (50) | 100 (50) | 100 (50) | 99 (50) | 100 (50) | 100 (50) | 48 | 49 | 49 |
| **30-39 (n, %)** [d] | 98 (11) | 100 (11) | 200 (10) | 200 (10) | 199 (10) | 200 (10) | 200 (10) | 200 (10) | 15 | 15 | 16 |
| Female (n, %) [c] | 49 (50) | 50 (50) | 100 (50) | 100 (50) | 100 (50) | 100 (50) | 100 (50) | 100 (50) | 50 | 50 | 49 |
| **40-49 (n, %)** [d] | 100 (11) | 99 (11) | 200 (10) | 200 (10) | 200 (10) | 199 (10) | 200 (10) | 200 (10) | 13 | 13 | 13 |
| Female (n, %) [c] | 50 (50) | 50 (51) | 100 (50) | 100 (50) | 100 (50) | 99 (50) | 100 (50) | 100 (50) | 52 | 52 | 51 |
| **50-59 (n, %)** [d] | 98 (11) | 98 (11) | 200 (10) | 200 (10) | 200 (10) | 199 (10) | 200 (10) | 200 (10) | 14 | 14 | 14 |
| Female (n, %) [c] | 50 (51) | 49 (50) | 100 (50) | 100 (50) | 100 (50) | 99 (50) | 100 (50) | 100 (50) | 51 | 51 | 52 |
| **60-69 (n, %)** [d] | 100 (11) | 100 (11) | 200 (10) | 200 (10) | 199 (10) | 200 (10) | 200 (10) | 200 (10) | 12 | 12 | 12 |
| Female (n, %) [c] | 50 (50) | 50 (50) | 100 (50) | 100 (50) | 100 (50) | 100 (50) | 100 (50) | 100 (50) | 52 | 52 | 52 |
| **70-79 (n, %)** [d] | 100 (11) | 99 (11) | 200 (10) | 200 (10) | 196 (10) | 200 (10) | 200 (10) | 200 (10) | 7 | 8 | 8 |
| Female (n, %) [c] | 50 (50) | 50 (51) | 100 (50) | 100 (50) | 99 (51) | 100 (50) | 100 (50) | 100 (50) | 53 | 53 | 53 |
| **80+ (n, %)** [d] | 97 (11) | 96 (11) | 200 (10) | 200 (10) | 199 (10) | 198 (10) | 200 (10) | 200 (10) | 4 | 4 | 4 |
| Female (n, %) [c] | 47 (48) | 50 (52) | 100 (50) | 100 (50) | 100 (50) | 99 (50) | 100 (50) | 100 (50) | 58 | 58 | 58 |
| **Median (years)** | 44 | 45 | 39.5 | 40 | 39 | 40 | 39.5 | 39.5 | 40 | 40 | 40 |
| Male | 45 | 45 | 39.5 | 39.5 | 39 | 40 | 39.5 | 39.5 | 38 | 39 | 39 |
| Female | 43 | 44 | 39.5 | 40 | 40 | 39.5 | 39.5 | 39.5 | 41 | 41 | 41 |

Aug = August; FHA = Fraser Health Authority; Jan = January; Oct = October; Sept = September; VCHA = Vancouver Coastal Health Authority

[a] BC STATS. Population estimates. Victoria, BC: BC Ministry of Citizens' Services, 2021. [Accessed 8 September 2022]. Available at: https://www2.gov.bc.ca/gov/content/data/statistics/people-population-community/population/population-projections

[b] BC STATS. Population projections. (P.E.O.P.L.E) Victoria, BC: BC Ministry of Citizens' Services, 2021. [Accessed 8 September 2022]. Available at: https://www2.gov.bc.ca/gov/content/data/statistics/people-population-community/population/population-projections

[c] Percentages female are the percentage of included specimens overall and by age group and sero-survey that were collected from females. The percentage male would be 1 minus the displayed percentage.

[d] Percentages by age group are the percentage of the overall sero-survey tally belonging to that age group

**Supplementary Table 3.** Percentage distribution by health authority, age group and sero-survey

Surveillance for SARS-CoV-2 and COVID-19 cases is administered across the Lower Mainland of British Columbia by two health authorities: Fraser Health Authority (FHA) and Vancouver Coastal Health Authority (VCHA). Displayed below are the percentages of the Lower Mainland general population (FHA and VCHA overall combined) and sero-survey specimens that came from FHA residents, by age group and sero-survey sero-survey.

| **Sero-survey** | **Percentage of FHA + VCHA general population and sero-survey specimens from within FHA, by age group (years) and sero-survey** | | | | | | | | | | |
|---|---|---|---|---|---|---|---|---|---|---|---|
| | **0-4** | **5-9** | **10-19** | **20-29** | **30-39** | **40-49** | **50-59** | **60-69** | **70-79** | **80+** | **Overall** |
| General population (%) | | | | | | | | | | | |
| 2020[a] | 67 | 69 | 68 | 58 | 58 | 61 | 60 | 60 | 59 | 57 | 61 |
| 2021[b] | 65 | 68 | 68 | 59 | 57 | 62 | 61 | 60 | 60 | 58 | 61 |
| 2022[b] | 64 | 68 | 68 | 60 | 57 | 62 | 61 | 61 | 60 | 58 | 61 |
| Sero-survey participants n/N (%) | | | | | | | | | | | |
| 1. March 2020 | 34/38 (89) | 64/76 (84) | 59/89 (66) | 59/99 (60) | 62/98 (63) | 64/100 (64) | 59/98 (60) | 61/100 (61) | 61/100 (61) | 61/97 (63) | 584/895 (65) |
| 2. May 2020 | 14/16 (88) | 66/89 (74) | 59/95 (62) | 62/98 (63) | 45/100 (45) | 57/99 (58) | 52/98 (53) | 60/100 (60) | 54/99 (55) | 52/96 (54) | 521/890 (59) |
| 3. September 2020 | 172/200 (86) | 162/200 (81) | 146/200 (73) | 111/200 (56) | 120/200 (60) | 121200 (61) | 109/200 (55) | 122/200 (61) | 117/200 (59) | 118/200 (59) | 1298/2000 (65) |
| 4. January 2021 | 171/199 (86) | 159/200 (80) | 139/200 (70) | 120/200 (60) | 128/200 (64) | 133/200 (67) | 124/200 (62) | 105/200 (53) | 127/200 (64) | 109/200 (55) | 1315/1999 (66) |
| 5. May/June 2021 | 179/201 (89) | 164/197 (83) | 156/200 (78) | 132/200 (66) | 126/199 (63) | 126/200 (63) | 126/200 (63) | 135/199 (68) | 142/196 (72) | 130/199 (65) | 1416/1991 (71) |
| 6. September/October 2021 | 167/195 (86) | 164/200 (82) | 157/200 (79) | 131/199 (66) | 115/200 (58) | 119/199 (60) | 123/199 (62) | 121/200 (61) | 124/200 (62) | 129/198 (65) | 1350/1990 (68) |
| 7. March 2022 | 170/200 (85) | 161/200 (81) | 161/200 (81) | 139/200 (70) | 110/200 (55) | 128/200 (64) | 133/200 (67) | 119/200 (60) | 111/200 (56) | 123/200 (62) | 1355/2000 (68) |
| 8. July/August 2022 | 183/200 (92) | 161/200 (82) | 144/200 (72) | 147/200 (74) | 163/200 (82) | 158/200 (79) | 154/200 (77) | 119/200 (60) | 124/200 (62) | 119/200 (60) | 1472/2000 (74) |

[a] BC STATS. Population estimates. Victoria, BC: BC Ministry of Citizens' Services, 2021. [Accessed 8 September 2022]. Available at: https://www2.gov.bc.ca/gov/content/data/statistics/people-population-community/population/population-estimates

[b] BC STATS. Population projections. (P.E.O.P.L.E) Victoria, BC: BC Ministry of Citizens' Services, 2021. [Accessed 8 September 2022]. Available at: https://www2.gov.bc.ca/gov/content/data/statistics/people-population-community/population/population-projections

**Supplementary Table 4.** Crude and Bayesian sero-prevalence overall and HA-, age-, sex- stratified: “any” vaccine- and/or infection-induced

| Stratum | Estimation method | Sero-prevalence estimates by sero-survey: any dual-assay positivity, indicative of vaccine- and/or infection-induced antibody [a] | | | | | | | |
|---|---|---|---|---|---|---|---|---|---|
| | | 1. March 2020 | 2. May 2020 | 3. Sept 2020 | 4. January 2021 | 5. May/June 2021 | 6. Sept/Oct 2021 | 7. March 2022 | 8. July/Aug 2022 |
| **OVERALL** | Crude tallies n/N<br>% (95% CI) [b] | 2/895<br>0.2 (0.03, 0.8) | 4/890<br>0.45 (0.1, 1.2) | 19/2000<br>1.0 (0.6, 1.48) | 87/1999<br>4.4 (3.50, 5.3) | 1060/1991<br>53.2 (51.0, 55.45) | 1502/1990<br>75.48 (73.53, 77.4) | 1864/2000<br>93.2 (92.0, 94.3) | 1921/2000<br>96.1 (95.1, 96.9) |
| | **Bayesian adjusted [c] % (95% CrI)** | **0.3 (0.07, 0.8)** | **0.6 (0.2, 1.1)** | **1.0 (0.6, 1.52)** | **4.0 (3.2, 5.0)** | **56.2 (54.1, 58.4)** | **82.7 (81.1, 84.2)** | **95.2 (94.3, 96.0)** | **97.0 (96.2, 97.8)** |
| **By health authority (HA)** | | | | | | | | | |
| Fraser HA (FHA) | Crude tallies n/N<br>% (95% CI) [b] | 1/584<br>0.2 (0.00, 1.0) | 2/521<br>0.4 (0.1, 1.4) | 13/1298<br>1.0 (0.53, 1.7) | 65/1315<br>4.9 (3.8, 6.3) | 728/1416<br>51.4 (48.8, 54.1) | 976/1350<br>72.4 (69.9, 74.8) | 1251/1355<br>92.3 (90.8, 93.7) | 1407/1472<br>95.6 (94.4, 96.6) |
| | **Bayesian adjusted [c] % (95% CrI)** | **0.3 (0.1, 0.8)** | **0.6 (0.2, 1.2)** | **1.0 (0.6, 1.53)** | **4.3 (3.3, 5.3)** | **56.1 (53.46, 58.6)** | **82.1 (80.2, 83.9)** | **94.9 (93.8, 95.9)** | **97.0 (96.2, 97.7)** |
| Vancouver Coastal HA (VCHA) | Crude tallies n/N<br>% (95% CI) [b] | 1/311<br>0.3 (0.01, 1.8) | 2/369<br>0.54 (0.1, 1.9) | 6/702<br>0.9 (0.3, 1.9) | 22/684<br>3.2 (2.0, 4.8) | 332/575<br>57.7 (53.6, 61.8) | 526/640<br>86.2 (79.0, 85.1) | 613/645<br>95.0 (93.1, 96.6) | 514/528<br>97.4 (95.6, 98.54) |
| | **Bayesian adjusted [c] % (95% CrI)** | **0.3 (0.1-0.9)** | **0.6 (0.2, 1.2)** | **1.0 (0.6, 1.6)** | **3.7 (2.6, 5.0)** | **56.4 (52.7, 60.0)** | **83.6 (81.2, 85.9)** | **95.7 (94.2, 96.9)** | **97.1 (95.6, 98.3)** |
| **By sex** | | | | | | | | | |
| Male | Crude tallies n/N<br>% (95% CI) [b] | 1/443<br>0.2 (0.01, 1.3) | 4/440<br>0.9 (0.3, 2.3) | 9/1000<br>0.9 (0.4, 1.7) | 43/1000<br>4.3 (3.1, 5.8) | 528/994<br>53.1 (50.0, 56.3) | 745/996<br>74.8 (72.0, 77.47) | 931/1000<br>93.1 (91.4, 94.6) | 962/1000<br>96.2 (94.8, 97.3) |
| | **Bayesian adjusted [c] % (95% CrI)** | **0.4 (0.06, 0.8)** | **0.6 (0.2, 1.3)** | **1.0 (0.6, 1.54)** | **4.2 (3.1, 5.3)** | **55.4 (52.4, 58.47)** | **81.8 (79.6, 83.8)** | **95.0 (93.8, 96.2)** | **96.9 (95.8, 97.8)** |
| Female | Crude tallies n/N<br>% (95% CI) [b] | 1/452<br>0.2 (0.01, 1.2) | 0/450<br>0 (0-0.8) | 10/1000<br>1.0 (0.48, 1.8) | 44/999<br>4.4 (3.2, 5.9) | 532/997<br>53.4 (50.2, 56.49) | 757/994<br>76.2 (73.4, 78.8) | 933/1000<br>93.3 (91.6, 94.8) | 959/2000<br>95.9 (94.48, 97.0) |
| | **Bayesian adjusted [c] % (95% CrI)** | **0.3 (0.06, 0.8)** | **0.53 (0.2, 1.1)** | **1.0 (0.6, 1.6)** | **3.9 (2.9, 5.1)** | **57.0 (53.9, 60.0)** | **83.6 (81.49, 85.6)** | **95.4 (94.1, 96.4)** | **97.1 (96.1, 98.0)** |
| **By age group** | | | | | | | | | |
| 0-4 years | Crude tallies n/N<br>% (95% CI) [b] | 0/38<br>0 (0-9.3) | 0/16<br>0 (0-20.6) | 2/200<br>1.0 (0.1, 3.6) | 17/199<br>8.54 (5.1, 13.3) | 34/201<br>16.9 (12.0, 22.8) | 34/195<br>17.4 (12.4, 23.50) | 136/200<br>68.0 (61.0, 74.4) | 164/200<br>82.0 (76.0, 87.1) |
| | **Bayesian adjusted [c] % (95% CrI)** | **0.3 (0.046, 0.9)** | **0.6 (0.1, 1.4)** | **1.0 (0.52, 1.7)** | **6.1 (3.9, 9.0)** | **18.7 (13.3, 25.0)** | **17.6 (12.52, 23.49)** | **71.9 (64.8, 78.4)** | **84.48 (77.6, 89.8)** |
| 5-9 years | Crude tallies n/N<br>% (95% CI) [b] | 0/76<br>0 (0-4.7) | 0/89<br>0 (0-4.1) | 1/200<br>0.50 (0.01, 2.8) | 15/200<br>7.50 (4.3, 12.1) | 35/197<br>17.8 (12.7, 23.8) | 28/200<br>14.0 (9.51, 19.6) | 178/200<br>89.0 (83.8, 93.0) | 182/200<br>91.0 (86.2, 94.6) |
| | **Bayesian adjusted [c] % (95% CrI)** | **0.3 (0.04, 1.0)** | **0.53 (0.1, 1.3)** | **1.0 (0.48, 1.6)** | **5.7 (3.7, 8.6)** | **18.4 (13.6, 24.1)** | **14.8 (10.4, 20.2)** | **89.9 (85.51, 93.53)** | **92.8 (89.3, 95.53)** |
| 10-19 years | Crude tallies n/N<br>% (95% CI) [b] | 0/89<br>0 (0-4.1) | 1/95<br>1.1 (0.03-5.7) | 3/200<br>1.50 (0.3, 4.3) | 3/200<br>1.50 (0.3, 4.3) | 41/200<br>20.50 (15.1, 26.8) | 168/200<br>84.0 (78.2, 88.8) | 193/200<br>96.50 (92.9, 98.6) | 194/200<br>97.0 (93.6, 98.9) |
| | **Bayesian adjusted [c] % (95% CrI)** | **0.3 (0.045, 0.9)** | **0.6 (0.2, 1.52)** | **1.1 (0.6, 1.8)** | **3.1 (1.6, 4.8)** | **21.7 (16.4, 27.51)** | **83.8 (78.6, 88.4)** | **96.1 (93.55, 98.2)** | **96.53 (93.9, 98.4)** |
| 20-29 years | Crude tallies n/N<br>% (95% CI) [b] | 0/99<br>0 (0-3.7) | 0/98<br>0 (0-3.7) | 3/200<br>1.50 (0.3, 4.3) | 10/200<br>5.0 (2.4, 9.0) | 88/200<br>44.0 (37.0, 51.2) | 175/199<br>87.9 (82.6, 92.1) | 196/200<br>98.0 (95.0, 99.45) | 200/200<br>100 (98.2, 100) |
| | **Bayesian adjusted [c] % (95% CrI)** | **0.3 (0.1, 0.9)** | **0.54 (0.1, 1.2)** | **1.0 (0.6, 1.8)** | **4.9 (3.0, 7.50)** | **44.2 (37.7, 50.8)** | **87.7 (82.8, 91.9)** | **97.2 (95.0, 98.7)** | **98.8 (97.2, 99.7)** |
| 30-39 years | Crude tallies n/N<br>% (95% CI) [b] | 0/98<br>0 (0-3.7) | 1/100<br>1.0 (0.03-5.45) | 5/200<br>2.50 (0.8, 5.7) | 6/200<br>3.0 (1.1, 6.4) | 110/199<br>55.3 (48.1, 62.3) | 178/200<br>89.0 (83.8, 93.0) | 197/200<br>98.50 (95.7, 99.7) | 198/200<br>99.0 (96.4, 99.9) |
| | **Bayesian adjusted [c] % (95% CrI)** | **0.3 (0.046, 0.9)** | **0.6 (0.2, 1.3)** | **1.1 (0.6, 2.1)** | **3.7 (2.1, 5.55)** | **54.8 (48.2, 61.6)** | **88.6 (84.1, 92.4)** | **97.46 (95.4, 98.9))** | **98.1 (95.7, 99.4)** |
| 40-49 years | Crude tallies n/N<br>% (95% CI) [b] | 1/100<br>1.0 (0.03, 5.45) | 0/99<br>0 (0, 3.7) | 0/200<br>0 (0-1.8) | 10/200<br>5.0 (2.4, 9.0) | 137/200<br>68.50 (61.6, 74.9) | 183/199<br>92.0 (87.3, 95.3) | 195/200<br>97.50 (94.3, 99.2) | 196/200<br>98.0 (95.0, 99.45) |
| | **Bayesian adjusted [c] % (95% CrI)** | **0.4 (0.1, 1.1)** | **0.53 (0.1, 1.2)** | **0.9 (0.4, 1.54)** | **4.4 (2.7, 6.6)** | **68.0 (61.8, 73.8)** | **91.3 (87.3, 94.50)** | **96.8 (94.4, 98.50)** | **97.8 (95.53, 99.1)** |
| 50-59 years | Crude tallies n/N<br>% (95% CI) [b] | 1/98<br>1.0 (0.03, 5.6) | 2/98<br>2.0 (0.3, 7.2) | 3/200<br>1.50 (0.3, 4.3) | 4/200<br>2.0 (0.6, 5.0) | 148/200<br>74.0 (67.3, 80.0) | 184/199<br>92.46 (87.9, 95.7) | 194/200<br>97.0 (93.6, 98.9) | 197/200<br>98.50 (95.7, 99.7) |
| | **Bayesian adjusted [c] % (95% CrI)** | **0.4 (0.1, 1.1)** | **0.6 (0.2, 1.7)** | **1.0 (0.6, 1.8)** | **3.3 (1.8, 5.2)** | **73.1 (67.1, 78.7)** | **91.7 (87.6, 94.9)** | **96.3 (93.8, 98.2)** | **97.9 (95.7, 99.2)** |
| 60-69 years | Crude tallies n/N<br>% (95% CI) [b] | 0/100<br>0 (0-3.6) | 0/100<br>0 (0, 3.6) | 1/200<br>0.50 (0.01, 2.8) | 4/200<br>2.0 (0.6, 5.0) | 163/199<br>81.9 (75.9, 87.0) | 180/200<br>90.0 (85.0, 93.8) | 196/200<br>98.0 (95.0, 99.45) | 197/200<br>98.50 (95.7, 99.7) |
| | **Bayesian adjusted [c] % (95% CrI)** | **0.3 (0.04, 0.9)** | **0.53 (0.1, 1.2)** | **1.0 (0.49, 1.6)** | **3.3 (1.7, 5.2)** | **80.1 (74.3, 85.4)** | **89.6 (85.3, 93.3)** | **97.1 (94.8, 98.7)** | **98.0 (96.1, 99.3)** |
| 70-79 years | Crude tallies n/N<br>% (95% CI) [b] | 0/100<br>0 (0-3.6) | 0/99<br>0 (0, 3.7) | 1/200<br>0.50 (0.01, 2.8) | 5/200<br>2.50 (0.8, 5.7) | 159/196<br>81.1 (74.9, 86.4) | 187/200<br>93.50 (89.1, 96.49) | 192/200<br>96.0 (92.3, 98.3) | 195/200<br>97.50 (94.3, 99.2) |
| | **Bayesian adjusted [c] % (95% CrI)** | **0.3 (0.046, 0.9)** | **0.53 (0.1, 1.2)** | **1.0 (0.48, 1.6)** | **3.49 (1.9, 5.4)** | **79.4 (73.53, 84.7)** | **92.9 (89.2, 95.8)** | **95.7 (93.0, 97.8)** | **97.4 (95.2, 98.9)** |
| 80+ years | Crude tallies n/N<br>% (95% CI) [b] | 0/97<br>0 (0-3.7) | 0/96<br>0 (0, 3.8) | 0/200<br>0 (0-1.8) | 13/200<br>6.50 (3.51, 10.9) | 145/199<br>72.9 (66.1, 78.9) | 185/198<br>93.4 (89.0, 96.46) | 187/200<br>93.50 (89.1, 96.49) | 198/200<br>99.0 (96.4, 99.9) |
| | **Bayesian adjusted [c] % (95% CrI)** | **0.3 (0.1, 0.9)** | **0.53 (0.1, 1.2)** | **0.9 (0.4, 1.53)** | **5.0 (3.1, 7.7)** | **71.9 (65.6, 77.8)** | **92.7 (88.9, 95.8)** | **93.4 (89.6, 96.3)** | **98.2 (96.49, 99.4)** |

Green shading signifies primary analysis based on adjusted Bayesian logistic regression model. 95% CI = 95% confidence interval; 95% CrI = 95% credible interval; Aug = August; Oct = October; Sept = September

[a] Sero-prevalence based on positivity on any two assays included, with or without anti-nucleocapsid protein detection.
[b] 95% CIs around crude sero-prevalence estimates are based on exact method. One-sided 97.5% CIs for cells with zero counts.
[c] HA, sex and age group standardized.

**Supplementary Table 5.** Crude and Bayesian sero-prevalence overall and HA-, age-, sex- stratified: infection-induced, with/without vaccination

| Stratum | Estimation method | Sero-prevalence estimates by sero-survey: dual-assay positivity inclusive of anti-nucleocapsid detection, infection-induced antibody (with/without vaccination [a] | | | | | | | |
|---|---|---|---|---|---|---|---|---|---|
| | | 1. March 2020 | 2. May 2020 | 3. Sept 2020 | 4. January 2021 | 5. May/June 2021 | 6. Sept/Oct 2021 | 7. March 2022 | 8. July/Aug 2022 |
| OVERALL | Crude tallies n/N | 2/895 | 4/890 | 19/2000 | 76/1999 | 214/1991 | 193/1990 | 850/2000 | 1235/2000 |
| | % (95% CI) [b] | 0.2 (0.03, 0.8) | 0.45 (0.1, 1.2) | 1.0 (0.6, 1.48) | 3.8 (3.0, 4.7) | 10.8 (9.4, 12.2) | 9.7 (8.4, 11.1) | 42.50 (40.3, 44.7) | 61.8 (59.6, 63.9) |
| | **Bayesian adjusted [c] % (95% CrI)** | **0.3 (0.07, 0.8)** | **0.6 (0.2, 1.1)** | **1.0 (0.6, 1.52)** | **3.48 (2.7, 4.4)** | **10.1 (8.8, 11.6)** | **9.4 (8.1, 10.8)** | **42.48 (40.2, 44.9)** | **61.1 (58.8, 63.4)** |
| **By health authority (HA)** | | | | | | | | | |
| Fraser HA (FHA) | Crude tallies n/N | 1/584 | 2/521 | 13/1298 | 62/1315 | 184/1416 | 156/1350 | 625/1355 | 974/1472 |
| | % (95% CI) [b] | 0.2 (0.00, 1.0) | 0.4 (0.1, 1.4) | 1.0 (0.53, 1.7) | 4.7 (3.6, 6.0) | 13.0 (11.3, 14.9) | 11.6 (9.9, 13.4) | 46.1 (43.4, 48.8) | 66.2 (63.7, 68.6) |
| | **Bayesian adjusted [c] % (95% CrI)** | **0.3 (0.1, 0.8)** | **0.6 (0.2, 1.2)** | **1.0 (0.6, 1.53)** | **3.8 (2.9, 4.9)** | **11.3 (9.7, 13.0)** | **10.3 (8.7, 12.0)** | **44.7 (42.0, 47.6)** | **65.3 (62.8, 67.7)** |
| Vancouver Coastal HA (VCHA) | Crude tallies n/N | 1/311 | 2/369 | 6/702 | 14/684 | 30/575 | 37/640 | 225/645 | 261/528 |
| | % (95% CI) [b] | 0.3 (0.01, 1.8) | 0.54 (0.1, 1.9) | 0.9 (0.3, 1.9) | 2.1 (1.1, 3.4) | 5.2 (3.6, 7.4) | 5.8 (4.1, 7.9) | 34.9 (31.2, 38.7) | 49.4 (45.1, 53.8) |
| | **Bayesian adjusted [c] % (95% CrI)** | **0.3 (0.1-0.9)** | **0.6 (0.2, 1.2)** | **1.0 (0.6, 1.6)** | **3.0 (2.0, 4.2)** | **8.3 (6.3, 10.51)** | **7.9 (6.0, 9.8)** | **39.0 (35.1, 42.8)** | **54.6 (50.0, 59.1)** |
| **By sex** | | | | | | | | | |
| Male | Crude tallies n/N | 1/443 | 4/440 | 9/1000 | 40/1000 | 109/994 | 103/996 | 419/100 | 634/1000 |
| | % (95% CI) [b] | 0.2 (0.01, 1.3) | 0.9 (0.3, 2.3) | 0.9 (0.4, 1.7) | 4.0 (2.9, 5.4) | 11.0 (9.1, 13.1) | 10.3 (8.52, 12.4) | 41.9 (38.8, 45.0) | 63.4 (60.3, 66.4) |
| | **Bayesian adjusted [c] % (95% CrI)** | **0.4 (0.06, 0.8)** | **0.6 (0.2, 1.3)** | **1.0 (0.6, 1.54)** | **3.7 (2.7, 4.8)** | **10.2 (8.6, 12.1)** | **10.0 (8.2, 11.8)** | **42.53 (39.4, 45.6)** | **63.1 (59.9, 66.2)** |
| Female | Crude tallies n/N | 1/452 | 0/450 | 10/1000 | 36/999 | 105/997 | 90/994 | 431/100 | 601/1000 |
| | % (95% CI) [b] | 0.2 (0.01, 1.2) | 0 (0-0.8) | 1.0 (0.48, 1.8) | 3.6 (2.54, 5.0) | 10.53 (8.7, 12.6) | 9.1 (7.3, 11.0) | 43.1 (40.0, 46.2) | 60.1 (57.0, 63.2) |
| | **Bayesian adjusted [c] % (95% CrI)** | **0.3 (0.06, 0.8)** | **0.53 (0.2, 1.1)** | **1.0 (0.6, 1.6)** | **3.3 (2.4, 4.3)** | **10.0 (8.3, 11.7)** | **8.8 (7.1, 10.51)** | **42.4 (39.3, 45.6)** | **59.2 (56.0, 62.4)** |
| **By age group** | | | | | | | | | |
| 0-4 years | Crude tallies n/N | 0/38 | 0/16 | 2/200 | 17/199 | 34/201 | 33/195 | 132/200 | 157/200 |
| | % (95% CI) [b] | 0 (0-9.3) | 0 (0-20.6) | 1.0 (0.1, 3.6) | 8.54 (5.1, 13.3) | 16.9 (12.0, 22.8) | 16.9 (11.9, 22.9) | 66.0 (59.0, 72.53) | 78.50 (72.2, 84.0) |
| | **Bayesian adjusted [c] % (95% CrI)** | **0.3 (0.046, 0.9)** | **0.6 (0.1, 1.4)** | **1.0 (0.52, 1.7)** | **5.9 (3.6, 8.8)** | **13.4 (9.9, 17.46)** | **13.3 (9.8, 17.4)** | **62.9 (55.8, 69.46)** | **72.6 (65.6, 79.2)** |
| 5-9 years | Crude tallies n/N | 0/76 | 0/89 | 1/200 | 15/200 | 33/197 | 26/200 | 140/200 | 147/200 |
| | % (95% CI) [b] | 0 (0-4.7) | 0 (0-4.1) | 0.50 (0.01, 2.8) | 7.50 (4.3, 12.1) | 17.8 (12.7, 23.8) | 13.0 (8.7, 18.47) | 70.0 (63.1, 76.3) | 73.50 (66.8, 79.48) |
| | **Bayesian adjusted [c] % (95% CrI)** | **0.3 (0.04, 1.0)** | **0.53 (0.1, 1.3)** | **1.0 (0.48, 1.6)** | **5.52 (3.47, 8.4)** | **13.7 (10.1, 17.8)** | **11.3 (8.1, 15.0)** | **65.9 (59.3, 72.2)** | **70.46 (64.6, 76.3)** |
| 10-19 years | Crude tallies n/N | 0/89 | 1/95 | 3/200 | 3/200 | 26/200 | 26/200 | 117/200 | 159/200 |
| | % (95% CI) [b] | 0 (0-4.1) | 1.1 (0.03-5.7) | 1.50 (0.3, 4.3) | 1.50 (0.3, 4.3) | 13.0 (8.7, 18.47) | 13.0 (8.7, 18.47) | 58.50 (51.3, 65.4) | 79.50 (73.2, 84.9) |
| | **Bayesian adjusted [c] % (95% CrI)** | **0.3 (0.045, 0.9)** | **0.6 (0.2, 1.52)** | **1.1 (0.6, 1.8)** | **2.7 (1.4, 4.4)** | **11.8 (8.6, 15.6)** | **11.3 (8.1, 15.0)** | **56.0 (49.54, 62.4)** | **76.0 (70.46, 81.4)** |
| 20-29 years | Crude tallies n/N | 0/99 | 0/98 | 3/200 | 9/200 | 28/200 | 18/199 | 102/200 | 147/200 |
| | % (95% CI) [b] | 0 (0-3.7) | 0 (0-3.7) | 1.50 (0.3, 4.3) | 4.50 (2.1, 8.4) | 14.0 (9.51, 19.6) | 9.1 (5.45, 13.9) | 51.0 (43.9, 58.1) | 73.50 (66.8, 79.48) |
| | **Bayesian adjusted [c] % (95% CrI)** | **0.3 (0.1, 0.9)** | **0.54 (0.1, 1.2)** | **1.0 (0.6, 1.8)** | **4.3 (2.47, 6.7)** | **11.9 (8.6, 15.54)** | **8.9 (6.1, 12.2)** | **49.7 (43.0, 56.6)** | **69.51 (63.3, 75.4)** |
| 30-39 years | Crude tallies n/N | 0/98 | 1/100 | 5/200 | 5/200 | 18/199 | 23/200 | 111/200 | 139/200 |
| | % (95% CI) [b] | 0 (0-3.7) | 1.0 (0.03-5.45) | 2.50 (0.8, 5.7) | 2.50 (0.8, 5.7) | 9.1 (5.45, 13.9) | 11.50 (7.4, 16.8) | 55.50 (48.3, 62.51) | 69.50 (62.6, 75.8) |
| | **Bayesian adjusted [c] % (95% CrI)** | **0.3 (0.046, 0.9)** | **0.6 (0.2, 1.3)** | **1.1 (0.6, 2.1)** | **3.1 (1.6, 4.9)** | **9.51 (6.6, 12.9)** | **10.50 (7.4, 14.2)** | **54.4 (48.2, 60.8)** | **64.6 (57.6, 71.2)** |
| 40-49 years | Crude tallies n/N | 1/100 | 0/99 | 0/200 | 8/200 | 19/200 | 22/199 | 90/200 | 134/200 |
| | % (95% CI) [b] | 1.0 (0.03, 5.45) | 0 (0-3.7) | 0 (0-1.8) | 4.0 (1.7, 7.7) | 9.50 (5.8, 1.4) | 11.1 (7.1, 16.3) | 45.0 (38.0, 52.2) | 67.0 (60.0, 73.47) |
| | **Bayesian adjusted [c] % (95% CrI)** | **0.4 (0.1, 1.1)** | **0.53 (0.1, 1.2)** | **0.9 (0.4, 1.54)** | **3.7 (2.2, 5.8)** | **9.7 (6.9, 12.9)** | **10.1 (7.1, 13.7)** | **44.51 (38.1, 51.3)** | **64.1 (57.6, 70.51)** |
| 50-59 years | Crude tallies n/N | 1/98 | 2/98 | 3/200 | 3/200 | 10/200 | 19/199 | 64/200 | 121/200 |
| | % (95% CI) [b] | 1.0 (0.03, 5.6) | 2.0 (0.3-7.2) | 1.50 (0.3, 4.3) | 1.50 (0.3, 4.3) | 5.0 (2.4, 9.0) | 9.6 (5.9, 14.51) | 32.0 (25.6, 39.0) | 60.50 (53.4, 67.3) |
| | **Bayesian adjusted [c] % (95% CrI)** | **0.4 (0.1, 1.1)** | **0.6 (0.2, 1.7)** | **1.0 (0.6, 1.8)** | **2.8 (1.4, 4.55)** | **7.6 (4.8, 10.7)** | **9.2 (6.4, 12.51)** | **32.7 (26.7, 39.0)** | **59.6 (53.0, 66.3)** |
| 60-69 years | Crude tallies n/N | 0/100 | 0/100 | 1/200 | 3/200 | 15/199 | 6/200 | 47/200 | 79/200 |
| | % (95% CI) [b] | 0 (0-3.6) | 0 (0-3.6) | 0.50 (0.01, 2.8) | 1.50 (0.3, 4.3) | 7.54 (4.3, 12.1) | 3.0 (1.1, 6.4) | 23.50 (17.8, 30.0) | 39.50 (32.7, 46.6) |
| | **Bayesian adjusted [c] % (95% CrI)** | **0.3 (0.04, 0.9)** | **0.53 (0.1, 1.2)** | **1.0 (0.49, 1.6)** | **2.8 (1.4, 4.6)** | **8.7 (5.9, 11.9)** | **5.8 (3.4, 8.7)** | **24.9 (19.55, 30.6)** | **42.3 (36.0, 48.6)** |
| 70-79 years | Crude tallies n/N | 0/100 | 0/99 | 1/200 | 5/200 | 21/196 | 15/200 | 22/200 | 81/200 |
| | % (95% CI) [b] | 0 (0-3.6) | 0 (0-3.7) | 0.50 (0.01, 2.8) | 2.50 (0.8, 5.7) | 10.7 (6.8, 15.9) | 7.50 (4.3, 12.1) | 11.0 (7.0, 16.2) | 40.50 (33.6, 47.7) |
| | **Bayesian adjusted [c] % (95% CrI)** | **0.3 (0.046, 0.9)** | **0.53 (0.1, 1.2)** | **1.0 (0.48, 1.6)** | **3.1 (1.7, 4.8)** | **10.2 (7.3, 13.46)** | **8.2 (5.50, 11.34)** | **14.3 (9.8, 19.3)** | **43.1 (36.55, 49.7)** |
| 80+ years | Crude tallies n/N | 0/97 | 0/96 | 0/200 | 8/200 | 10/199 | 5/198 | 25/200 | 71/200 |
| | % (95% CI) [b] | 0 (0-3.7) | 0 (0-3.8) | 0 (0-1.8) | 4.0 (1.7, 7.7) | 5.0 (2.4, 9.1) | 2.53 (0.8, 5.8) | 12.50 (8.3, 17.9) | 35.50 (28.9, 42.6) |
| | **Bayesian adjusted [c] % (95% CrI)** | **0.3 (0.1, 0.9)** | **0.53 (0.1, 1.2)** | **0.9 (0.4, 1.53)** | **3.7 (2.1, 5.9)** | **7.6 (4.8, 10.7)** | **5.6 (3.2, 8.6)** | **15.4 (11.1, 20.4)** | **37.9 (31.7, 44.3)** |

Green shading signifies primary analysis based on adjusted Bayesian logistic regression model. 95% CI = 95% confidence interval; 95% CrI = 95% credible interval; Aug = August; Oct = October; Sept = September

[a] Sero-prevalence based on positivity on any two assays, of which from the January 2021 sero-survey, at least one must include anti-nucleocapsid detection. Specimens sero-positive on any two assays in prior sero-surveys (1-3) considered infection-induced regardless of assay type.
[b] 95% CIs around crude sero-prevalence estimates are based on exact method. One-sided 97.5% CIs for cells with zero counts.
[c] HA, sex and age group standardized.

**Supplementary Table 6.** Crude and Bayesian sero-prevalence stratified for both age and sex: "any" vaccine- and/or infection-induced

| Age and sex | Estimation method | Sero-prevalence by sero-survey: any dual-assay positivity, indicative of vaccine- and/or infection-induced antibody [a] | | | | | | | |
|---|---|---|---|---|---|---|---|---|---|
| | | 1. March 2020 | 2. May 2020 | 3. Sept 2020 | 4. January 2021 | 5. May/June 2021 | 6. Sept/Oct 2021 | 7. March 2022 | 8. July/Aug 2022 |
| 0-4 years | Male crude tallies n/N; % (95% CI) [b] | 0/19; 0 (0-17.7) | 0/5; 0 (0-52.2) | 1/100; 1.0 (0.02, 5.45) | 9/100; 9.0 (4.2, 16.4) | 18/100; 18.0 (11.0, 27.0) | 16/97; 16.49 (9.7, 25.4) | 65/100; 65.0 (54.8, 74.3) | 80/100; 80.0 (70.8, 87.3) |
| | Female crude tallies n/N; % (95% CI) [b] | 0/19; 0 (0-17.7) | 0/11; 0 (0-28.49) | 2/100; 2.0 (0.2, 7.0) | 8/99; 8.1 (3.6, 15.3) | 16/101; 15.8 (9.3, 24.45) | 17/98; 17.4 (10.4, 26.3) | 71/100; 71.0 (61.1, 79.6) | 84/100; 84.0 (75.3, 90.6) |
| | **Male Bayesian adjusted [c] % (95% CrI)** | **0.3 (0.03, 1.0)** | **0.6 (0.1, 1.7)** | **0.9 (0.4, 1.6)** | **6.3 (3.48, 10.48)** | **18.4 (11.47, 27.2)** | **16.4 (9.8, 24.2)** | **68.6 (58.0, 77.8)** | **83.2 (72.1, 90.6)** |
| | **Female Bayesian adjusted [c] % (95% CrI)** | **0.4 (0.03, 1.0)** | **0.6 (0.1, 1.6)** | **1.1 (0.50, 2.05)** | **5.9 (3.2, 9.6)** | **19.1 (11.4, 28.3)** | **18.9 (11.8, 27.3)** | **75.47 (66.6, 83.0)** | **85.8 (77.1, 92.3)** |
| 5-9 years | Male crude tallies n/N; % (95% CI) [b] | 0/38; 0 (0-9.3) | 0/45; 0 (0-7.9) | 1/100; 1.0 (0.02, 5.45) | 5/100; 5.0 (1.6, 11.3) | 19/100; 19.0 (11.8, 28.1) | 14/100; 14.0 (7.9, 22.4) | 85/100; 85.0 (76.47, 91.4) | 96/100; 96.0 (90.1, 98.9) |
| | Female crude tallies n/N; % (95% CI) [b] | 0/38; 0 (0-9.3) | 0/44; 0 (0-8.0) | 0/100; 0 (0-3.6) | 10/100; 10.0 (4.9, 17.6) | 16/97; 16.49 (9.7, 25.4) | 14/100; 14.0 (7.9, 22.4) | 91/100; 91.0 (83.6, 95.8) | 86/100; 86.0 (77.6, 92.1) |
| | **Male Bayesian adjusted [c] % (95% CrI)** | **0.3 (0.03, 1.0)** | **0.53 (0.09, 1.4)** | **1.0 (0.4, 1.8)** | **4.4 (2.2, 7.6)** | **19.1 (15.6, 26.6)** | **14.53 (8.6, 21.9)** | **88.1 (81.1, 93.4)** | **96.4 (92.6, 98.8)** |
| | **Female Bayesian adjusted [c] % (95% CrI)** | **0.3 (0.03, 1.0)** | **0.54 (0.09, 1.4)** | **0.9 (0.4, 1.6)** | **7.2 (3.8, 12.0)** | **17.8 (11.2, 25.9)** | **15.2 (9.2, 22.8)** | **91.9 (86.3, 95.9)** | **88.9 (82.6, 93.8)** |
| 10-19 years | Male crude tallies n/N; % (95% CI) [b] | 0/40; 0 (0-8.8) | 1/49; 2.0 (0.05, 10.9) | 2/100; 2.0 (0.2, 7.0) | 1/100; 1.0 (0.02, 5.45) | 20/100; 20.0 (12.7, 29.2) | 85/100; 85.0 (76.47, 91.4) | 97/100; 97.0 (91.48, 99.4) | 94/100; 94.0 (87.4, 97.8) |
| | Female crude tallies n/N; % (95% CI) [b] | 0/49; 0 (0-7.3) | 0/46; 0 (0-7.7) | 1/100; 1.0 (0.02, 5.45) | 2/100; 2.0 (0.2, 7.0) | 21/100; 21.0 (13.49, 30.3) | 83/100; 83.0 (74.2, 89.8) | 96/100; 96.0 (90.1, 98.9) | 100/100; 100 (96.4, 100) |
| | **Male Bayesian adjusted [c] % (95% CrI)** | **0.3 (0.03, 1.0)** | **0.7 (0.2, 2.0)** | **1.1 (0.52, 2.3)** | **2.9 (1.2, 5.3)** | **21.3 (14.1, 29.4)** | **84.6 (77.1, 91.0)** | **96.4 (92.7, 98.8)** | **94.3 (89.48, 97.8)** |
| | **Female Bayesian adjusted [c] % (95% CrI)** | **0.3 (0.03, 1.0)** | **0.54 (0.09, 1.4)** | **1.0 (0.45, 1.9)** | **3.2 (1.4, 5.9)** | **22.1 (14.7, 30.4)** | **83.1 (75.1, 89.7)** | **95.8 (91.9, 98.4)** | **98.8 (96.6, 99.8)** |
| 20-29 years | Male crude tallies n/N; % (95% CI) [b] | 0/39; 0 (0-9.0) | 0/48; 0 (0-7.4) | 1/100; 1.0 (0.02, 5.45) | 7/100; 7.0 (2.9, 13.9) | 43/100; 43.0 (33.1, 53.3) | 91/100; 91.0 (83.6, 95.8) | 98/100; 98.0 (93.0, 99.8) | 100/100; 100 (96.4, 100) |
| | Female crude tallies n/N; % (95% CI) [b] | 0/50; 0 (0-7.1) | 0/50; 0 (0-7.1) | 2/100; 2.0 (0.2, 7.0) | 3/100; 3.0 (0.6, 8.52) | 45/100; 45.0 (35.0, 55.3) | 84/99; 84.9 (76.2, 91.3) | 98/100; 98.0 (93.0, 99.8) | 100/100; 100 (96.4, 100) |
| | **Male Bayesian adjusted [c] % (95% CrI)** | **0.3 (0.04, 1.0)** | **0.54 (0.09, 1.4)** | **1.0 (0.46, 1.9)** | **6.0 (3.1, 10.1)** | **43.3 (34.0, 52.4)** | **90.7 (84.6, 95.4)** | **97.1 (93.8, 99.1)** | **98.8 (96.48, 99.8)** |
| | **Female Bayesian adjusted [c] % (95% CrI)** | **0.3 (0.03, 1.0)** | **0.54 (0.09, 1.4)** | **1.1 (0.51, 2.0)** | **3.7 (1.7, 6.52)** | **45.2 (35.3, 55.1)** | **84.4 (76.7, 91.0)** | **97.2 (93.9, 99.2)** | **98.8 (96.54, 99.8)** |
| 30-39 years | Male crude tallies n/N; % (95% CI) [b] | 0/49; 0 (0-7.3) | 1/50; 2.0 (0.05, 10.7) | 3/100; 3.0 (0.6, 8.52) | 2/100; 2.0 (0.2, 7.0) | 58/99; 58.6 (48.2, 68.4) | 86/100; 86.0 (77.6, 92.1) | 99/100; 99.0 (94.6, 100) | 99/100; 99.0 (94.6, 100) |
| | Female crude tallies n/N; % (95% CI) [b] | 0/49; 0 (0-7.3) | 0/50; 0 (0-7.1) | 2/100; 2.0 (0.2, 7.0) | 4/100; 4.0 (1.1, 9.9) | 52/100; 52.0 (41.8, 62.1) | 92/100; 92.0 (84.8, 96.48) | 98/100; 98.0 (93.0, 99.8) | 99/100; 99.0 (94.6, 100) |
| | **Male Bayesian adjusted [c] % (95% CrI)** | **0.3 (0.03, 1.0)** | **0.6 (0.1, 1.7)** | **1.2 (0.6, 2.4)** | **3.3 (1.4, 5.8)** | **58.3 (48.9, 67.54)** | **85.9 (78.7, 91.8)** | **97.7 (94.9, 99.4)** | **97.9 (94.0, 99.6)** |
| | **Female Bayesian adjusted [c] % (95% CrI)** | **0.3 (0.03, 0.9)** | **0.52 (0.1, 1.3)** | **1.1 (0.52, 2.0)** | **4.0 (2.0, 7.0)** | **51.3 (41.9, 60.7)** | **91.3 (85.1, 95.9)** | **97.2 (93.9, 99.1)** | **98.3 (95.2, 99.7)** |
| 40-49 years | Male crude tallies n/N; % (95% CI) [b] | 1/50; 2.0 (0.05, 10.7) | 0/49; 0 (0-7.3) | 0/100; 0 (0-3.6) | 6/100; 6.0 (2.2, 12.6) | 68/100; 68.0 (57.9, 77.0) | 90/100; 90.0 (82.4, 95.1) | 98/100; 98.0 (93.0, 99.8) | 98/100; 98.0 (93.0, 99.8) |
| | Female crude tallies n/N; % (95% CI) [b] | 0/50; 0 (0-7.1) | 0/50; 0 (0-7.1) | 0/100; 0 (0-3.6) | 4/100; 4.0 (1.1, 9.9) | 69/100; 69.0 (59.0, 77.9) | 93/99; 93.9 (87.3, 97.7) | 97/100; 97.0 (91.48, 99.4) | 98/100; 98.0 (93.0, 99.8) |
| | **Male Bayesian adjusted [c] % (95% CrI)** | **0.4 (0.06, 1.50)** | **0.53 (0.09, 1.4)** | **0.9 (0.4, 1.6)** | **4.9 (2.54, 8.3)** | **67.9 (58.7, 76.2)** | **89.3 (83.1, 94.2)** | **97.1 (93.7, 99.1)** | **97.53 (94.2, 99.4)** |
| | **Female Bayesian adjusted [c] % (95% CrI)** | **0.3 (0.03, 1.0)** | **0.53 (0.09, 1.4)** | **0.9 (0.4, 1.6)** | **4.0 (1.9, 6.9)** | **68.2 (59.3, 76.6)** | **93.2 (87.54, 97.0)** | **96.48 (92.9, 98.8)** | **97.8 (94.7, 99.4)** |
| 50-59 years | Male crude tallies n/N; % (95% CI) [b] | 0/48; 0 (0-7.4) | 2/49; 4.1 (0.50, 14.0) | 1/100; 1.0 (0.02, 5.45) | 2/100; 2.0 (0.2, 7.0) | 70/100; 70.0 (60.0, 78.8) | 94/100; 94.0 (87.4, 97.8) | 95/100; 95.0 (88.7, 98.4) | 100/100; 100 (96.4, 100) |
| | Female crude tallies n/N; % (95% CI) [b] | 1/50; 2.0 (0.05, 10.7) | 0/49; 0 (0-7.3) | 2/100; 2.0 (0.2, 7.0) | 2/100; 2.0 (0.2, 7.0) | 78/100; 78.0 (68.6, 85.7) | 90/99; 90.9 (83.4, 95.8) | 99/100; 99.0 (94.6, 100) | 97/100; 97.0 (91.48, 99.4) |
| | **Male Bayesian adjusted [c] % (95% CrI)** | **0.3 (0.04, 1.0)** | **0.8 (0.2, 2.47)** | **1.0 (0.46, 1.8)** | **3.4 (1.4, 6.0)** | **69.48 (60.7, 77.47)** | **93.2 (88.0, 97.9)** | **94.9 (90.6, 97.9)** | **98.8 (96.7, 99.8)** |
| | **Female Bayesian adjusted [c] % (95% CrI)** | **0.4 (0.06, 1.50)** | **0.53 (0.09, 1.4)** | **1.1 (0.51, 2.1)** | **3.3 (1.4, 5.8)** | **76.6 (68.2, 84.0)** | **90.3 (83.9, 95.1)** | **97.6 (94.7, 99.4)** | **97.0 (93.2, 99.1)** |
| 60-69 years | Male crude tallies n/N; % (95% CI) [b] | 0/50; 0 (0-7.1) | 0/50; 0 (0-7.1) | 0/100; 0 (0-3.6) | 2/100; 2.0 (0.2, 7.0) | 81/99; 81.8 (72.8, 88.9) | 88/100; 88.0 (80.0, 93.6) | 98/100; 98.0 (93.0, 99.8) | 98/100; 98.0 (93.0, 99.8) |
| | Female crude tallies n/N; % (95% CI) [b] | 0/50; 0 (0-7.1) | 0/50; 0 (0-7.1) | 1/100; 1.0 (0.02, 5.45) | 2/100; 2.0 (0.2, 7.0) | 82/100; 82.0 (73.1, 89.0) | 92/100; 92.0 (84.8, 96.48) | 98/100; 98.0 (93.0, 99.8) | 99/100; 99.0 (94.6, 100) |
| | **Male Bayesian adjusted [c] % (95% CrI)** | **0.3 (0.03, 1.0)** | **0.53 (0.09, 1.3)** | **0.9 (0.4, 1.7)** | **3.3 (1.4, 5.8)** | **80.1 (71.3, 87.1)** | **87.7 (80.45, 93.1)** | **97.1 (93.9, 99.1)** | **97.8 (95.1, 99.4)** |
| | **Female Bayesian adjusted [c] % (95% CrI)** | **0.3 (0.03, 1.0)** | **0.52 (1.0, 1.3)** | **1.0 (0.4, 1.8)** | **3.3 (1.3, 6.0)** | **80.0 (72.3, 87.0)** | **91.45 (85.8, 95.9)** | **97.2 (93.9, 99.1)** | **98.3 (95.6, 99.7)** |
| 70-79 years | Male crude tallies n/N; % (95% CI) [b] | 0/50; 0 (0-7.1) | 0/49; 0 (0-7.3) | 0/100; 0 (0-3.6) | 3/100; 3.0 (0.6, 8.52) | 79/97; 81.4 (72.3, 88.6) | 89/100; 89.0 (81.2, 94.4) | 98/100; 98.0 (93.0, 99.8) | 97/100; 97.0 (91.48, 99.4) |
| | Female crude tallies n/N; % (95% CI) [b] | 0/50; 0 (0-7.1) | 0/50; 0 (0-7.1) | 1/100; 1.0 (0.02, 5.45) | 2/100; 2.0 (0.2, 7.0) | 80/99; 80.8 (71.7, 88.0) | 98/100; 98.0 (93.0, 99.8) | 94/100; 94.0 (87.4, 97.8) | 98/100; 98.0 (93.0, 99.8) |
| | **Male Bayesian adjusted [c] % (95% CrI)** | **0.3 (0.03, 1.0)** | **0.53 (0.1, 1.30** | **0.9 (0.3, 1.7)** | **3.7 (1.7, 6.4)** | **80.1 (71.7, 87.3)** | **88.6 (82.0, 93.8)** | **97.1 (94.0, 99.1)** | **97.1 (93.50, 99.1)** |
| | **Female Bayesian adjusted [c] % (95% CrI)** | **0.3 (0.03, 0.9)** | **0.53 (0.09, 1.3)** | **1.0 (0.4, 1.8)** | **3.3 (1.4, 5.9)** | **78.8 (70.0, 86.3)** | **96.8 (92.7, 99.1)** | **94.4 (89.9, 97.54)** | **97.6 (94.4, 99.4)** |
| 80+ years | Male crude tallies n/N; % (95% CI) [b] | 0/50; 0 (0-7.1) | 0/46; 0 (0-7.7) | 0/100; 0 (0-3.6) | 6/100; 6.0 (2.2, 12.6) | 72/99; 72.7 (62.9, 81.2) | 92/99; 92.9 (86.0, 97.1) | 96/100; 96.0 (90.1, 98.9) | 100/100; 100 (96.4, 100) |
| | Female crude tallies n/N; % (95% CI) [b] | 0/47; 0 (0-7.6) | 0/50; 0 (0-7.1) | 0/100; 0 (0-3.6) | 7/100; 7.0 (2.9, 13.9) | 73/100; 73.0 (63.2, 81.4) | 93/99; 93.9 (87.3, 97.7) | 91/100; 91.0 (83.6, 95.8) | 98/100; 98.0 (93.0, 99.8) |
| | **Male Bayesian adjusted [c] % (95% CrI)** | **0.3 (0.03, 1.0)** | **0.53 (0.1, 1.3)** | **0.9 (0.4, 1.6)** | **4.8 (2.4, 8.2)** | **72.4 (63.45, 80.3)** | **91.9 (86.0, 96.3)** | **95.8 (91.8, 98.50)** | **98.8 (96.7, 99.8)** |
| | **Female Bayesian adjusted [c] % (95% CrI)** | **0.3 (0.04, 1.0)** | **0.52 (0.1, 1.3)** | **0.9 (0.4, 1.7)** | **5.2 (2.8, 9.0)** | **71.49 (62.2, 79.9)** | **93.3 (88.1, 97.1)** | **91.6 (85.6, 96.0)** | **97.8 (95.0, 99.4)** |

Green shading signifies primary analysis based on adjusted Bayesian logistic regression model.
95% CI = 95% confidence interval; 95% CrI = 95% credible interval; Aug = August; Oct = October; Sept = September

[a] Sero-prevalence based on positivity on any two assays, with or without anti-nucleocapsid protein detection.
[b] 95% CIs around crude sero-prevalence estimates are based on exact method. One-sided 97.5% CIs for cells with zero counts.
[c] HA, sex and age group standardized

**Supplementary Table 7.** Crude and Bayesian sero-prevalence stratified for both age and sex: infection-induced, with/without vaccination

| Age and sex | Estimation method | Sero-prevalence by sero-survey: dual-assay positivity inclusive of anti-nucleocapsid, indicative of infection-induced antibody (with/without vaccination) [a] | | | | | | | |
|---|---|---|---|---|---|---|---|---|---|
| | | 1. March 2020 | 2. May 2020 | 3. Sept 2020 | 4. January 2021 | 5. May/June 2021 | 6. Sept/Oct 2021 | 7. March 2022 | 8. July/Aug 2022 |
| 0-4 years | Male crude tallies n/N; % (95% CI) [b] | 0/19; 0 (0-17.7) | 0/5; 0 (0-52.2) | 1/100; 1.0 (0.02, 5.45) | 9/100; 9.0 (4.2, 16.4) | 18/100; 18.0 (11.0, 27.0) | 16/97; 16.49 (9.7, 25.4) | 62/100; 62.0 (51.8, 71.52) | 78/100; 78.0 (68.6, 85.7) |
| | Female crude tallies n/N; % (95% CI) [b] | 0/19; 0 (0-17.7) | 0/11; 0 (0-28.49) | 2/100; 2.0 (0.2, 7.0) | 8/99; 8.1 (3.6, 15.3) | 16/101; 15.8 (9.3, 24.45) | 17/98; 17.4 (10.4, 26.3) | 70/100; 70.0 (60.0, 78.8) | 79/100; 79.0 (69.7, 86.51) |
| | **Male Bayesian adjusted [c] % (95% CrI)** | **0.3 (0.03, 1.0)** | **0.6 (0.1, 1.7)** | **0.9 (0.4, 1.6)** | **6.1 (3.2, 13.4)** | **13.9 (9.3, 19.50)** | **13.0 (8.45, 18.8)** | **58.7 (48.7, 68.6)** | **72.1 (62.2, 81.3)** |
| | **Female Bayesian adjusted [c] % (95% CrI)** | **00.4 (0.03, 1.0)** | **0.6 (0.1, 1.6)** | **1.1 (0.50, 2.05)** | **5.7 (3.0, 9.6)** | **12.9 (8.4, 18.6)** | **13.52 (8.8, 19.4)** | **67.3 (57.51, 76.0)** | **73.2 (64.0, 81.7)** |
| 5-9 years | Male crude tallies n/N; % (95% CI) [b] | 0/38; 0 (0-9.3) | 0/45; 0 (0-7.9) | 1/100; 1.0 (0.02, 5.45) | 5/100; 5.0 (1.6, 11.3) | 18/100; 18.0 (11.0, 27.0) | 13/100; 13.0 (7.1, 21.2) | 71/100; 71.0 (61.1, 79.6) | 77/100; 77.0 (67.51, 84.8) |
| | Female crude tallies n/N; % (95% CI) [b] | 0/38; 0 (0-9.3) | 0/44; 0 (0-8.0) | 0/100; 0 (0-3.6) | 10/100; 10.0 (4.9, 17.6) | 15/97; 15.46 (8.9. 24.2) | 13/100; 13.0 (7.1, 21.2) | 69/100; 69.0 (59.0, 77.9) | 70/100; 70.0 (90.0, 78.8) |
| | **Male Bayesian adjusted [c] % (95% CrI)** | **0.3 (0.03, 1.0)** | **0.53 (0.09, 1.4)** | **1.0 (0.4, 1.8)** | **4.1 (2.0, 7.4)** | **14.46 (9.7, 20.47)** | **11.3 (7.2, 16.4)** | **66.49 (57.53, 75.3)** | **73.7 (65.3, 51.2)** |
| | **Female Bayesian adjusted [c] % (95% CrI)** | **0.3 (0.03, 1.0)** | **0.54 (0.09, 1.4)** | **0.9 (0.4, 1.6)** | **7.1 (3.8, 11.7)** | **12.8 (8.4, 18.4)** | **11.3 (6.9, 16.6)** | **65.3 (55.8, 74.1)** | **67.0 (58.2, 75.2)** |
| 10-19 years | Male crude tallies n/N; % (95% CI) [b] | 0/40; 0 (0-8.8) | 1/49; 2.0 (0.05, 10.9) | 2/100; 2.0 (0.2, 7.0) | 1/100; 1.0 (0.02, 5.45) | 12/100; 12.0 (6.4, 20.0) | 14/100; 14.0 (7.9, 22.4) | 51/100; 51.0 (40.8, 61.1) | 78/100; 78.0 (68.6, 85.7) |
| | Female crude tallies n/N; % (95% CI) [b] | 0/49; 0 (0-7.3) | 0/46; 0 (0-7.7) | 1/100; 1.0 (0.02, 5.45) | 2/100; 2.0 (0.2, 7.0) | 14/100; 14.0 (7.9, 22.4) | 12/100; 12.0 (6.4, 20.0) | 66/100; 66.0 (55.9, 75.2) | 81/100; 81.0 (71.9, 88.2) |
| | **Male Bayesian adjusted [c] % (95% CrI)** | **0.3 (0.03, 1.0)** | **0.7 (0.2, 2.0)** | **1.1 (0.52, 2.3)** | **2.54 (1.0, 4.9)** | **11.3 (7.0, 16.7)** | **11.8 (7.2, 17.46)** | **49.2 (39.7, 58.4)** | **75.3 (67.4, 82.6)** |
| | **Female Bayesian adjusted [c] % (95% CrI)** | **0.3 (0.03, 1.0)** | **0.54 (0.09, 1.4)** | **1.0 (0.45, 1.9)** | **2.9 (1.1, 5.4)** | **12.4 (7.9, 17.8)** | **10.7 (6.47, 15.8)** | **63.1 (53.53, 72.4)** | **76.8 (65.6, 84.3)** |
| 20-29 years | Male crude tallies n/N; % (95% CI) [b] | 0/39; 0 (0-9.0) | 0/48; 0 (0-7.4) | 1/100; 1.0 (0.02, 5.45) | 6/100; 6.0 (2.2, 12.6) | 12/100; 12.0 (6.4, 20.0) | 8/100; 8.0 (3.52, 15.2) | 49/100; 49.0 (38.9, 59.2) | 76/100; 76.0 (66.4, 84.0) |
| | Female crude tallies n/N; % (95% CI) [b] | 0/50; 0 (0-7.1) | 0/50; 0 (0-7.1) | 2/100; 2.0 (0.2, 7.0) | 3/100; 3.0 (0.6, 8.52) | 16/100; 16.0 (9.4, 24.7) | 10/99; 10.1 (5.0, 17.8) | 53/100; 53.0 (42.8, 63.1) | 71/100; 71.0 (61.1, 79.6) |
| | **Male Bayesian adjusted [c] % (95% CrI)** | **0.3 (0.04, 1.0)** | **0.54 (0.09, 1.4)** | **1.0 (0.46, 1.9)** | **5.1 (2.4, 9.2)** | **10.9 (7.0, 15.8)** | **8.4 (4.8, 12.9)** | **48.3 (39.2, 57.49)** | **72.3 (63.7, 80.3)** |
| | **Female Bayesian adjusted [c] % (95% CrI)** | **0.3 (0.03, 1.0)** | **0.54 (0.09, 1.4)** | **1.1 (0.51, 2.0)** | **3.3 (1.4, 6.1)** | **12.9 (8.4, 18.8)** | **9.49 (5.6, 14.51)** | **51.2 (41.4, 60.8)** | **66.6 (57.4, 75.1)** |
| 30-39 years | Male crude tallies n/N; % (95% CI) [b] | 0/49; 0 (0-7.3) | 1/50; 2.0 (0.05, 10.7) | 3/100; 3.0 (0.6, 8.52) | 2/100; 2.0 (0.2, 7.0) | 11/99; 11.1 (5.7, 19.0) | 12/100; 12.0 (6.4, 20.0) | 57/100; 57.0 (46.7, 66.9) | 66/100; 66.0 (55.9, 75.2) |
| | Female crude tallies n/N; % (95% CI) [b] | 0/49; 0 (0-7.3) | 0/50; 0 (0-7.1) | 2/100; 2.0 (0.2, 7.0) | 3/100; 3.0 (0.6, 8.52) | 7/100; 7.0 (2.9, 13.9) | 11/100; 11.0 (5.6, 18.8) | 54/100; 54.0 (43.7, 64.0) | 73/100; 73.0 (63.2, 81.4) |
| | **Male Bayesian adjusted [c] % (95% CrI)** | **0.3 (0.03, 1.0)** | **0.6 (0.1, 1.7)** | **1.2 (0.6, 2.4)** | **2.9 (1.2, 5.3)** | **10.7 (6.6, 16.0)** | **11.1 (6.8, 16.4)** | **55.7 (46.7, 64.3)** | **61.8 (52.2, 71.2)** |
| | **Female Bayesian adjusted [c] % (95% CrI)** | **0.3 (0.03, 0.9)** | **0.52 (0.1, 1.3)** | **1.1 (0.52, 2.0)** | **3.3 (1.4, 5.9)** | **8.3 (4.7, 12.8)** | **9.9 (6.0, 14.8)** | **53.1 (44.0, 62.4)** | **67.3 (57.8, 76.4)** |
| 40-49 years | Male crude tallies n/N; % (95% CI) [b] | 1/50; 2.0 (0.05, 10.7) | 0/49; 0 (0-7.3) | 0/100; 0 (0-3.6) | 6/100; 6.0 (2.2, 12.6) | 11/100; 11.0 (5.6, 18.8) | 16/100; 16.0 (9.4, 24.7) | 49/100; 49.0 (38.9, 59.2) | 73/100; 73.0 (63.2, 81.4) |
| | Female crude tallies n/N; % (95% CI) [b] | 0/50; 0 (0-7.1) | 0/50; 0 (0-7.1) | 0/100; 0 (0-3.6) | 2/100; 2.0 (0.2, 7.0) | 8/100; 8.0 (3.52, 15.2) | 6/99; 6.1 (2.3, 12.7) | 41/10; 41.0 (31.3, 51.3) | 61/100; 61.0 (50.7, 70.6) |
| | **Male Bayesian adjusted [c] % (95% CrI)** | **0.4 (0.06, 1.50)** | **0.53 (0.09, 1.4)** | **0.9 (0.4, 1.6)** | **4.6 (2.2, 8.1)** | **10.4 (6.3, 15.3)** | **13.0 (8.2, 19.0)** | **47.9 (38.7, 57.7)** | **69.8 (60.8, 78.4)** |
| | **Female Bayesian adjusted [c] % (95% CrI)** | **0.3 (0.03, 1.0)** | **0.53 (0.09, 1.4)** | **0.9 (0.4, 1.6)** | **2.9 (1.1, 5.4)** | **9.0 (5.4, 13.4)** | **7.51 (4.1, 11.9)** | **41.3 (32.6, 50.3)** | **58.9 (49.7, 68.1)** |
| 50-59 years | Male crude tallies n/N; % (95% CI) [b] | 0/48; 0 (0-7.4) | 2/49; 4.1 (0.50, 14.0) | 1/100; 1.0 (0.02, 5.45) | 2/100; 2.0 (0.2, 7.0) | 5/100; 5.0 (1.6, 11.3) | 11/100; 11.0 (5.6, 18.8) | 32/100; 32.0 (23.0, 42.1) | 61/100; 61.0 (50.7, 70.6) |
| | Female crude tallies n/N; % (95% CI) [b] | 1/50; 2.0 (0.05, 10.7) | 0/49; 0 (0-7.3) | 2/100; 2.0 (0.2, 7.0) | 1/100; 1.0 (0.02, 5.45) | 5/100; 5.0 (1.6, 11.3) | 8/99; 8.1 (3.6, 15.3) | 32/100; 32.0 (23.0, 42.1) | 60/100; 60.0 (49.7, 69.7) |
| | **Male Bayesian adjusted [c] % (95% CrI)** | **0.3 (0.04, 1.0)** | **0.8 (0.2, 2.47)** | **1.0 (0.46, 1.8)** | **2.9 (1.2, 5.50)** | **7.47 (4.1, 11.50)** | **10.1 (6.0, 15.3)** | **32.4 (24.0, 41.6)** | **61.1 (52.1, 69.6)** |
| | **Female Bayesian adjusted [c] % (95% CrI)** | **0.4 (0.06, 1.50)** | **0.53 (0.09, 1.4)** | **1.1 (0.51, 2.1)** | **2.6 (1.0, 4.9)** | **7.7 (4.1, 12.0)** | **8.3 (4.7, 12.8)** | **33.0 (24.8, 42.1)** | **58.1 (48.6, 67.7)** |
| 60-69 years | Male crude tallies n/N; % (95% CI) [b] | 0/50; 0 (0-7.1) | 0/50; 0 (0-7.1) | 0/100; 0 (0-3.6) | 2/100; 2.0 (0.2, 7.0) | 7/99; 7.1 (2.9, 14.0) | 3/100; 3.0 (0.6, 8.52) | 23/100; 23.0 (15.2, 32.49) | 42/100; 42.0 (32.2, 52.3) |
| | Female crude tallies n/N; % (95% CI) [b] | 0/50; 0 (0-7.1) | 0/50; 0 (0-7.1) | 1/100; 1.0 (0.02, 5.45) | 1/100; 1.0 (0.02, 5.45) | 8/100; 8.0 (3.52, 15.2) | 3/100; 3.0 (0.6, 8.52) | 24/100; 24.0 (16.0, 33.6) | 37/100; 37.0 (27.6, 47.2) |
| | **Male Bayesian adjusted [c] % (95% CrI)** | **0.3 (0.03, 1.0)** | **0.53 (0.09, 1.3)** | **0.9 (0.4, 1.7)** | **2.9 (1.1, 5.6)** | **8.47 (4.7, 12.9)** | **5.8 (2.7, 9.7)** | **24.6 (17.2, 33.0)** | **44.47 (35.49, 53.55)** |
| | **Female Bayesian adjusted [c] % (95% CrI)** | **0.3 (0.03, 1.0)** | **0.52 (1.0, 1.3)** | **1.0 (0.4, 1.8)** | **2.6 (0.9, 5.0)** | **8.9 (5.2, 13.4)** | **5.9 (2.8, 9.6)** | **25.2 (17.9, 33.6)** | **40.3 (31.8, 49.4)** |
| 70-79 years | Male crude tallies n/N; % (95% CI) [b] | 0/50; 0 (0-7.1) | 0/49; 0 (0-7.3) | 0/100; 0 (0-3.6) | 3/100; 3.0 (0.6, 8.52) | 11/97; 11.3 (5.8, 19.4) | 7/100; 7.0 (2.9, 13.9) | 11/100; 11.0 (5.6, 18.8) | 42/100; 42.0 (32.2, 52.3) |
| | Female crude tallies n/N; % (95% CI) [b] | 0/50; 0 (0-7.1) | 0/50; 0 (0-7.1) | 1/100; 1.0 (0.02, 5.45) | 2/100; 2.0 (0.2, 7.0) | 10/99; 10.1 (5.0, 17.8) | 8/100; 8.0 (3.52, 15.2) | 11/100; 11.0 (5.6, 18.8) | 39/100; 39.0 (29.4, 49.3) |
| | **Male Bayesian adjusted [c] % (95% CrI)** | **0.3 (0.03, 1.0)** | **0.53 (0.1, 1.30** | **0.9 (0.3, 1.7)** | **3.4 (1.4, 6.4)** | **10.51 (6.6, 15.47)** | **8.0 (4.4, 12.6)** | **14.1 (8.4, 20.7)** | **44.51 (35.7, 53.3)** |
| | **Female Bayesian adjusted [c] % (95% CrI)** | **0.3 (0.03, 0.9)** | **0.53 (0.09, 1.3)** | **1.0 (0.4, 1.8)** | **2.9 (1.2, 5.2)** | **9.8 (6.1, 14.53)** | **8.4 (4.8, 13.1)** | **14.4 (8.49, 21.54)** | **41.9 (32.8, 51.0)** |
| 80+ years | Male crude tallies n/N; % (95% CI) [b] | 0/50; 0 (0-7.1) | 0/46; 0 (0-7.7) | 0/100; 0 (0-3.6) | 4/100; 4.0 (1.1, 9.9) | 4/99; 4.0 (1.1, 10.0) | 3/99; 3.1 (0.6, 8.6) | 14/100; 14.0 (7.9, 22.4) | 41/100; 41.0 (31.3, 51.3) |
| | Female crude tallies n/N; % (95% CI) [b] | 0/47; 0 (0-7.6) | 0/50; 0 (0-7.1) | 0/100; 0 (0-3.6) | 4/100; 4.0 (1.1, 9.9) | 6/100; 6.0 (2.2, 12.6) | 2/99; 2.0 (0.3, 7.1) | 11/100; 11.0 (5.6, 18.8) | 30/100; 30.0 (21.2, 40.0) |
| | **Male Bayesian adjusted [c] % (95% CrI)** | **0.3 (0.03, 1.0)** | **0.53 (0.1, 1.3)** | **0.9 (0.4, 1.6)** | **3.7 (1.7, 6.7)** | **7.0 (3.7, 11.0)** | **5.9 (2.9, 9.7)** | **16.6 (10.1, 24.1)** | **43.4 (34.6, 52.4)** |
| | **Female Bayesian adjusted [c] % (95% CrI)** | **0.3 (0.04, 1.0)** | **0.52 (0.1, 1.3)** | **0.9 (0.4, 1.7)** | **3.7 (1.6, 6.7)** | **8.0 (4.4, 12.1)** | **5.4 (2.4, 9.2)** | **14.48 (8.7, 21.4)** | **33.9 (25.8, 42.51)** |

Green shading signifies primary analysis based on adjusted Bayesian logistic regression model.
95% CI = 95% confidence interval; 95% CrI = 95% credible interval; Aug = August; Oct = October; Sept = September

[a] Sero-prevalence based on positivity on any two assays, of which from the January 2021 sero-survey, at least one positive assay must include anti-nucleocapsid protein detection.
[b] 95% CIs around crude sero-prevalence estimates are based on exact method. One-sided 97.5% CIs for cells with zero counts.
[c] HA, sex and age group standardized

**Supplementary Table 8.** Crude SARS-CoV-2 positivity by individual screening assay, age and sero-survey

| Sero-survey and screening assay | | Crude positivity [n/N % (95% confidence interval (CI)) by age group (years), sero-survey and individual screening assay | | | | | | | | | | |
|---|---|---|---|---|---|---|---|---|---|---|---|---|
| | | 0-4 | 5-9 | 10-19 | 20-29 | 30-39 | 40-49 | 50-59 | 60-69 | 70-79 | 80+ | All ages |
| March 2020 | Ortho-S1 | 0/38 (0, 9.3) | 0/77 (0, 4.7) | 0/89 (0, 4.1) | 0/99 (0, 3.7) | 0/98 (0, 3.7) | 1/100 1.0 (0.02, 5.45) | 2/99 2.0 (2.50, 7.1) | 1/100 1.0 (0.02, 5.45) | 1/100 1.0 (0.02, 5.45) | 2/97 2.1 (0.25, 7.3) | 6/897 0.7 (0.3, 1.45) |
| | Siemens-S1-RBD (subset only)[a] | 0/0 | 0/0 | 0/0 | 0/1 (0, 97.50) | 0/0 | 1/1 100 (2.50, 100) | 1/1 100 (2.50, 100) | 0/1 (0, 97.50) | 0/2 (0, 84.2) | 0/4 (0, 60.2) | 2/10 20.0 (2.52, 55.6) |
| | Abbott-NP | 0/38 (0, 9.3) | 0/76 (0, 4.7) | 0/89 (0, 4.1) | 1/99 (1.0) (0.03, 5.50) | 0/98 (0, 3.7) | 0/100 (0, 3.6) | 0/99 (0, 3.7) | 0/100 (0, 3.6) | 1/100 1.0 (0.02, 5.45) | 2/97 2.1 (0.25, 7.3) | 4/896 0.45 (0.1, 1.1) |
| May 2020 | Ortho-S1 | 0/16 (0, 20.6) | 0/89 (0, 4.1) | 1/96 1.0 (0.03, 5.7) | 0/98 (0, 3.7) | 1/100 1.0 (0.02, 5.45) | 0/99 (0, 3.7) | 3/98 3.1 (0.64, 8.7) | 0/100 (0, 3.6) | 1/100 1.0 (0.02, 5.45) | 0/98 (0, 3.7) | 6/894 0.7 (0.3, 1.46) |
| | Siemens-S1-RBD (subset only) [a] | 0/0 | 0/0 | 1/1 100 (2.50, 100) | 0/0 | 1/1 100 (2.50, 100) | 0/0 | 2/4 50.0 (6.8, 93.2) | 0/0 | 0/2 (0, 84.2) | 0/1 (0, 97.50) | 4/9 44.4 (13.7, 78.8) |
| | Abbott-NP | 0/16 (0, 20.6) | 0/89 (0, 4.1) | 1/95 (1.1) (0.03, 5.7) | 0/98 (0, 3.7) | 1/100 1.0 (0.02, 5.45) | 0/99 (0, 3.7) | 3/98 3.1 (0.64, 8.7) | 0/100 (0, 3.6) | 1/99 1.0 (0.03, 5.45) | 1/96 (1.0) (0.03-5.7) | 7/890 0.8 (0.3, 1.6) |
| Sept 2020 | Ortho-S1 | 2/200 1.0 (0.12, 3.6) | 2/200 1.0 (0.1, 3.6) | 3/200 1.50 (0.31, 4.3) | 3/200 1.50 (0.31, 4.3) | 5/200 2.50 (0.82, 5.7) | 1/200 0.50 (0.01, 2.8) | 3/200 1.50 (0.31, 4.3) | 2/200 1.0 (0.12, 3.6) | 0/200 (0, 1.8) | 0/200 (0, 1.8) | 21/2000 1.1 (0.7, 1.6) |
| | Siemens-S1-RBD (subset only) [a] | 2/2 100 (15.8, 100) | 1/3 33.3 (0.8, 90.6) | 3/3 100 (29.2, 100) | 3/4 75.0 (19.4, 99.4) | 5/6 83.3 (35.9, 99.6) | 0/1 (0, 97.50) | 3/3 100 (29.2, 100) | 1/2 50.0 (1.3, 98.7) | 1/3 33.3 (0.8, 90.6) | 0/1 (0, 97.50) | 19/28 67.9 (47.7, 84.1) |
| | Abbott-NP | 1/200 0.50 (0.01, 2.8) | 2/200 1.0 (0.12, 3.6) | 3/200 1.50 (0.31, 4.3) | 3/200 1.50 (0.31, 4.3) | 5/200 2.50 (0.82, 5.7) | 0/200 (0, 1.8) | 1/200 0.50 (0.01, 2.8) | 1/200 0.50 (0.01, 2.8) | 3/200 1.50 (0.31, 4.3) | 1/200 0.50 (0.01, 2.8) | 20/2000 1.0 (0.6, 1.54) |
| Jan 2021 | Ortho-S1 | 18/200 9.0 (5.4, 13.85) | 16/200 8.0 (4.6, 12.7) | 3/200 1.50 (0.3, 4.3) | 10/200 5.0 (2.4, 9.0) | 8/200 4.0 (1.7, 7.7) | 10/200 5.0 (2.4, 9.0) | 6/200 3.0 (1.1, 6.4) | 4/200 2.0 (0.6, 5.0) | 5/200 2.50 (0.8, 5.7) | 16/200 8.0 (4.6, 12.7) | 96/2000 4.8 (3.9, 5.8) |
| | Siemens-S1-RBD (subset only) [a] | 11/11 100 (71.51, 100) | 11/12 91.7 (61.52, 99.8) | 3/3 100 (29.2, 100) | 9/11 81.8 (48.2, 97.7) | 6/8 75.0 (34.9, 96.8) | 9/12 75.0 (42.8, 94.51) | 4/8 50.0 (15.7, 84.3) | 4/7 57.1 (18.4, 90.1) | 5/7 71.4 (29.0, 96.3) | 13/18 72.2 (46.52, 90.3) | 75/97 77.3 (67.7, 85.2) |
| | Abbott-NP | 16/200 8.0 (4.6, 12.7) | 15/200 7.50 (4.3, 12.1) | 3/200 1.50 (0.3, 4.3) | 8/200 4.0 (1.7, 7.7) | 4/200 2.0 (0.6, 5.0) | 11/200 6.0 (3.1, 10.3) | 5/200 2.50 (0.8, 5.7) | 6/200 3.0 (1.1, 6.4) | 6/200 3.0 (1.1, 6.4) | 10/200 5.0 (2.4, 9.0) | 84/2000 4.2 (3.4, 5.2) |
| | Roche-NP (subset only) [b] | 7/7 100 (59.0, 100) | 10/10 100 (69.1, 100) | 3/3 100 (29.2, 100) | 8/9 88.9 (51.8, 99.7) | 5/6 83.3 (35.9, 99.6) | 6/10 60.0 (26.2, 87.8) | 3/6 50.0 (11.8, 88.2) | 3/7 42.9 (9.9, 81.6) | 4/5 80.0 (28.4, 99.49) | 6/11 54.6 (23.4, 83.3) | 55/74 74.3 (62.8, 83.8) |
| | Abbott-NP or Roche-NP | 17/200 8.50 (5.0, 13.3) | 15/200 7.50 (4.3, 12.1) | 3/200 1.50 (0.3, 4.3) | 11/200 5.50 (2.8, 9.6) | 5/200 2.50 (0.8, 5.7) | 11/200 6.0 (3.1, 10.3) | 5/200 2.50 (0.8, 5.7) | 6/200 (3.0) (1.1, 6.4) | 7/200 3.50 (1.4, 7.1) | 10/200 5.0 (2.4, 9.0) | 90/2000 4.50 (3.6, 5.50) |
| May/ June 2021 | Ortho-S1 | 34/201 16.9 (12.0, 22.8) | 37/198 18.7 (13.51, 24.8) | 45/200 22.50 (16.9, 28.9) | 101/200 50.50 (43.4, 57.6) | 123/200 61.50 (54.4, 68.3) | 149/200 74.50 (67.9, 80.4) | 156/200 78.0 (71.6, 83.54) | 181/200 90.50 (85.6, 94.2) | 181/200 90.50 (85.6, 94.2) | 171/200 85.50 (79.8, 90.1) | 1178/1999 58.9 (56.7, 61.1) |
| | Siemens-S1-RBD (subset) [a] | 24/24 100 (85.8, 100) | 34/36 94.4 (81.3, 99.3) | 41/45 91.1 (78.8, 97.52) | 88/101 87.1 (79.0, 93.0) | 109/122 89.3 (82.47, 94.2) | 137/149 92.0 (86.4, 95.8) | 148/158 93.7 (88.7, 96.9) | 163/180 90.6 (85.3, 94.4) | 158/177 89.3 (83.8, 93.4) | 144/171 84.2 (77.9, 89.3) | 1046/1163 89.9 (88.1, 91.6) |
| | Abbott-NP | 28/201 13.9 (9.46, 19.50) | 30/198 15.2 (10.46, 20.9) | 17/200 8.50 (5.0, 13.3) | 13/200 6.50 (3.51, 10.9) | 7/200 3.50 (1.4, 7.1) | 14/200 7.0 (3.9, 11.47) | 12/200 6.0 (3.1, 10.3) | 12/200 6.0 (3.1, 10.3) | 17/200 8.50 (5.0, 13.3) | 10/200 5.0 (2.4, 9.0) | 160/1999 8.0 (6.9, 9.3) |
| | Roche-NP (subset only) [b] | 27/27 100 (87.2, 100) | 25/29 86.2 (68.3, 96.1) | 23/42 54.8 (38.7, 70.1) | 28/100 28.0 (19.48, 37.9) | 17/122 13.9 (8.3, 21.4) | 18/149 12.1 (7.3, 18.4) | 10/158 6.3 (3.1, 11.3) | 15/180 8.3 (4.7, 13.4) | 21/181 11.6 (7.3, 17.2) | 9/172 5.2 (2.4, 9.7) | 193/1160 16.6 (14.54, 18.9) |
| | Abbott-NP or Roche-NP | 34/201 16.9 (12.0, 22.8) | 34/198 17.2 (12.2, 23.2) | 26/200 13.0 (8.7, 18.47) | 28/200 14.0 (9.51, 19.6) | 18/200 9.0 (5.4, 13.9) | 19/200 9.50 (5.8, 14.4) | 12/200 6.0 (3.1, 10.3) | 15/200 7.50 (4.3, 12.1) | 21/200 10.50 (6.6, 15.6) | 11/200 5.50 (2.8, 9.6) | 218/1999 10.9 (9.6, 12.4) |
| Sept/ Oct 2021 | Ortho-S1 | 39/194 20.1 (14.7, 26.4) | 28/199 14.1 (9.6, 19.7) | 168/200 84.0 (78.2, 88.8) | 179/199 90.0 (84.9, 93.8) | 180/200 90.0 (85.0, 93.8) | 184/200 92.0 (87.3, 95.4) | 185/199 93.0 (88.48, 96.1) | 182/200 91.0 (86.2, 94.6) | 190/200 95.0 (91.0, 97.6) | 193/198 97.47 (94.2, 99.2) | 1528/1990 76.8 (74.9, 78.7) |
| | Siemens-S1-RBD | 33/195 16.9 (11.9, 22.9) | 29/200 14.50 (9.9, 20.1) | 168/200 84.0 (78,2, 88.8) | 174/199 87.4 (82.0, 91.7) | 178/200 89.0 (83.8, 93.0) | 183/199 92.0 (87.3, 95.3) | 184/199 92.46 (87.9, 95.7) | 180/200 90.0 (85.0, 93.8) | 187/200 93.50 (89.1, 96.49) | 185/198 93.4 (89.0, 96.46) | 1501/1990 75.4 (73.47, 77.3) |
| | Roche-NP | 33/195 16.9 (11.9, 22.9) | 26/200 13 (8.7, 18.47) | 26/200 13.0 (8.7, 18.47) | 18/199 9.1 (5.45, 13.9) | 23/200 11.50 (7.4, 16.8) | 22/200 11.0 (7.0, 16.2) | 19/199 9.6 (5.9, 14.51) | 6/200 3.0 (1.1, 6.4) | 15/200 7.50 (4.3, 12.1) | 5/198 2.53 (0.8, 5.8) | 193/1991 9.7 (8.4, 11.1) |
| March 2022 | Ortho-S1 | 140/200 70.0 (63.1, 76.3) | 181/200 90.50 (85.6, 94.2) | 193/200 96.50 (92.9, 98.6) | 196/200 98.0 (95.0, 99.45) | 197/200 98.50 (95.7, 99.7) | 196/200 98.0 (95.0, 99.45) | 195/200 97.50 (94.3, 99.2) | 197/200 98.50 (95.7, 99.7) | 195/200 97.50 (94.3, 99.2) | 188/200 94.0 (89.8, 96.9) | 1878/2000 93.9 (92.8, 94.9) |
| | Siemens-S1-RBD | 134/200 67.0 (60.0, 73.47) | 168/200 84.0 (78.2, 88.8) | 190/199 95.48 (91.6. 97.9) | 193/200 96.50 (92.9, 98.6) | 195/200 97.50 (94.3, 99.2) | 193/200 96.50 (92.9, 98.6) | 192/200 96.0 (92.3, 98.3) | 196/200 98.0 (95.0, 99.45) | 190/200 95.0 (91.0, 97.6) | 187/200 93.50 (89.1, 96.49) | 1838/1999 92.0 (90.7, 93.1) |
| | Roche-NP | 133/200 66.50 (59.50, 73.0) | 140/200 70.0 (63.1, 76.3) | 117/200 58.50 (51.3, 65.4) | 102/200 51.0 (43.9, 58.1) | 111/200 55.50 (48.3, 62.51) | 90/200 45.0 (38.0, 52.2) | 64/200 32.0 (25.6, 39.0) | 47/200 23.50 (17.8, 30.0) | 22/200 11.0 (7.0, 16.2) | 25/200 12.50 (8.3, 17.9) | 851/2000 42.6 (40.4, 44.8) |
| July/ Aug 2022 | Abbott-S1 | 158/200 79.0 (72.7, 84.4) | 177/200 88.50 (83.3, 92.6) | 186/200 93 (88.53, 96.1) | 199/200 99.50 (97.3, 100) | 198/200 99.0 (96.4, 99.9) | 193/200 96.50 (92.9, 98.6) | 197/200 98.50 (95.7, 99.7) | 195/200 97.50 (94.3, 99.2) | 195/200 97.50 (94.3, 99.2) | 198/200 99.0 (96.4, 99.9) | 1896/2000 94.8 (93.7, 95.7) |
| | Roche-NP | 159/200 79.50 (73.2, 84.9) | 148/200 74.0 (67.3, 79.9) | 163/200 81.50 (75.4, 86.6) | 147/200 73.50 (66.8, 79.48) | 139/200 69.50 (62.6, 75.8) | 135/200 67.50 (60.53, 73.9) | 122/200 61.0 (53.9, 67.8) | 81/200 40.50 (33.6, 47.7) | 81/200 40.50 (33.6, 47.7) | 71/200 35.50 (28.9, 42.6) | 1246/2000 62.3 (60.1, 64.4) |
| | Siemens-S1-RBD | 183/200 91.50 (86.7, 94.9) | 187/200 93.50 (89.1, 96.49) | 194/200 97.0 (93.6, 98.9) | 200/200 100 (98.2-100) | 198/200 99.0 (96.4, 99.9) | 195/200 97.50 (94.3, 99.2) | 197/200 98.50 (95.7, 99.7) | 198/200 99.0 (96.4, 99.9) | 200/200 100 (98.2-100) | 200/200 100 (98.2-100) | 1952/2000 97.60 (96.8, 98.2) |

All 95% CIs are based on exact method. One-sided 97.5% CIs for cells with zero counts. NC = nucleocapsid; S1 = spike 1; S1-RBD = S1 receptor binding domain.

[a] Only specimens that were positive on Ortho-S1 or Abbott-NP screening assays and with sufficient residual sera were tested by Siemens-S1-RBD for these sero-surveys.
[b] Only specimens that were positive on Ortho-S1 or Abbott-NP screening assays and with sufficient residual sera were tested by Roche-NP for these sero-surveys.

**Supplementary Table 9.** Cumulative SUARs, 4th-8th sero-surveys

| Cumulative surveillance under-ascertainment ratios (SUARs) | Sero-surveys | | | | |
|---|---|---|---|---|---|
| | 4. January 2021 | 5. May/June 2021 | 6. Sept/Oct 2021 | 7. March 2022 | 8. July/Aug 2022 |
| Epi-week span, calendar date (inclusive) | To end epi-week 2 (January 16) | To end epi-week 21 (May 29) | To end epi-week 38 (September 25) | To end epi-week 10 (March 12) | To end epi-week 30 (July 30) |
| Cumulative cases to end of epi-week span [a] | 50,336 | 116,933 | 136,234 | 225,178 | 239,263 |
| **Bayesian SUAR [b,c] (95% CrI)** | **1.6 (0.9, 2.3)** | **2.4 (1.9, 2.9)** | **1.9 (1.4, 2.3)** | **5.3 (4.8, 5.7)** | **7.9 (7.50, 8.3)** |

95% CrI = 95% credible interval; FHA = Fraser Health Authority; HA = Health Authority; SUAR = surveillance under-ascertainment ratio VCHA = Vancouver Coastal Health Authority

See **Supplementary Material 3** for methodological details related to SUAR estimation. Population estimates include long-term care facility (LTCF) and assisted (ALF) or independent living facility (ILF) residents; whereas, sero-survey sampling and surveillance case report tallies excluded these individuals. There may be ~50,000 LTCF/ALF/ILF residents in British Columbia (BC)[d,e]; recognizing about half the population of elderly adults 65+ years old in BC reside in FHA + VCHA we may estimate ~25,000 LTCF/ALF/ILF residents in the Lower Mainland included in population estimates used to derive estimated infections whereas surveillance case reports excluded these individuals. This suggests SUAR's in the elderly may be slight over-estimates.

[a] Surveillance case reports based upon episode date hierarchically defined by onset date or if not available then specimen collection date or if not available then test result date. Excludes out of province cases and residents of long-term care facilities or assisted living facilities.
[b] Age, sex and HA standardized
[c] BC STATS. Population projections. (P.E.O.P.L.E) Victoria, BC: BC Ministry of Citizens' Services, 2021. [Accessed 8 September 2022]. Available at: https://www2.gov.bc.ca/gov/content/data/statistics/people-population-community/population/population-projections. Lower Mainland population (Fraser Health Authority + Vancouver Coastal Health Authority) estimated to be 3,203,743 for sero-surveys 4-6 and 3,249,077 for sero-surveys 7-8.
[d] Canadian Institute for Health Information. 2021. How many long-term care beds are there in Canada? [Accessed August 10, 2022]. Available: https://www.cihi.ca/en/how-many-long-term-care-beds-are-there-in-canada#:~:text=In%20Canada%2C%20there%20are%202%2C076,of%20February%2028%2C%202021
[e] Statistics Canada. A profile of nursing and residential care facilities, 2019. [Accessed August 10, 2022]. Available: https://www150.statcan.gc.ca/n1/daily-quotidien/210916/dq210916c-eng.htm

**Supplementary Table 10.** Period-specific SUARs, overall, between all consecutive sero-surveys

| | Span of sero-survey snapshots defining period-specific SUAR analyses | | | | | | |
|---|---|---|---|---|---|---|---|
| **Period between sero-surveys:** | **1-2** [a] March – May, 2020 | **2-3** [a] May – September, 2020 | **3-4** [a] September, 2020 – January, 2021 | **4-5** [b] January- May, 2021 | **5-6** [b] May/June– September, 2021 | **6-7** [b] September/October, 2021 – March, 2022 | **7-8** [c] March – July/August, 2022 |
| Period-specific epi-week span | 10-20 | 21-38 | 39-2 | 3-21 | 22-38 | 39-10 | 11-30 |
| Period-specific surveillance case report tallies [d] | 1,784 | 5,331 | 43,181 | 66,597 | 19,301 | 88,944 | 14,085 |
| Period-specific Δ Bayesian sero-prevalence (%) [e] (95% CrI) | 0.2 (-0.4 – 0.9) | 0.5 (-0.2 – 1.1) | 3.0 (2.0, 4.1) | 6.6 (5.0, 8.3) | -0.7 (-2.6, 1.2) | 33.1 (30.47, 35.8) | 18.6 (15.4, 21.9) |
| **Bayesian SUAR [f,g,h] (95% CrI)** | **17.1 (1.8, 48.8)** | **5.2 (1.46, 9.7)** | **3.0 (2.2, 4.0)** | **3.8 (3.0, 4.7)** | **7.8 (11.0, 13.2)** | **12.1 (11.0, 13.2)** | **91.9 (75.3, 110.1)** |

95% CrI = 95% credible interval; SP = sero-prevalence; SUAR = surveillance under-ascertainment ratio.

See **Supplementary Material 3** for methodological details related to SUAR estimation. Population estimates include long-term care facility (LTCF) and assisted (ALF) or independent living facility (ILF) residents; whereas, sero-survey sampling and surveillance case report tallies excluded these individuals.

[a] 2020 population estimate (Vancouver Coastal and Fraser Health Authorities, combined): 3,173,160 based on: BC STATS. Population estimates. Victoria, BC: BC Ministry of Citizens' Services, 2021. [Accessed 8 September 2022]. Available at: https://www2.gov.bc.ca/gov/content/data/statistics/people-population-community/population/population-projections

[b] 2021 population estimate (Vancouver Coastal and Fraser Health Authorities, combined): 3,203,743 based on: BC STATS. Population projections. (P.E.O.P.L.E) Victoria, BC: BC Ministry of Citizens' Services, 2021. [Accessed 8 September 2022]. Available at: https://www2.gov.bc.ca/gov/content/data/statistics/people-population-community/population/population-projections.

[c] 2022 population estimate (Vancouver Coastal and Fraser Health Authorities, combined): 3,249,077 based on: BC STATS. Population projections. (P.E.O.P.L.E) Victoria, BC: BC Ministry of Citizens' Services, 2021. [Accessed 8 September 2022]. Available at: https://www2.gov.bc.ca/gov/content/data/statistics/people-population-community/population/population-projections.

[d] Surveillance case reports based upon episode date hierarchically defined by onset date or if not available then specimen collection date or if not available then test result date. Excludes out of province cases and residents of long-term care facilities or assisted living facilities.

[e] Infection-induced sero-prevalence based on dual-assay positivity, of which from January 2021 at least one positive assay must include anti-nucleocapsid protein detection. Bayesian estimates age, sex, and HA standardized.

[f] Assuming no previously infected are re-infected during the specified analysis period

[g] Age, sex and HA standardized.

[h] Period-specific SUAR estimates exclude samples from the posterior where the difference in sero-prevalence between sero-surveys is less than zero.

**Supplementary Table 11.** Period-specific SUARs, overall and by age, between 6th-7th and 7th-8th sero-surveys

| | Age group (years) | | | | | | | | | | Overall |
|---|---|---|---|---|---|---|---|---|---|---|---|
| | 0-4 | 5-9 | 10-19 | 20-29 | 30-39 | 40-49 | 50-59 | 60-69 | 70-79 | 80+ | |
| **General population estimates, FHA + VCHA, combined** | | | | | | | | | | | |
| 2021 Population Estimates [a] | 142,779 | 149,664 | 314,910 | 473,049 | 495,620 | 419,465 | 444,663 | 382,294 | 243,627 | 137,672 | 3,203,743 |
| 2022 Population Estimates [a] | 142,825 | 150,102 | 317,492 | 472,965 | 507,929 | 425,441 | 442,950 | 392,127 | 254,142 | 143,104 | 3,249,077 |
| **Between 6th and 7th sero-surveys** (September/October, 2021 – March, 2022) | | | | | | | | | | | |
| Period-specific surveillance case report tallies epi-weeks 39-10 (September 26,2021-March 12, 2022) [b] | 5,265 | 5,296 | 7,667 | 15,587 | 17,097 | 13,502 | 10,339 | 6,783 | 3,545 | 3,856 | 88,944 |
| Period-specific Δ Bayesian sero-prevalence (%) (95% CrI) [c,d,e] | 49.6 (41.6, 57.4) | 54.7 (47.4, 61.9) | 44.7 (37.1, 52.2) | 40.8 (33.4, 48,2) | 43.9 (36.5, 51.1) | 34.4 (27.2, 42.1) | 23.53 (16.7, 30.46) | 19.1 (12.9, 25.4) | 6.1 (0.7, 11.8) | 9.7 (4.4, 15.47) | 33.1 (30.47, 35.8) |
| Bayesian SUAR [c,d f] (95% CrI) | 13.4 (11.3, 15.51) | 15.49 (13.3, 17.6) | 19.3 (15.8, 22.6) | 13.2 (10.7, 15.7) | 13.2 (11.0, 15.3) | 10.8 (8.45, 13.4) | 10.8 (7.7, 14.1) | 10.7 (7.3, 14.4) | 5.1 (1.8, 9.2) | 3.4 (1.7, 5.3) | 12.1 (11.0, 13.2) |
| **Between 7th and 8th sero-surveys** (March – July/August, 2022) | | | | | | | | | | | |
| Period-specific surveillance case report tallies epi-weeks 11-30 (March 13-July 30, 2022) [b] | 824 | 135 | 246 | 1,206 | 1,745 | 1,181 | 1,341 | 1,534 | 2,082 | 3,790 | 14,085 |
| Period-specific Δ Bayesian sero-prevalence (%) [c] (95% CrI) [c,d,e] | 9.8 (0.2, 19.4) | 4.5 (4.2, 13.3) | 20.1 (11.6, 28.6) | 19.8 (10.52, 29.1) | 10.2 (0.9, 19.4) | 19.6 (10.4, 28.9) | 26.8 (17.9, 35.8) | 17.4 (8.7, 25.9) | 28.8 (20.7, 36.54) | 22.5 (14.6, 30.46) | 18.6 (15.4, 21.9) |
| Bayesian SUAR [c,d,f] (95% CrI) | 23.9 (10.6, 39.6) | 115.8 (25.4, 262.9) | 313.1 (192.51, 433.9) | 101.2 (60.9, 147.0) | 39.6 (16.9, 68.6) | 78.4 (44.4, 113.6) | 95.4 (63.8, 128.3) | 44.7 (25.1, 64.8) | 33.1 (23.6, 42.2) | 8.6 (5.8, 11.8) | 91.9 (75.3, 110.1) |

95% CrI = 95% credible interval; FHA = Fraser Health Authority; HA = Health Authority; SUAR = surveillance under-ascertainment ratio VCHA = Vancouver Coastal Health Authority

See **Supplementary Material 3** for methodological details related to SUAR estimation. Population estimates include long-term care facility (LTCF) and assisted (ALF) or independent living facility (ILF) residents; whereas, sero-survey sampling and surveillance case report tallies excluded these individuals. There may be at most ~50,000 LTCF/ALF/ILF residents in British Columbia (BC)[g,h]; recognizing about half the population of elderly adults 65+ years old in BC reside in FHA + VCHA we may estimate at most about 25,000 LTCF/ALF/ILF residents in the Lower Mainland included in population estimates used to derive estimated infections whereas surveillance case reports excluded these individuals. This suggests SUAR's in the elderly could be slight over-estimates.

[a] BC STATS. Population projections. (P.E.O.P.L.E) Victoria, BC: BC Ministry of Citizens' Services, 2021. [Accessed 5 July 2022]. Available at: https://www2.gov.bc.ca/gov/content/data/statistics/people-population-community/population/population-projections. Although excluded from sero-survey sampling, VCHA population estimates include certain coastal areas that are not typically considered part of the Lower Mainland, BC.

[b] Surveillance case reports based upon episode date hierarchically defined by onset date or if not available then specimen collection date or if not available then test result date. Excludes out of province cases and residents of long-term care facilities or assisted living facilities.

[c] Infection-induced sero-prevalence based on orthogonal dual-assay positivity, of which at least one positive assay must include anti-nucleocapsid protein detection. Bayesian estimates age, sex, and HA standardized.

[d] Assuming no previously infected are re-infected during the specified analysis period

[e] Age, sex and HA standardized.

[f] Period-specific SUAR estimates exclude samples from the posterior where the difference in sero-prevalence between sero-surveys is less than zero.

[g] Canadian Institute for Health Information. 2021. How many long-term care beds are there in Canada? [Accessed August 10, 2022]. Available: https://www.cihi.ca/en/how-many-long-term-care-beds-are-there-in-canada#:~:text=In%20Canada%2C%20there%20are%202%2C076,of%20February%2028%2C%202021

[h] Statistics Canada. A profile of nursing and residential care facilities, 2019. [Accessed August 10, 2022]. Available: https://www150.statcan.gc.ca/n1/daily-quotidien/210916/dq210916c-eng.htm

**Supplementary Table 12.** Exploratory analysis: Bayesian sero-prevalence estimates, overall and by age, adjusted for sensitivity and specificity

| Stratum | Bayesian adjusted estimates [a] | Sero-prevalence estimates by sero-survey: Dual-assay positivity inclusive of anti-nucleocapsid detection, infection-induced antibody (with/without vaccination) [b] % (95% CrI) | | | | | | | |
|---|---|---|---|---|---|---|---|---|---|
| | | 1. March 2020 | 2. May 2020 | 3. Sept 2020 | 4. January 2021 | 5. May/June 2021 | 6. Sept/Oct 2021 | 7. March 2022 | 8. July/Aug 2022 |
| OVERALL | Any | 0.2 (0.01, 0.7) | 0.3 (0.01, 0.9) | 0.54 (0.04, 1.2) | 3.4 (2.0, 4.6) | 56.4 (54.0, 59.0) | 87.8 (84.1, 90.4) | 95.7 (94.49, 97.1) | 97.3 (96.4, 98.2) |
| | Infection-induced | 0.2 (0.01, 0.7) | 0.3 (0.01, 0.9) | 0.54 (0.04, 1.2) | 3.3 (2.1, 4.3) | 10.3 (8.6, 11.9) | 8.5 (6.7, 10.3) | 42.4 (39.8 - 45.3) | 61.5 (58.7, 64.6) |
| By age group | | | | | | | | | |
| 0-4 years | Any | 0.2 (0.01, 0.8) | 0.3 (0.01, 1.1) | 0.53 (0.03, 1.3) | 5.6 (3.1, 8.6) | 18.2 (12.7, 24.6) | 16.6 (11.3, 22.4) | 71.8 (64.8, 78.3) | 84.4 (76.9, 90.0) |
| | Infection-induced | 0.2 (0.01, 0.8) | 0.3 (0.01, 1.1) | 0.53 (0.03, 1.3) | 5.9 (3.4, 9.1) | 13.7 (10.1, 18.1) | 12.53 (8.8, 16.9) | 63.4 (56.2, 70.54) | 73.4 (66.2, 80.4) |
| 5-9 years | Any | 0.2 (0.01, 0.7) | 0.3 (0.01,1.0) | 0.51 (0.03, 1.2) | 5.2 (2.8, 8.2) | 18.1 (13.1, 23.6) | 13.8 (9.1, 19.2) | 90.4 (85.51, 94.49) | 93.1 (89.47, 96.0) |
| | Infection-induced | 0.2 (0.01, 0.7) | 0.3 (0.01,1.0) | 0.51 (0.03, 1.2) | 5.47 (3.1, 8.6) | 14.0 (10.2, 18.6) | 10.46 (6.9, 14.4) | 66.6 (59.9, 73.3) | 71.1 (65.1, 77.0) |
| 10-19 years | Any | 0.2 (0.01, 0.7) | 0.3 (0.01, 1.2) | 0.6 (0.03, 1.46) | 2.48 (0.8, 4.3) | 21.3 (15.7, 27.2) | 90.4 (83.3, 96.9) | 96.7 (93.9, 98.9) | 96.7 (94.1, 98.7) |
| | Infection-induced | 0.2 (0.01, 0.7) | 0.3 (0.01, 1.2) | 0.6 (0.03, 1.46) | 2.48 (0.9, 4.4) | 12.1 (8.7, 16.1) | 10.4 (6.9, 14.6) | 56.2 (49.1, 63.3) | 76.8 (70.7, 82.8) |
| 20-29 years | Any | 0.2 (0.01, 0.7) | 0.3 (0.01, 0.9) | 0.6 (0.03, 1.4) | 4.3 (2.0, 7.0) | 44.2 (37.4, 51.0) | 93.9 (87.6, 98.8) | 97.6 (95.2, 99.3) | 99.0 (97.4, 99.8) |
| | Infection-induced | 0.2 (0.01, 0.7) | 0.3 (0.01, 0.9) | 0.6 (0.03, 1.4) | 4.1 (2.2, 6.7) | 12.1 (8.6, 16.2) | 8.0 (5.0, 11.49) | 49.9 (43.1, 56.9) | 70.2 (63.5, 76.8) |
| 30-39 years | Any | 0.2 (0.01, 0.7) | 0.3 (0.01, 1.1) | 0.6 (0.03, 1.6) | 3.0 (1.1, 5.0) | 54.9 (48.0, 61.7) | 94.8 (88.7, 99.3) | 97.9 (95.7, 99.46) | 98.4 (96.0, 99.7) |
| | Infection-induced | 0.2 (0.01, 0.7) | 0.3 (0.01, 1.1) | 0.6 (0.03, 1.6) | 2.8 (1.2, 4.9) | 9.7 (6.6, 13.1) | 9.7 (6.2, 13.6) | 54.7 (47.7, 61.4) | 65.1 (57.8, 72.2) |
| 40-49 years | Any | 0.2 (0.01, 0.8) | 0.3 (0.01, 1.0) | 0.49 (0.03, 1.2) | 3.8 (1.7, 6.2) | 68.4 (61.7, 74.6) | 96.6 (91.6, 99.6) | 97.3 (94.7, 99.3) | 98.0 (95.7, 99.53) |
| | Infection-induced | 0.2 (0.01, 0.8) | 0.3 (0.01, 1.0) | 0.49 (0.03, 1.2) | 3.5 (1.7, 5.8) | 9.8 (6.8, 13.4) | 9.3 (5.8, 13.2) | 44.51 (37.8, 51.4) | 64.6 (57.9, 71.48) |
| 50-59 years | Any | 0.2 (0.01, 0.9) | 0.4 (0.01, 1.2) | 0.6 (0.03, 1.4) | 2.7 (0.9, 4.6) | 73.50 (67.1, 79.4) | 97.0 (92.2, 99.8) | 96.8 (94.1, 98.9) | 98.1 (95.8, 99.54) |
| | Infection-induced | 0.2 (0.01, 0.9) | 0.4 (0.01, 1.2) | 0.6 (0.03, 1.4) | 2.6 (1.0, 4.53) | 7.7 (4.6, 11.1) | 8.3 (5.0, 11.9) | 32.4 (25.9, 39.1) | 60.0 (52.9, 67.0) |
| 60-69 years | Any | 0.2 (0.01, 0.7) | 0.3 (0.01, 1.0) | 0.51 (0.03, 1.2) | 2.6 (1.0, 4.6) | 80.6 (74.9, 86.2) | 95.9 (90.3, 99.6) | 97.6 (95.3, 99.4) | 98.4 (96.52, 99.6) |
| | Infection-induced | 0.2 (0.01, 0.7) | 0.3 (0.01, 1.0) | 0.51 (0.03, 1.2) | 2.51 (1.0, 4.3) | 8.8 (5.7, 12.1) | 5.0 (2.48, 8.1) | 24.3 (18.50, 30.4) | 42.1 (35.4, 49.1) |
| 70-79 years | Any | 0.2 (0.01, 0.7) | 0.3 (0.01, 1.0) | 0.52 (0.03, 1.2) | 2.8 (1.0, 4.8) | 79.9 (73.7, 85.7) | 97.2 (92.9, 99.8) | 96.2 (93.1, 98.8) | 97.8 (95.47, 99.4) |
| | Infection-induced | 0.2 (0.01, 0.7) | 0.3 (0.01, 1.0) | 0.52 (0.03, 1.2) | 2.9 (1.2, 4.9) | 10.3 (7.2, 13.8) | 7.3 (4.3, 10.6) | 13.47 (8.7, 18.6) | 43.0 (36.3, 49.7) |
| 80+ years | Any | 0.2 (0.01, 0.7) | 0.3 (0.01, 1.0) | 0.49 (0.03, 1.2) | 4.3 (2.0, 7.1) | 72.2 (65.8, 78.4) | 97.7 (93.4, 99.8) | 94.0 (90.0, 97.3) | 98.52 (96.6, 99.7) |
| | Infection-induced | 0.2 (0.01, 0.7) | 0.3 (0.01, 1.0) | 0.49 (0.03, 1.2) | 3.47 (1.6, 5.8) | 7.7 (4.6, 10.9) | 4.9 (2.4, 7.9) | 14.6 (9.7, 20.0) | 37.8 (31.3, 44.7) |

95% CrI = 95% credible interval; Aug = August; BC = British Columbia, Canada; Oct = October; Sept = September. Sensitivity and specificity considerations are addressed in **Supplementary Material 2**.

[a] HA, sex and age group standardized and adjusted for sensitivity and specificity.
[b] Sero-prevalence based on positivity on dual-assay positivity, of which from the January 2021 sero-survey, at least one must include anti-nucleocapsid protein detection. Dual-assay positive specimens in prior sero-surveys (1-3) considered infection-induced regardless of assay type.

**Supplementary Table 13.** Exploratory analysis: Period-specific SUARs, overall, adjusted for sensitivity and specificity

| | **Span of sero-survey snapshots defining period-specific SUAR analyses** | | | | | | |
|---|---|---|---|---|---|---|---|
| **Period between sero-surveys:** | **1-2**<br>March –<br>May, 2020 | **2-3**<br>May –<br>September, 2020 | **3-4**<br>September, 2020 –<br>January, 2021 | **4-5**<br>January-<br>May, 2021 | **5-6**<br>May/June–<br>September, 2021 | **6-7**<br>September/October, 2021 –<br>March, 2022 | **7-8**<br>March –<br>July/August, 2022 |
| Period-specific epi-week span | 10-20 | 21-38 | 39-2 | 3-21 | 22-38 | 39-10 | 11-30 |
| Period-specific Δ Bayesian sero-prevalence (%) [a, b,c]<br>(95% CrI) | 0.1<br>(-0.47, 0.8) | 0.2<br>(-0.56, 1.0) | 2.9<br>(1.3, 4.2) | 7.0<br>(5.0, 9.0) | -1.8<br>(-4.2, 0.7) | 33.9<br>(30.8, 37.4) | 19.1<br>(15.3, 23.1) |
| **Bayesian SUAR** [b, c ,d]<br>**(95% CrI)** | 11.6<br>(0.4, 37.6) | 3.6<br>(0.2, 8.7) | 2.9<br>(1.7 - 4.0) | 4.0<br>(3.0, 5.1) | 7.3<br>(3.4, 12.1) | 12.4<br>(11.2, 13.7) | 94.3<br>(75.1, 114.6) |

95% CrI = 95% credible interval; SP = sero-prevalence; SUAR = surveillance under-ascertainment ratio

Sensitivity and specificity considerations addressed in **Supplementary Material 2**. See **Supplementary Material 3** for methodological details related to SUAR estimation. Population estimates include long-term care facility (LTCF) and assisted (ALF) or independent living facility (ILF) residents; whereas, sero-survey sampling and surveillance case report tallies excluded these individuals.

[a] Infection-induced sero-prevalence based on dual-assay positivity, of which from January 2021 at least one positive assay must include anti-nucleocapsid protein detection. Bayesian estimates age, sex, and HA standardized and additionally adjusted for sensitivity and specificity.
[b] Assuming no previously infected are re-infected during the specified analysis period
[c] Age, sex and HA standardized and adjusted for sensitivity and specificity.
[d] Period-specific SUAR estimates exclude samples from the posterior where the difference in sero-prevalence between sero-surveys is less than zero.
